# Impact of unequal cluster sizes for GEE analyses of stepped wedge cluster randomized trials with binary outcomes

**DOI:** 10.1101/2021.04.07.21255090

**Authors:** Zibo Tian, John Preisser, Denise Esserman, Elizabeth Turner, Paul Rathouz, Fan Li

**Author notes:** Corresponding author: Fan Li.

## Abstract

The stepped wedge design is a type of unidirectional crossover design where cluster units switch from control to intervention condition at different pre-specified time points. While a convention in study planning is to assume the cluster-period sizes are identical, stepped wedge cluster randomized trials (SW-CRTs) involving repeated cross-sectional designs frequently have unequal cluster-period sizes, which can impact the efficiency of the treatment effect estimator. In this article, we provide a comprehensive investigation of the efficiency impact of unequal cluster sizes for generalized estimating equation analyses of SW-CRTs, with a focus on binary outcomes as in the Washington State Expedited Partner Therapy trial. Several major distinctions between our work and existing work include: (i) we consider multilevel correlation structures in marginal models with binary outcomes; (ii) we study the implications of both the between-cluster and within-cluster imbalances in sizes; and (iii) we provide a comparison between the independence working correlation versus the true working correlation and detail the consequences of ignoring correlation estimation in SW-CRTs with unequal cluster sizes. We conclude that the working independence assumption can lead to substantial efficiency loss and a large sample size regardless of cluster-period size variability in SW-CRTs, and recommend accounting for correlations in the analysis. To improve study planning, we additionally provide a computationally efficient search algorithm to estimate the sample size in SW-CRTs accounting for unequal cluster-period sizes, and conclude by illustrating the proposed approach in the context of the Washington State study.

## 1 Introduction

The stepped wedge (SW) design is a type of unidirectional crossover design where cluster units switch from control to intervention condition at different pre-specified time points, or steps (Hussey and Hughes, 2007; Turner et al., 2017). This design has been increasingly adopted in cluster randomized trials (CRTs), where the unit of randomization is often a group of individuals such as hospitals or clinics (Barker et al., 2016). In a typical SW-CRT, the intervention is scheduled to be implemented in only a small fraction of the clusters at each step, which is often logistically more feasible compared to concurrently implementing the intervention within half of the clusters as in a typical parallel arms CRT design. In addition, SW-CRT allows all participating clusters to receive the intervention prior to the end of the study, and may facilitate study participant recruitment when stakeholders perceive the intervention to be beneficial to the cluster population (Prost et al., 2015).

Methods for planning SW-CRTs, especially those associated with sample size and power calculations, have been an active topic for statistical research over the past decades (Hemming et al., 2015; Hooper et al., 2016; Kasza et al., 2019; Li et al., 2018b; Li, 2020). However, a convention in deriving sample size formulae is to assume that the number of observations in each cluster-period (referred to as the cluster-period size) is the same both within and between clusters. This equal cluster-period size assumption, while operationally convenient for study planning, is often questionable, especially in cross-sectional designs where different individuals present for health care in each cluster during each period. For example, the Washington State Expedited Partner Therapy (EPT) trial rolled out an partner therapy intervention in 22 local health jurisdictions (LHJ) over 4 steps, where women attending sentinel clinics in each six-month time period were tested for chlamydia and Gonorrhea infection (Golden et al., 2015). Figure 1 presents a cluster-by-period diagram of this study, along with the cluster-period sizes. While the average cluster-period size is roughly 300, the actual cluster-period sizes range from 17 to 1553 across 22 *×* 5 = 110 cluster-periods.

**Figure 1.**
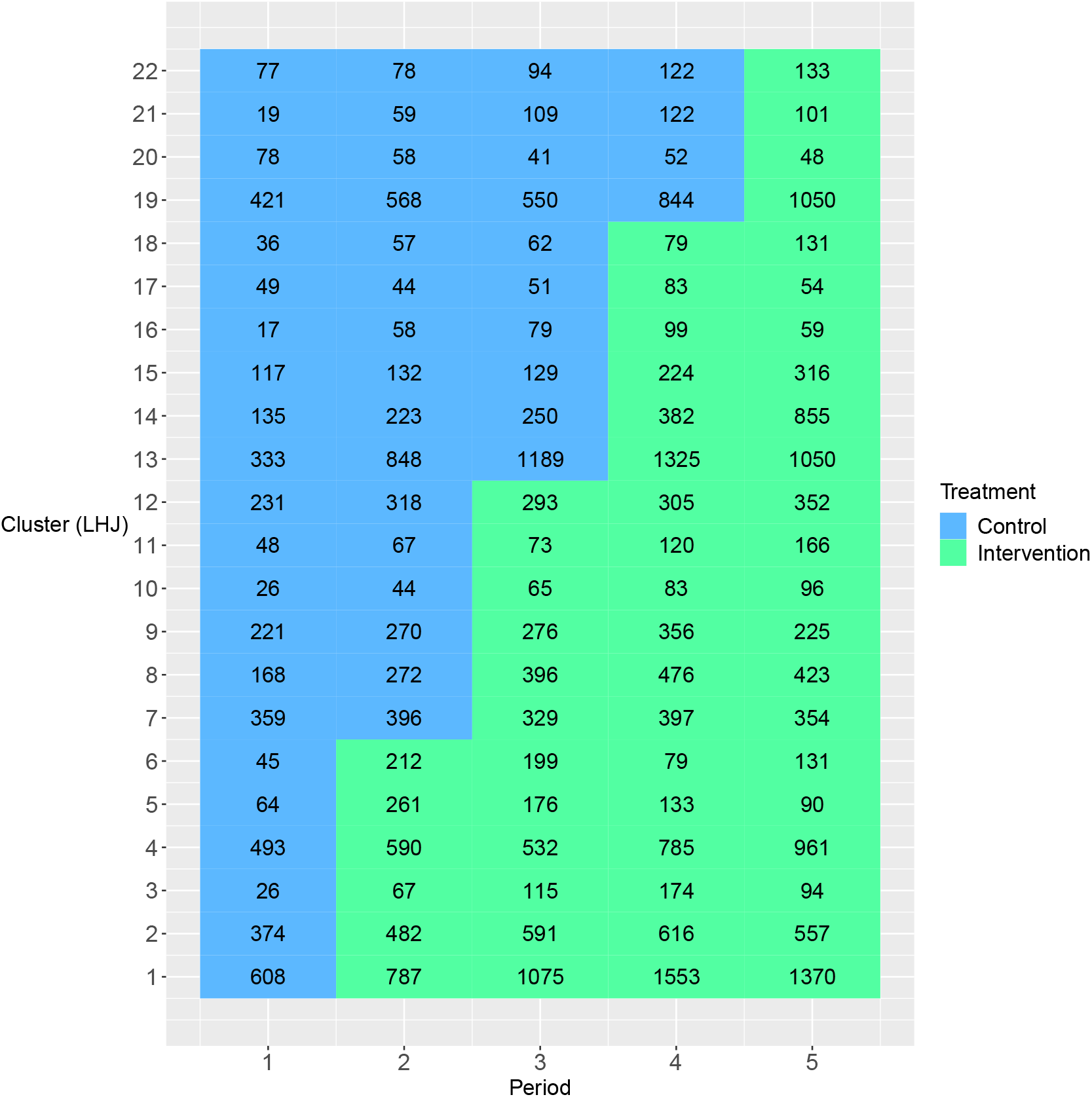
The cluster-by-period diagram for the Washington State Expedited Partner Therapy (EPT) trial. Local health jurisdictions (LHJ) are the clusters in this trial. Each cell represents a cluster-period along with its cluster-period size. The blue color and green color indicate the control and intervention condition, respectively.

While the impact of unequal cluster sizes has been well studied for continuous, binary and count outcomes in parallel CRTs (Kerry and Bland, 2001; Manatunga et al., 2001; Eldridge et al., 2006; van Breukelen et al., 2007; Candel and van Breukelen, 2010; Liu and Colditz, 2018; Li and Tong, 2021a,b), there are a limited number of studies investigating the impact of unequal cluster sizes in SW-CRTs, all of which are restricted to continuous outcomes. For example, Kristunas et al. (2017) studied the impact of unequal cluster sizes in SW-CRTs via simulations and found cluster size imbalances did not lead to notable loss in power. Martin et al. (2019) designed a series of simulations to study the relative efficiency (RE) of a linear mixed model treatment effect estimator under equal versus unequal cluster sizes. They concluded that the median RE is smaller in SW-CRTs compared to parallel CRTs, while the variation of RE can be substantially larger in SW-CRTs. Assuming a more general linear mixed model, Girling (2018) developed an analytical formula for RE in SW-CRTs when the randomization is stratified by cluster size. Harrison et al. (2019) proposed analytical formulae as a function of the mean and variance of the cluster size based on the Hussey and Hughes (2007) linear mixed model for efficient sample size determination in SW-CRTs. Matthews (2020) considered optimal SW-CRTs that achieve the smallest variance of the treatment effect estimator under unequal cluster sizes. Despite these efforts, there is currently limited empirical evidence for the RE in SW-CRTs with binary outcomes, whereas binary outcomes are of interest in the Washington State EPT trial, and are also fairly common according to the systematic review by Barker et al. (2016). Furthermore, the sample size formulae developed for continuous outcomes in SW-CRTs can lead to inaccurate approximations when the outcomes are binary, even under equal cluster sizes (Zhou et al., 2020). Therefore, new sample size procedures that explicitly account for the mean-variance relationship of binary outcomes as well as unequal cluster sizes are needed.

Generalized linear mixed models and marginal models represent two mainstream approaches for analyzing SW-CRTs with binary outcomes. Because SW-CRTs are often used in health care research to inform policy decisions, marginal models, which carry a population-averaged interpretation, may be preferred (Drum et al., 1993; Preisser et al., 2003; Li et al., 2018b) Additional advantages of marginal models for analyzing SW-CRTs were summarized in Li et al. (2021). In this article, we aim to study the impact of unequal cluster sizes for marginal model analysis of SW-CRTs such as the Washington State EPT study, with the purpose to inform study planning. Several major distinctions between our work and existing work on unequal cluster sizes for SW-CRTs (Kristunas et al., 2017; Martin et al., 2019; Girling, 2018; Harrison et al., 2019) include (i) we consider multilevel correlation structures in the context of binary outcomes arising from SW-CRTs, including the nested exchangeable and the exponential decay structure (Kasza et al., 2019; Li et al., 2021); while most previous efforts restrict to continuous outcomes with an overly simplified exchangeable correlation structure; the limitations of the exchangeable correlation structure for SW-CRT applications have been pointed out by Taljaard et al. (2016) and Li et al. (2020); (ii) we study the implications of both the between-cluster and within-cluster imbalances in sizes, as opposed to previous efforts that exclusively focus on the between-cluster variability; and (iii) we provide a comparison between the independence working correlation and the true working correlation and study the consequence of ignoring correlation estimation in SW-CRTs with unequal cluster sizes. GEEs with independence working correlation structure has been studied, for example, in Wang (2019) for designing SW-CRTs, and in Thompson et al. (2020) for analyzing SW-CRTs. Although the independence working assumption offers computational convenience and simplicity, we will show that it can lead to dramatic efficiency loss in SW-CRTs with unequal cluster sizes. Finally, we also introduce a computationally efficient Monte Carlo approach to estimate the sample size for SW-CRTs with binary outcomes with unequal cluster sizes.

The outline of this paper is as follows. Section 2 reviews the individual-level and cluster-period-level generalized estimating equations (GEE) methods used to estimate treatment effect parameter in SW-CRTs with binary outcomes. Section 3 defines the RE of unequal versus equal cluster-period sizes for treatment effect estimation, and introduces a special result on RE under a three-period SW design. Section 4, 5 and 6 present our simulation design and results on RE in SW-CRTs. We provide a Monte Carlo sample size method and demonstrate its application to our motivating trial in Section 7. Section 8 summarizes the key observations and discusses connections between this article and the previous works.

## 2 Marginal models for stepped wedge designs with binary outcomes

### 2.1 Marginal model with individual-level observations

We consider a cross-sectional SW-CRT with *I* clusters and *J* periods, where different sets of individuals are included in each period and their outcome measurements are taken at the end of that period. Let *y*_*ijk*_ be the binary outcome of individual *k* = 1, …, *n*_*ij*_ from cluster *i* = 1, …, *I* during period *j* = 1, …, *J*. We assume a complete design so that outcomes are taken for all individuals in each period (Hemming et al., 2015). Let *µ*_*ijk*_ be the marginal mean outcome; the marginal model for an SW-CRT was studied in Li et al. (2018b); Ford and Westgate (2020) and relates the marginal mean to the period effect and treatment via the following generalized linear model

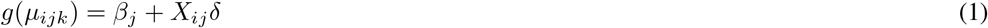

where *g* is a link function, *β*_*j*_ is the *j*-th time effect describing the secular trend, *X*_*ij*_ is the treatment indicator that equals to 1 if cluster *i* receives treatment during period *j* and 0 otherwise, and *δ* denotes the treatment effect of interest. For example, if *g* is chosen as the identity link, model (1) is a linear probability model and *δ* measures the time-adjusted risk difference; if *g* is chosen as the log link, model (1) is a log-binomial model and exp(*δ*) is interpreted as the time-adjusted relative risk; and if *g* is the canonical logit link, then model (1) is a logistic model and exp(*δ*) is interpreted as the time-adjusted odds ratio.

Because outcomes in a SW-CRT are correlated within each cluster, appropriate within-cluster correlation structures are required to characterize their covariance. We consider two multilevel correlation structures developed for cross-sectional SW-CRTs: the nested exchangeable (NEX) correlation structure (Li et al., 2018b) and the exponential decay (ED) correlation structure (Kasza et al., 2019; Li et al., 2021). Both correlation structures distinguish between the within-period and between-period intraclass correlation coefficients (WP-ICC and BP-ICC) compared to the simple exchangeable correlation structure (Hussey and Hughes, 2007) and has been considered to be more realistic (Taljaard et al., 2016; Li et al., 2020). Under the NEX correlation structure, we define *α*_0_ as the WP-ICC that measures the correlation between two outcomes from different individuals within the same cluster-period, i.e., corr(*y*_*ijk*_, *y*_*ijk′*_) = *α*_0_ for *k* ≠ *k′*. Further, we define the *α*_1_ as the BP-ICC that measures the correlation between two outcomes from two different cluster-periods, i.e., corr(*y*_*ijk*_, *y*_*ij′ k′*_) = *α*_1_ for *j* ≠ *j′, k* ≠ *k′*. Under the ED correlation structure, the WP-ICC is still defined as *α*_0_, whereas the BP-ICC is allowed to exponentially decay over time, i.e., corr(*y*_*ijk*_, *y*_*ij′* *k′*_) = *α*_0_*ρ*^|*j−j′*|^ for *j* ≠ *j′, k* ≠ *k′* given the decay parameter *ρ ∈* [0, 1]. In matrix notation, if we write 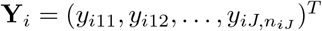 as the collection of outcomes in cluster *i* ordered by period, the NEX correlation structure is given by

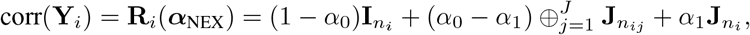

where ***α***_NEX_ = (*α*_0_, *α*_1_)^*T*^, **I**_*s*_ is the *s × s* identity matrix, **J**_*s*_ is the *s × s* matrix of ones, 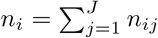 is the *i*-th cluster size, and ‘⊕’ is the block diagonal operator. With arbitrary cluster-period sizes, there exists a closed-form inverse of the NEX correlation matrix. We derive the explicit expression in Web Appendix A, which generalizes an earlier expression derived by Li et al. (2019) for *J* = 2. On the other hand, the ED structure is given by

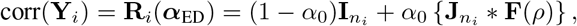

where ***α***_ED_ = (*α*_0_, *ρ*)^*T*^, **F**(*ρ*) is the *J ×J* first-order auto-regressive (AR-1) correlation matrix, ‘*∗*’ denotes the Khatri-Rao product operator (Khatri and Rao, 1968) applied on each *n*_*ij*_ *×n*_*ij′*_ block of 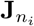 and each scalar element of **F**(*ρ*). Unlike the NEX structure, a closed-form inverse of the ED correlation matrix is not available. Of note, the simple exchangeable correlation structure implied by the Hussey and Hughes (2007) model is obtained when *α*_1_ = *α*_0_ under the NEX structure or *ρ* = 1 under the ED structure.

Generalized estimating equations (Liang and Zeger, 1986) are often used to estimate the treatment effect parameter *δ* in marginal model (1). Defining 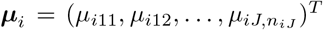 as the collection of marginal means in cluster *i* and mean model regression parameter ***θ*** = (*β*_1_, …, *β*_*J*_, *δ*)^*T*^, then the GEE with individual-level observations is written as

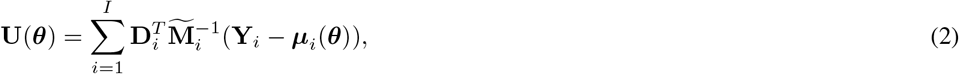

where 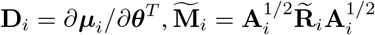 is the working variance, with 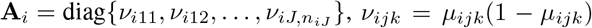, and 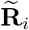 as the working correlation model. When the working independence assumption is adopted and 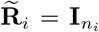, the working correlation is misspecified when the truth is otherwise, either NEX or ED in the current study, but 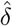 is still a consistent estimator of the treatment effect (Liang and Zeger, 1986). In this case, the large-sample variance of 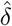 can be obtained as the (*J* + 1, *J* + 1)-th element of the sandwich variance matrix 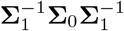, where 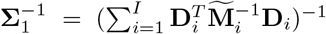 and 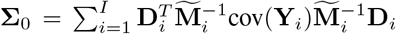. Alternatively, when 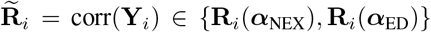 and the correlation structure is correctly specified, 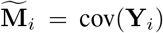 and the large-sample variance of 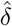 can be obtained as the (*J* + 1, *J* + 1)-th element of the model-based variance matrix 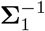. Details of the individual-level GEE approaches that simultaneously estimates ***θ*** and ICC parameters were developed elsewhere (Prentice, 1988; Preisser et al., 2008; Li et al., 2018b; Li, 2020). The left column of Table 1 provides example matrix representations of different working correlation models, 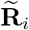, for a hypothetical cluster with 3 periods.

**Table 1.**
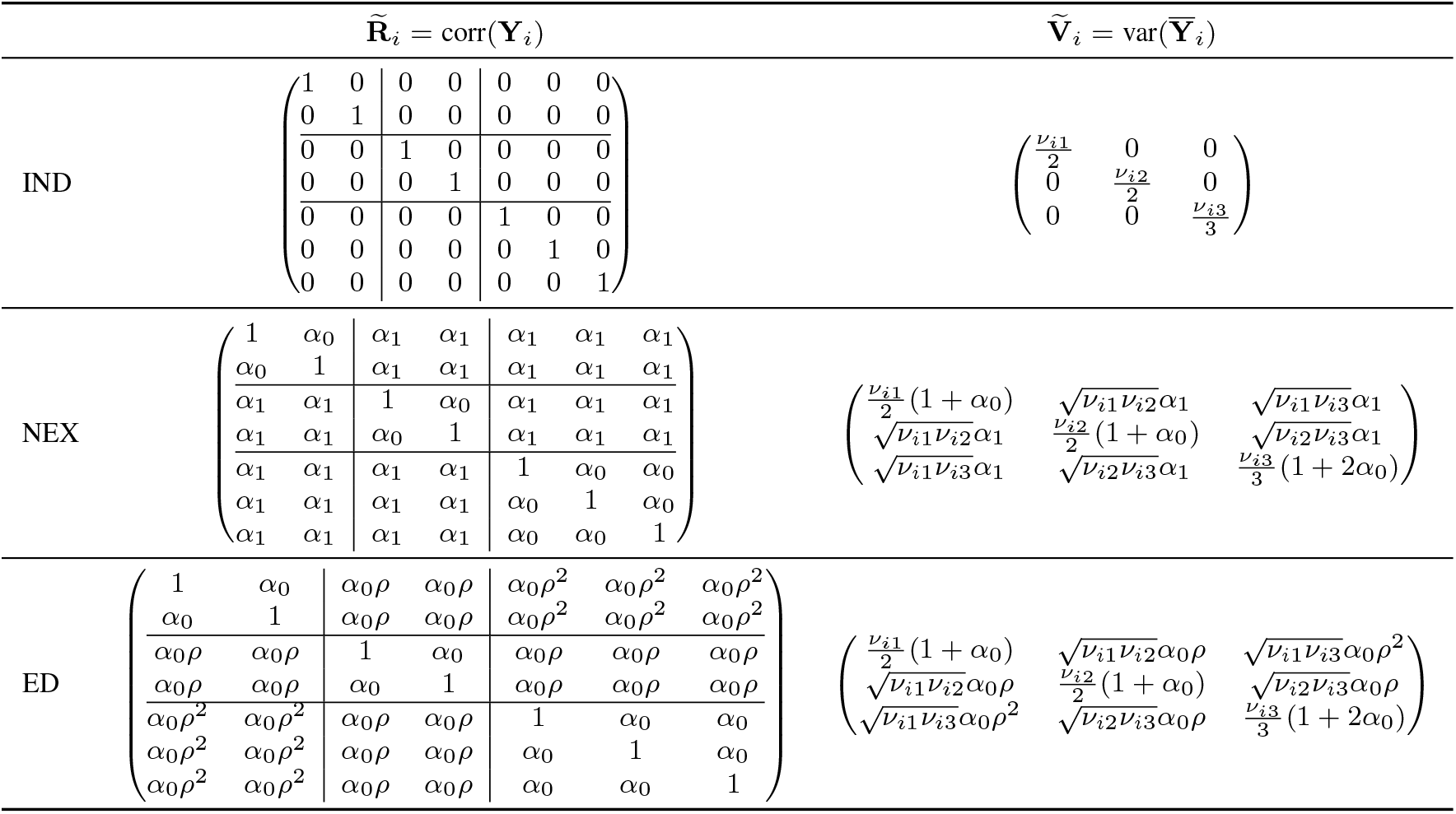
Examples of the working correlation matrix for the individual-level observations (left column) and the corresponding cluster-period-level working covariance matrix (right column) under the independence (IND), nested exchangeable (NEX) and exponential decay (ED) working assumptions. The illustration is based on a stepped wedge trial with *J* = 3 periods and (*n*_*i*1_, *n*_*i*2_, *n*_*i*3_) = (2, 2, 3) observations for cluster *i* with **Y**_*i*_ = (*y*_*i*11_, *y*_*i*12_, *y*_*i*21_, *y*_*i*22_, *y*_*i*31_, *y*_*i*32_, *y*_*i*33_)^*T*^.

### 2.2 Marginal model with cluster-period means

While the marginal model (1) provides a good basis for the design and analysis of SW-CRTs with binary outcomes, the GEE procedure based on (2) may be computationally intensive as one needs to invert **M**_*i*_ and **R**_*i*_, which may have quite sizable dimensions as in Figure 1. To circumvent computationally challenges, Li et al. (2021) proposed a cluster-period GEE approach. Specifically, we define the vector of cluster-period means as

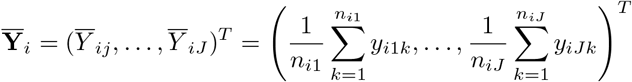

and the marginal mean of 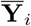 as 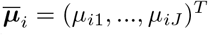. Then the individual-level marginal mean model (1) can be equivalently represented by

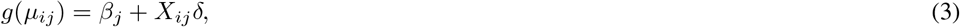

where *β*_*j*_ and *δ* can preserve their original interpretations. The cluster-period GEE is then represented as

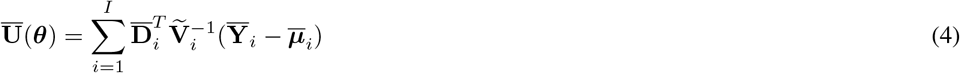

where 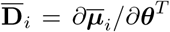 and 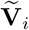 is the working covariance matrix for the cluster-period mean 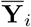, which is only of dimension *J × J*. In particular, the working variance 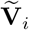 depends on the variance function, cluster-period sizes as well as the working correlation structure. In parallel to Section 2.1, if 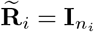 and the independence working assumption is adopted, then **V**_*i*_ = diag{*ν*_*i*1_/*n_i1_*, …, *ν*_*iJ*_ /*n*_*iJ*_}, where *ν*_*ij*_ = *µ*_*ij*_(1 *− µ*_*ij*_). Because the independence working correlation model is likely misspecified, the large-sample variance of 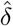 is obtained as the (*J* + 1, *J* + 1)-th element of the sandwich variance matrix 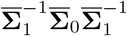, where 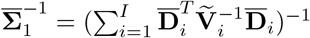 and 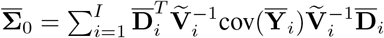. On the other hand, if the working correlation structure is the NEX or ED structure, the *j*-th diagonal element of 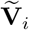 is 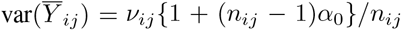. Furthermore, the (*j, j′)-th* off-diagnal element of 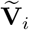 is 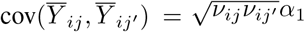 under the NEX correlation structure, and 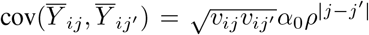 under the ED correlation structure. In these cases, if the working correlation model is also correctly specified, then the large-sample variance of 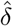 is obtained as the (*J* + 1, *J* + 1)-th element of the model-based variance matrix 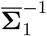. In particular, the cluster-period GEE approach for simultaneously estimating ***θ*** and ICCs based on cluster-period means was developed in Li et al. (2021). The right column of Table 1 provides example matrix representations of different working variance models, 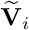, for a hypothetical cluster of 3 periods.

For numerically evaluating RE for marginal analyses of SW-CRTs under unequal cluster sizes, we will make use of the following Theorem 2.1 established in Li et al. (2021).

#### Theorem 2.1

*(Li et al*., *2021) With the same choice of working correlation model* 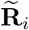 *(independence, nested exchangeable or exponential decay) and compatible marginal mean models* (1) *and* (3), *the individual-level GEE and the cluster-period GEE have the same model-based variance and the sandwich variance, even under unequal cluster-period sizes. In other words*, 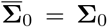, *and* 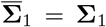, *regardless of the cluster-period size distribution*.

Theorem 2.1 shows that there is no loss of asymptotic efficiency by replacing the individual-level GEE with the cluster-period GEE under unequal cluster-period sizes, and doing so simplifies the computation of RE for estimating *δ*. For this reason, we will define RE in Section 3 based on the cluster-period GEE. As will be seen in due course, the computation associated with the cluster-period GEE is much faster given we only need to invert a *J × J* matrix 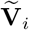 rather than the 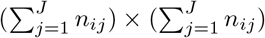 matrix M_*i*_. This insight also motivates the computationally efficient Monte Carlo sample size approach for SW-CRTs with binary outcomes in Section 7.

## 3 Relative efficiency of unequal versus equal cluster sizes

We define RE as the relative variance of the treatment effect estimator based on the cluster-period GEE under unequal versus equal cluster sizes. For equal cluster sizes, we only consider the scenarios where all cluster-period sizes are equal. For unequal cluster size scenarios, we consider two types of cluster-period size variability: (i) the cluster-period sizes are the same within each cluster, but differ across clusters (between-cluster imbalance) and (ii) the cluster-period sizes are different both within each cluster and across clusters (between-within-cluster imbalance). Let **Ω**_equal_ denote a design with equal cluster sizes, and **Ω**_unequal_ denote a design with unequal cluster-period sizes. The RE of equal to unequal cluster sizes is written as

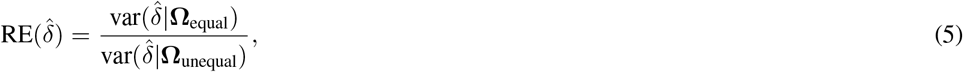

where the variance of 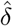 can be the model-based or sandwich variance, depending on the choice of the working correlation structure. With a continuous outcome and identity link function, the asymptotic variances of the treatment effect estimator based on GEE and linear mixed model coincide (Li et al., 2018b), and the analytical results on RE developed in Girling (2018) can be applied. However, binary outcomes have an explicit mean-variance relationship and therefore generally prohibit an analytical derivation of a scalar variance expression. Therefore, we will numerically study the trends and magnitude of RE under a variety of design configurations in Section 4, 5 and 6.

While a simple analytical expression for RE is intractable with binary outcomes, we are able to obtain an interesting result on RE under the basic three-period design, when the treatment effect is estimated under working independence assumption. We summarize the result in Theorem 3.1, with the detailed derivations in Web Appendix B.

### Theorem 3.1

*In a stepped wedge design with three time periods and hence two treatment sequences, if the true correlation structure is either nested exchangeable or exponential decay, the sandwich variance of the cluster-period GEE estimator* 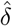 *assuming working independence is given in closed-form by*

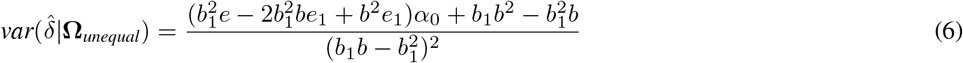

*where* 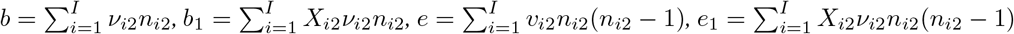, *and ν*_*ij*_ = *µ*_*ij*_(1 − *µ*_*ij*_) *is the variance function for binary outcomes. Further, variance* (6) *does not involve the BP-ICC, nor any information from the first or third periods*.

Theorem 3.1 suggests that the variance of the treatment effect estimator obtained from the independence GEE only depends on the cluster sizes and marginal variance of the outcome in the second period in a three-period design. This result is intuitive because all clusters receive the same intervention during the first and third period and there is no effective information for a between-cluster comparison, whereas the independence GEE heavily relies on between-cluster comparisons. Furthermore, assuming equal cluster-period sizes, we can obtain 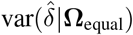 by setting *n*_*i*2_ = *n* for all *i* in (6). Because both 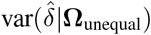 and 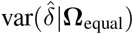 depend on the correlation structure only through *α*_0_, the RE under the independence working assumption does not vary as a function of *α*_1_ (under the NEX structure) or *ρ* (under the ED structure). This RE is also unaffected by any changes in cluster-period sizes during the first and third periods. Collectively, these observations suggest that the RE under the independence working assumption in a three-period design is invariable to correlation decay.

While Theorem 3.1 is simple and intuitive, it does not easily extend to cases where the working correlation structure is correctly specified as the NEX or ED structure, nor where there are more than three periods. This is due to the challenge of analytically inverting an unstructured *J × J* matrix for *J ≥* 4. In what follows, we will numerically evaluate the RE under a wide range of design configurations representing more general cases.

## 4 Simulation design

For binary outcomes, we investigate RE defined in (5) under standard and complete stepped wedge designs, where an equal number of clusters transition from control to intervention at each step. We consider two types of true correlation structures: NEX and ED, defined in Section 2. We study RE assuming a correctly specified working correlation structure as well as an incorrectly specified independence working correlation structure (IND). In the former case, 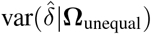 and 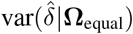 are given by the model-based variance, whereas in the latter case, 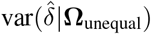 and 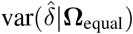 are given by the sandwich variance. Other design factors we consider include number of clusters *I*, number of periods *J* and the degree and type of cluster size variability; other model factors we consider include the true mean model coefficients determining the baseline prevalence, secular trend and the treatment effect, as well as the true ICC parameters ***α***_NEX_ and ***α***_ED_ for the respective true correlation structures. For each parameter combination, we will simulate two designs: one with equal cluster sizes, **Ω**_equal_, and one with unequal cluster sizes, **Ω**_unequal_, and numerically compute RE. The distributions of RE are then summarized across 1000 simulation runs. We follow Martin et al. (2019) and obtain the median and interquartile range (IQR) of REs for each scenario. We focus on the median and IQR to minimize the undue influence of extreme values. Source code to reproduce the simulation results is available as Supporting Information on the journal’s web page.

We consider number of clusters, *I* ∈ {12, 24, 48, 96}, with *I* = 24 resembling the Washington State EPT trial. We consider *J* ∈ {3, 5, 13} periods such that *I* is divisible by the number of steps *J* − 1. For example, when *I* = 24 ad *J* = 13, we assume *I*/(*J* − 1) = 2 randomly selected clusters switch from control to intervention during each post-baseline period.

To simulate unequal cluster sizes, we first consider the simplified scenario with only between-cluster imbalance, but homogeneous cluster-period sizes within each cluster. To do so, we assume 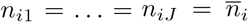 for each cluster *i*, and generate 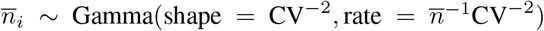, where 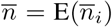 is the mean cluster-period sizes and CV is the coefficient of variation. We focus on 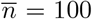; results for 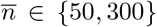 are discussed in Section 5.5. The between-cluster imbalance is measured by CV *∈* {0.25, 0.5, 0.75, 1, 1.25, 1.5}; these values were also explored in Martin et al. (2019) and Harrison et al. (2019) for continuous outcomes under simpler correlation structures. For computational stability, we round each 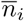 to the nearest integer and set 5 as the lower bound. To ensure better comparability between the two designs, **Ω**_unequal_ and **Ω**_equal_, we scaled the simulated cluster-period sizes proportionally such that the total number of observations across all cluster-periods is 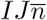. This procedure minimizes the difference in total sample size between **Ω**_equal_ and **Ω**_unequal_, which then guarantees the difference between the variance of estimated treatment effect under various designs is only attributable to the variability in cluster sizes rather than total sample size.

We further consider between-within cluster imbalance by allowing the cluster-period size to vary within each cluster. Conditional on the simulated mean cluster-period sizes 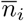, we generate the actual cluster-period sizes (*n*_*i*1_, …, *n*_*iJ*_) from a truncated multinomial distribution with number of trials 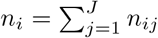 and a pre-specified probability vector (*p*_1_, …, *p*_*J*_), where 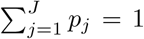. The truncated multinomial distribution is used to ensure the smallest cluster-period size is at least 2. The following four different specifications of (*p*_1_, …, *p*_*J*_) represent four different recruitment patterns that lead to unequal cluster-period sizes:

1. Constant: *p*_1_ = *p*_2_ = … = *p*_*J*_. This pattern assumes the absence of any systematic variation of cluster-period sizes for each cluster; any variation in cluster-period sizes is only due to chance in balanced multinomial sampling;
2. Monotonically increasing: *p*_1_ < *p*_2_ *<* … < *p*_*J*_. This pattern assumes that there is an increasing effort in recruitment leading to larger cluster-period sizes at the later time periods. Specifically, we define a difference parameter *d* such that *p*_*j*_ = *p*_1_ + (*j −* 1) *× d* with *j* = 1, …, *J*. With the initial probability *p*_1_ known, *d* = 2(1 *− Jp*_1_)/*{J* (*J −* 1)};
3. Monotonically decreasing: *p*_1_ *> p*_2_ *>* … *> p*_*J*_. This pattern assumes a scenario where recruitment of patients become more challenging over time and the cluster-period sizes on average decrease at the later time periods. Operationally, this is done by reversing the corresponding vector obtained under the monotonically increasing pattern;
4. Randomly permuted: perm {*p*_1_ < *p*_2_ *<* … < *p*_*J*_}. A probability vector (*p*_1_, …, *p*_*J*_) that satisfies the monotonically increasing pattern is obtained. Then a random permutation of (*p*_1_, …, *p*_*J*_) is used to simulate cluster-period sizes for each cluster. This pattern yields a more chaotic and non-monotone within-cluster imbalance.

When simulating the four patterns of between-within-cluster imbalance, we specify *p*_1_ = {0.2, 0.1, 0.05} for *J* = {3, 5, 13} respectively. Besides cluster-period sizes *n*_*ij*_ in the case of unequal cluster sizes, no other data are simulated considering that 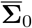 and 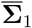 are computed using analytical calculations.

Finally, we choose several typical model parameters for our evaluation. We assume the logistic marginal mean model where the baseline prevalence exp(*β*_1_)/{1 + exp(*β*_1_)} = 0.3, and no true secular trend such that *β*_1_ = … = *β*_*J*_. Results with a smaller baseline prevalence 0.1, increasing or decreasing secular trend are presented in Section 5.6. We assume the intervention effect exp(*δ*) = 0.35; results under a smaller non-null intervention effects are compared with the previous *δ* = log(0.35) in the same section. For the true ICC parameters under the NEX or ED correlation structures, we consider the WP-ICC *α*_0_ ≤ 0.2, corresponding to the common range of reported ICCs in CRTs (Murray and Blitstein, 2003; Preisser et al., 2007; Martin et al., 2016). Under both correlation structures, we consider values of *α*_1_ or *ρ* such that the BP-ICC is positive and does not exceed the WP-ICC *α*_0_. Of note, there are natural restrictions of the range of plausible correlation parameters based on the marginal mean, and we have ensured that all combinations of *α*_0_, *α*_1_ or *ρ* used in the scenarios do not violate those restrictions. The specific restrictions of correlation parameters are defined in Qaqish (2003) and re-interpreted under the NEX correlation structure as

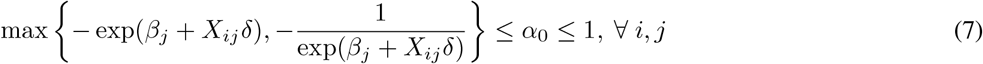

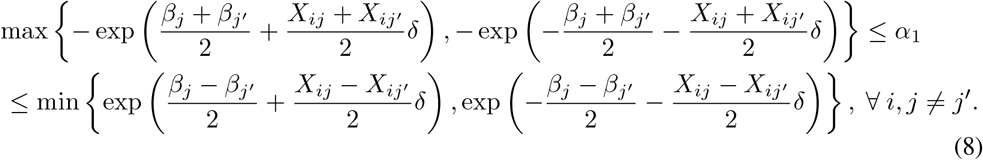

Because we specify *α*_0_ between 0 and 1, restriction (7) always holds and it is more critical to check (8). Furthermore, for the ED correlation structure, we modify restriction (8) by replacing *α*_1_ with *α*_0_*ρ*^|*j−j′*|^.

## 5 Simulation results when the true correlation structure is nested exchangeable

### 5.1 Cluster size variability

Figure 2 presents the median and IQR of RE as a function of CV with *I* = 12 and 96 clusters and *J* = 5 periods. The WP-ICC, *α*_0_, is fixed at 0.05, and three values of BP-ICC, *α*_1_ ∈ {0.001, 0.025, 0.05}, are considered. We first focus on the between-cluster imbalance as measured by the CV of the mean cluster-period sizes 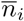, and assume no within-cluster imbalance. When the working correlation structure is correctly specified as NEX, a larger CV leads to a small to moderate efficiency loss for estimating the treatment effect. For example, when *I* = 12, the median RE is around 0.85 when *α*_1_ = 0.001 and CV = 1 in Figure 2 (A). The IQR of RE increases as CV becomes larger, suggesting that the RE is more dispersed over repeated experiments with larger between-cluster imbalance. Similar RE-CV relationships are observed when the working correlation structure is IND; however, the efficiency loss in estimating the treatment effect is much more substantial. For example, when *I* = 12, the median RE drops down to 0.63 when *α*_1_ = 0.001 and CV = 1 in Figure 2 (C).

**Figure 2.**
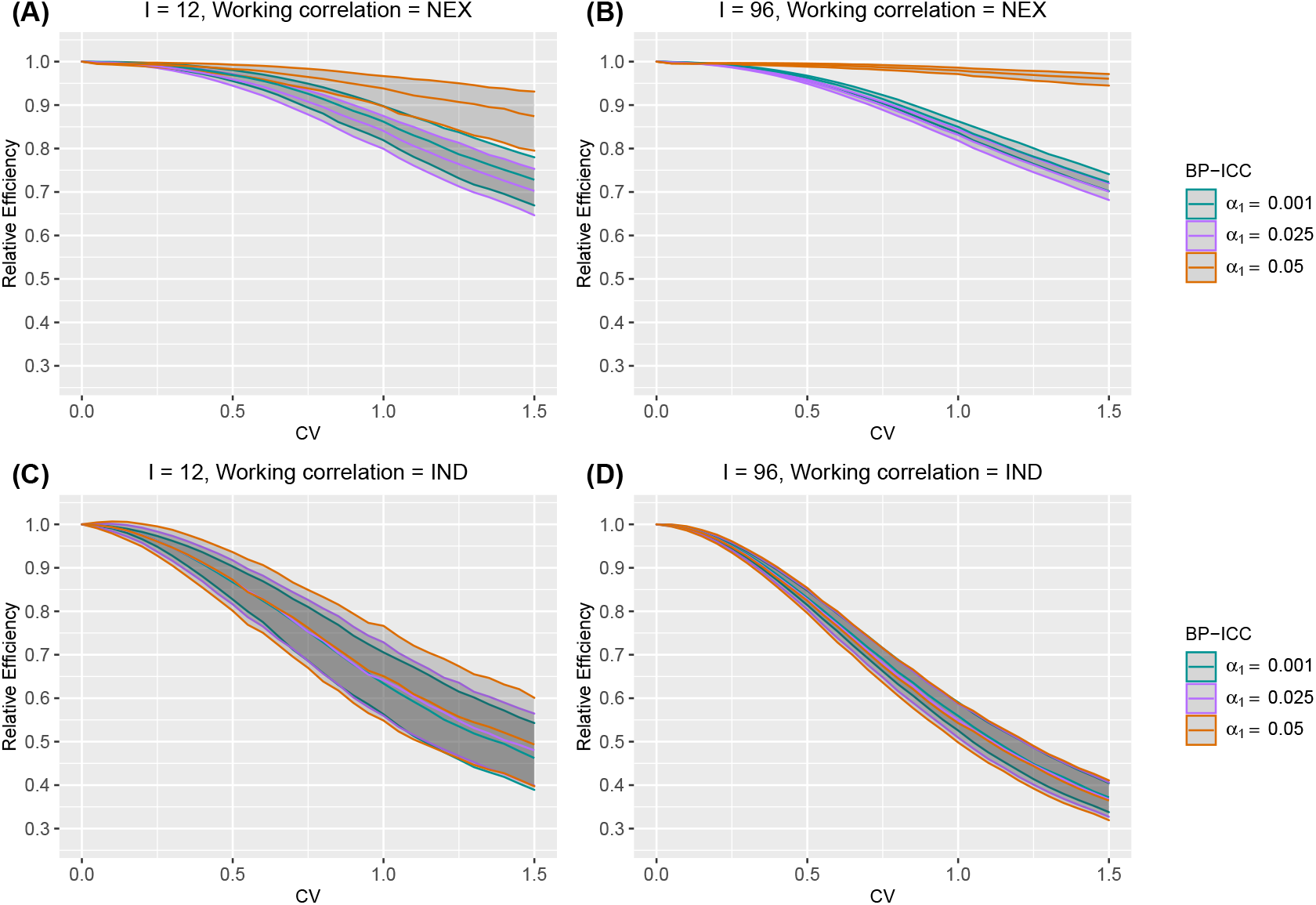
The median and interquartile range (IQR) of relative efficiency (RE) as a function of the coefficient of variation (CV) measuring between-cluster imbalance, when the true correlation model is nested exchangeable (NEX). Design factors considered are as follows: number of clusters *I* = 12 and 96, number of periods *J* = 5. The within-period intraclass correlation coefficient (WP-ICC) *α*_0_ = 0.05, and between-period intraclass correlation coefficient (BP-ICC) *α*_1_ ∈ {0.001, 0.025, 0.05}. The working correlation structure is either NEX or independence (IND). No within-cluster imbalance is introduced.

In the presence of between-cluster imbalance, the RE results are further impacted by the introduction of within-cluster imbalance. Figure 3 presents the counterparts of Figure 2 under the within-cluster imbalance pattern 4 (randomly permuted). Under the NEX working correlation structure, the RE further decreases and appears to be particularly sensitive to within-cluster imbalance when *α*_0_ = *α*_1_, i.e., there is no correlation decay between periods. When *α*_1_ < *α*_0_, the RE results are more robust to within-cluster imbalance, with only a slightly lower median RE and slightly wider IQR at each CV compared to Figure 2 (A-B). Under the IND working assumption, the RE, somewhat counter-intuitively, increases after introducing the within-cluster imbalance when *α*_0_ = *α*_1_ given a fixed level of between-cluster imbalance or CV. As *α*_1_ deviates from *α*_0_, the RE results become insensitive to within-cluster imbalance. This sharp contrast between the behaviour of RE under different working correlation models is further observed for other within-cluster imbalance patterns (see Web Figures 1-3). Among the different within-cluster imbalance patterns, pattern 1 (constant) corresponds to slightly larger RE compared to patterns 2-4, while any difference in RE across patterns 2-4 is negligible.

**Figure 3.**
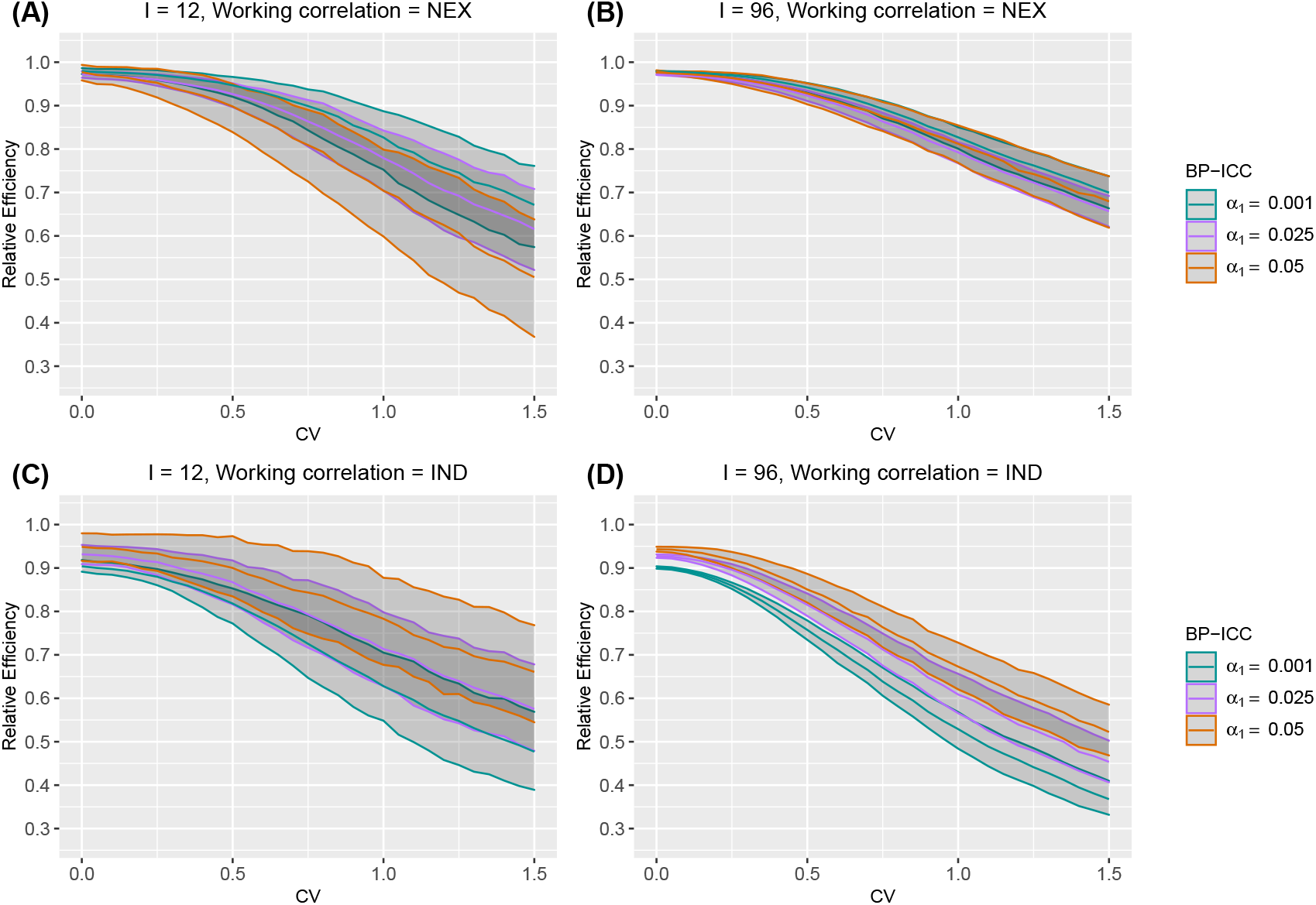
The median and interquartile range (IQR) of relative efficiency (RE) as a function of coefficient of variation (CV) measuring between-cluster imbalance, when the true correlation model is nested exchangeable (NEX). Design factors considered are as follows: number of clusters *I* = 12 and 96, number of periods *J* = 5. The within-period intraclass correlation coefficient (WP-ICC) *α*_0_ = 0.05, and between-period intraclass correlation coefficient (BP-ICC) *α*_1_ ∈ {0.001, 0.025, 0.05}. The working correlation structure is either NEX or independence (IND). Within-cluster imbalance (pattern 4: randomly permuted) is introduced.

### 5.2 Intraclass correlation coefficients

Figure 2 implies that the magnitude of ICCs can affect the median RE of the GEE analysis of stepped wedge trials due to unequal cluster sizes. To provide additional characterization of the RE-ICC relationship under the NEX working correlation structure, Figure 4 presents the median RE as a function of *α*_0_ ∈ {0.01, 0.05, 0.1, 0.2} and *α*_1_/*α*_0_ ∈ [0, 1], across CV ∈ {0.25, 0.75, 1.25} but without within-cluster imbalance. When the working correlation is correctly specified as NEX, the median RE increases with a larger WP-ICC, when the BP-ICC is much smaller than the WP-ICC. However, when the BP-ICC gets closer to the WP-ICC, this relationship can be reversed. This is partly because the relationship between median RE and the BP-ICC can be non-monotone. For example, in Figure 4, the median RE increases monotonically with larger BP-ICCs when the WP-ICC is 0.01. However, when the WP-ICC is at least 0.05, the median RE first sharply decreases and then gradually increases. Both Figure 2 and 4 suggest that the RE-CV relationship heavily depends on the difference between the WP-ICC and the BP-ICC, or the amount of correlation decay, *α*_1_/*α*_0_. For example, when the WP-ICC equals the BP-ICC, namely the true correlation model is simple exchangeable as implied by the classic Hussey and Hughes (2007) model, the efficiency loss due to unequal cluster sizes seems minimal. This agrees with the findings in Kristunas et al. (2017) and Martin et al. (2019), who restricted their investigations to the simple exchangeable correlation model. In general, however, the efficiency loss due to unequal cluster sizes becomes larger when the BP-ICC deviates from the WP-ICC.

**Figure 4.**
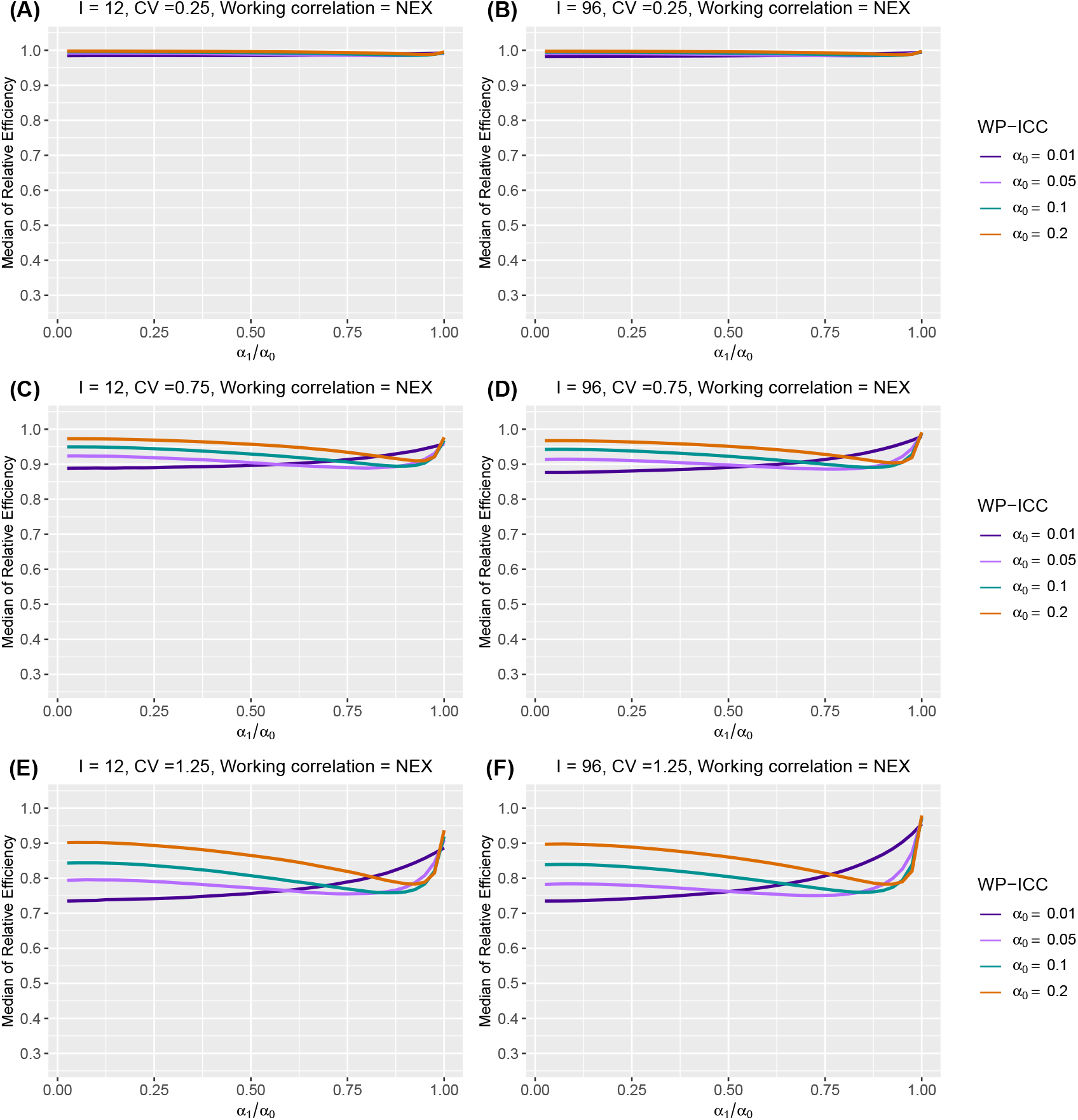
The median of relative efficiency (RE) as a function of the within-period intraclass correlation coefficient (WP-ICC) *α*_0_ ∈ {0.01, 0.05, 0.1, 0.2} and the ratio of between-period intraclass correlation coefficient (BP-ICC) to WP-ICC, *α*_1_/*α*_0_ ∈ [0, 1], when both the true correlation model and the working correlation model are nested exchangeable (NEX). Design factors considered are as follows: number of clusters *I* = 12 and 96, number of periods *J* = 5, and the degree of between-cluster imbalance is defined by CV ∈ {0.25, 0.75, 1.25}. No within-cluster imbalance is introduced.

Web Figures 4-7 present the counterparts to Figure 4, with the introduction of the four within-cluster imbalance patterns. As expected, the median RE becomes smaller when the cluster-period sizes are different within each cluster. Of note, the median RE decreases most dramatically when *α*_0_ = *α*_1_, suggesting that the simple exchangeable correlation model is most prone to efficiency loss as a result of within-cluster imbalance, but is relatively robust to between-cluster imbalance.

Figure 5 presents the counterpart of Figure 4 when the working correlation is IND. Different from the RE-ICC relationship under the NEX working correlation structure, the median RE monotonically decreases with a larger WP-ICC under the IND working correlation structure. The median RE is also insensitive to between-period ICC under the working independence assumption. This is not surprising, as in the special case of *J* = 3 periods, Theorem 3.1 points out that the distribution of RE is independent of *α*_1_. However, the BP-ICC, *α*_1_, plays a more prominent role in determining the RE in the presence of within-cluster imbalance. For example, Web Figure 8-11 shows that, with different within-cluster imbalance patterns, the median RE under the IND working structure becomes a mildly increasing function of the BP-ICC *α*_1_, for fixed values of *α*_0_.

**Figure 5.**
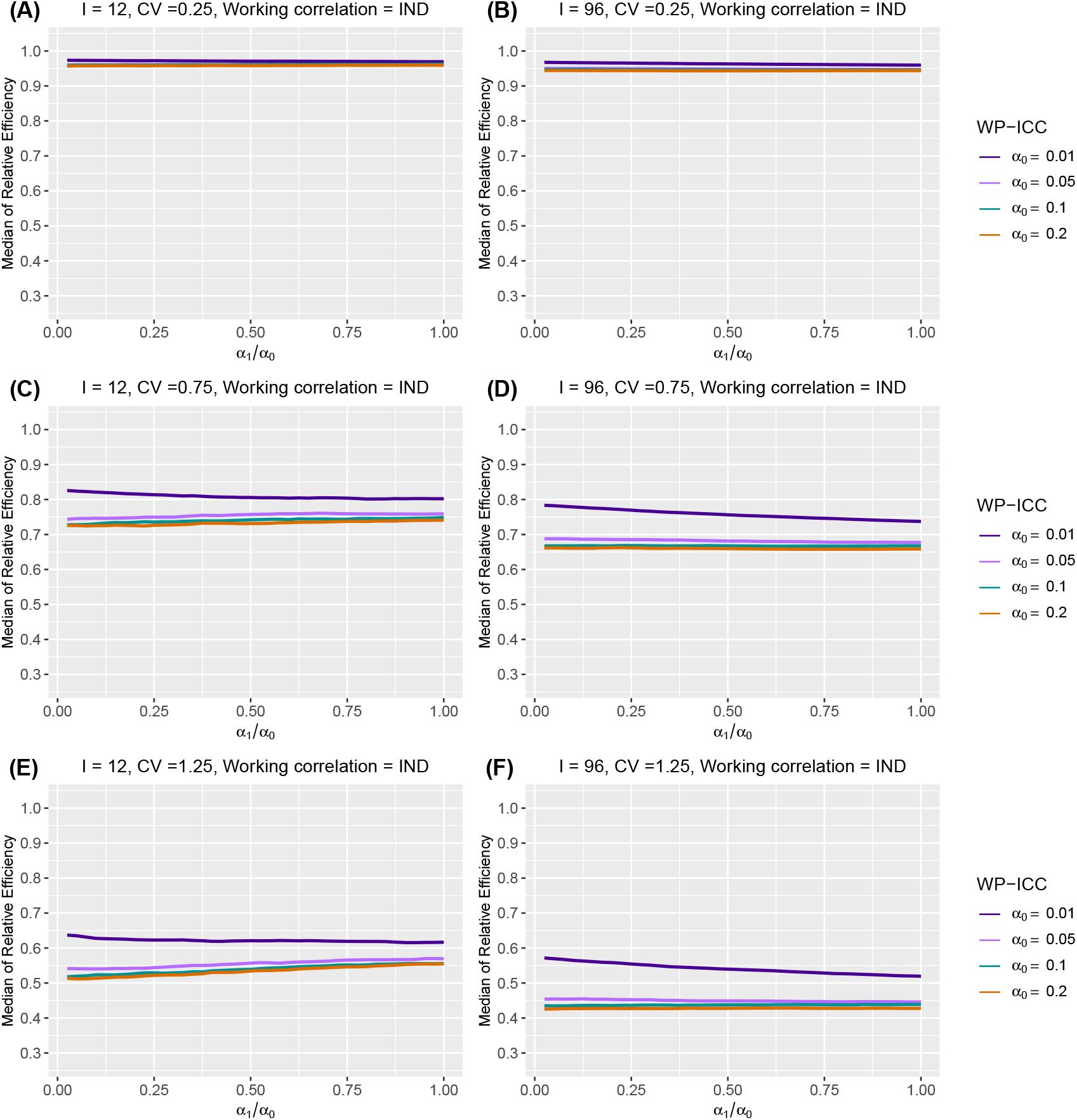
The median of relative efficiency (RE) as a function of the within-period intraclass correlation coefficient (WP-ICC) *α*_0_ ∈ {0.01, 0.05, 0.1, 0.2} and the ratio of between-period intraclass correlation coefficient (BP-ICC) to WP-ICC, *α*_1_/*α*_0_ ∈ [0, 1], when the true correlation model is nested exchangeable (NEX) and the working correlation model is independence (IND). Design factors considered are as follows: number of clusters *I* = 12 and 96, number of periods *J* = 5, and the degree of between-cluster imbalance is defined by CV ∈ {0.25, 0.75, 1.25}. No within-cluster imbalance is introduced.

### 5.3 Number of clusters

Web Figure 12 presents the counterparts of Figure 2 but with *I* = 24 and 48. When the working correlation structure is NEX, we observe a larger number of clusters *I* leads to a more concentrated distribution of REs. When *α*_1_ is close to *α*_0_, or when the true correlation structure is nearly simple exchangeable, the median RE increases most notably when *I* increases, indicating that a larger sample size effectively prevents efficiency loss due to between-cluster imbalance in the absence of between-period correlation decay. The change in median RE due to larger *I*, however, is almost negligible as *α*_1_ deviates from *α*_0_. The same pattern persists even after the introduction of within-cluster imbalance. On the other hand, the impact of number of clusters on RE is completely different when the working correlation structure is IND. When *I* increases from 12 to 96, even though the IQR of RE becomes smaller, the median RE decreases especially when the BP-ICC is large. This suggests that a larger stepped wedge trial is more susceptible to efficiency loss due to between-cluster imbalance if it is analyzed by an independence GEE, and when the BP-ICC is not negligible. The same pattern persists after introducing the within-cluster imbalance, and we conclude that ignoring ICC estimation induces the greatest efficiency loss when both *I* and *α*_1_ become large.

### 5.4 Number of periods

Table 2 summarizes the median and IQR of RE as a function of different number of periods *J* and CV under two different working correlation specifications, with and without the within-cluster imbalance, when there are *I* = 24 clusters (mimicking the Washington State EPT study). For illustration, we choose *α*_0_ = 0.05 and *α*_1_ = 0.025. We omitted the within-cluster imbalance pattern 3 (monotonically decreasing), because the RE results are almost identical to those under pattern 2 (monotonically increasing). As long as the working correlation is NEX, the number of periods *J* has negligible effect on the median and IQR of REs. The results are also not sensitive to within-cluster imbalance. However, when the IND working correlation structure is considered, although the number of periods has minimum effect on RE with only between-cluster imbalance, a trial with a longer duration can partially mitigate the efficiency loss in the presence of additional within-cluster imbalance. The median and IQR of RE can increase substantially with a larger *J* under any of the within-cluster imbalance patterns, when there is already moderate to large between-cluster imbalance. Web Tables 1 to 3 present the corresponding results with *I* = 12, 48 and 96 and the conclusions are identical.

**Table 2.** Median and interquartile range (IQR) (in parentheses) of relative efficiency (RE) as a function of periods *J*, under different degrees of between- and within-cluster imbalance, when the true correlation model is nested exchangeable (NEX). Number of clusters is *I* = 24. The within-period intraclass correlation coefficient (WP-ICC) *α*_0_ is 0.05, and the within-period intraclass correlation coefficient (BP-ICC) *α*_1_ is 0.025. Between cluster imbalance is measured by the coefficient of variation, CV ∈ {0.25, 0.75, 1.25}. The working correlation structure is either NEX or independence (IND).

| Working correlation | $J$ | CV | No within-cluster imbalance | Within-cluster imbalance pattern 1 | Within-cluster imbalance pattern 2 | Within-cluster imbalance pattern 4 |
| --- | --- | --- | --- | --- | --- | --- |
| NEX | 3 | 0.25 | 0.988 (0.985, 0.991) | 0.986 (0.974, 0.996) | 0.964 (0.953, 0.975) | 0.963 (0.951, 0.975) |
|  |  | 0.75 | 0.901 (0.880, 0.919) | 0.888 (0.833, 0.930) | 0.864 (0.816, 0.905) | 0.862 (0.815, 0.905) |
|  |  | 1.25 | 0.762 (0.724, 0.796) | 0.726 (0.648, 0.796) | 0.716 (0.640, 0.786) | 0.703 (0.622, 0.779) |
|  | 5 | 0.25 | 0.988 (0.986, 0.991) | 0.986 (0.977, 0.995) | 0.958 (0.949, 0.966) | 0.959 (0.949, 0.967) |
|  |  | 0.75 | 0.903 (0.882, 0.920) | 0.888 (0.849, 0.923) | 0.865 (0.824, 0.898) | 0.864 (0.820, 0.905) |
|  |  | 1.25 | 0.765 (0.730, 0.798) | 0.725 (0.659, 0.785) | 0.705 (0.639, 0.765) | 0.708 (0.647, 0.773) |
|  | 13 | 0.25 | 0.989 (0.986, 0.991) | 0.985 (0.981, 0.988) | 0.975 (0.971, 0.978) | 0.977 (0.972, 0.980) |
|  |  | 0.75 | 0.905 (0.886, 0.922) | 0.878 (0.851, 0.902) | 0.868 (0.841, 0.889) | 0.872 (0.847, 0.895) |
|  |  | 1.25 | 0.770 (0.735, 0.802) | 0.703 (0.661, 0.744) | 0.697 (0.653, 0.741) | 0.696 (0.655, 0.741) |
| IND | 3 | 0.25 | 0.955 (0.945, 0.964) | 0.958 (0.942, 0.972) | 0.878 (0.863, 0.892) | 0.880 (0.865, 0.896) |
|  |  | 0.75 | 0.721 (0.673, 0.763) | 0.767 (0.696, 0.831) | 0.693 (0.629, 0.756) | 0.700 (0.643, 0.756) |
|  |  | 1.25 | 0.501 (0.440, 0.560) | 0.591 (0.508, 0.659) | 0.532 (0.464, 0.611) | 0.523 (0.453, 0.604) |
|  | 5 | 0.25 | 0.954 (0.937, 0.972) | 0.971 (0.944, 0.994) | 0.899 (0.878, 0.919) | 0.903 (0.881, 0.928) |
|  |  | 0.75 | 0.722 (0.656, 0.776) | 0.818 (0.751, 0.887) | 0.760 (0.696, 0.816) | 0.760 (0.689, 0.824) |
|  |  | 1.25 | 0.502 (0.430, 0.564) | 0.639 (0.554, 0.725) | 0.593 (0.513, 0.671) | 0.593 (0.505, 0.681) |
|  | 13 | 0.25 | 0.953 (0.927, 0.978) | 0.987 (0.973, 1.000) | 0.975 (0.961, 0.988) | 0.973 (0.957, 0.989) |
|  |  | 0.75 | 0.714 (0.641, 0.783) | 0.909 (0.866, 0.945) | 0.891 (0.847, 0.928) | 0.891 (0.851, 0.928) |
|  |  | 1.25 | 0.492 (0.416, 0.573) | 0.778 (0.714, 0.839) | 0.770 (0.694, 0.832) | 0.770 (0.700, 0.825) |

### 5.5 Cluster-period size

Web Figures 13 and 14 present the counterparts to Figure 4 but with the mean cluster-period sizes 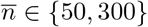 As the mean cluster-period size increases from 50 to 300, the median RE under the NEX working correlation model increase when the WP-ICC is at least 0.05, and decreases when the WP-ICC is 0.01. This pattern is mostly apparent when the degree of between-cluster imbalance is large (CV = 1.25), or there are a large number of clusters (*I* = 96). When the within-cluster imbalance patterns are introduced, we observed similar trends (see Web Figures 15 and 16). Web Figures 17 and 18 present the counterparts to Figure 5 with mean cluster-period sizes 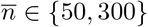 Under the IND working correlation, the median RE simply decreases as the mean cluster-period sizes increases, signaling additional efficiency loss under unequal cluster sizes with a larger total sample size. Findings remain the same when within-cluster imbalance patterns are introduced, as in Web Figure 19 and 20.

### 5.6 Sensitivity to baseline prevalence, intervention effect and secular trend

As a sensitivity analysis with limited scope, we also explored the impact of other model factors on the RE under the NEX and IND working correlation structures. We considered a smaller baseline prevalence {1 + exp(−*β*_1_)} ^−1^ = 0.1, a smaller intervention effect OR, {1 + exp(−*δ*)}^−1^ = 0.75, and an increasing or decreasing secular trend. Specifically, we explore a gently increasing secular trend such that

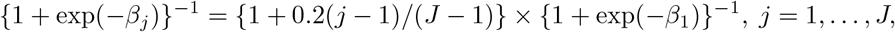

as well as a gently decreasing secular trend such that

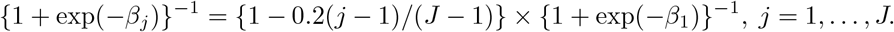

We did not consider a more dramatic secular trend as a recent re-analysis of the Washington State EPT trial suggests minimal secular trend (Li et al., 2021). Web Tables 4 to 11 each present the median and IQR of RE under a factorial combination of 2 levels of baseline prevalence *×* 2 levels of intervention effect *×* 3 different secular trends *×* 3 degrees of between-cluster imbalance (36 cells in total), while holding the number of clusters constant. Relative to the factors described in the preceding texts, the impact of a smaller baseline prevalence, a smaller intervention effect or non-constant secular trend generally have negligible additional impact on the efficiency loss due to unequal cluster sizes, regardless of the specification of working correlation structures.

## 6 Simulation results when the true correlation structure is exponential decay

In Web Appendix D, we present the results on RE when the underlying true correlation structure is ED, in parallel to results elaborated in Section 5. Under the ED correlation structure, we use *ρ* to measure the degree of BP-ICC decay, which plays a similar role to *α*_1_/*α*_0_ under the NEX correlation structure. Generally, we find that all the observed relationships between RE and key design and model factors are similar regardless of whether the true correlation structure is NEX or ED. For example, Web Figure 21 and 26 present highly similar RE-CV relationships to those in Figure 2 and 3. Surprisingly, while the exact value of the asymptotic variances can be quite different when the true correlation structure is NEX versus ED as studied in Kasza et al. (2019) under equal cluster sizes and with a continuous outcome, the impact of unequal cluster sizes measured by the median RE can be highly similar across the two true correlation models in our evaluations with binary outcomes.

## 7 A Monte Carlo procedure for sample size calculation

The RE of the treatment effect estimator has important implications for designing SW-CRTs. In particular, our simulation procedure suggests a Monte Carlo power calculation procedure for SW-CRTs with unequal cluster sizes and binary outcomes. Here we present this procedure as an extension to the sample size method developed in Li et al. (2018b), which assumes equal cluster-period sizes.

Suppose we are interesting in testing the null *H*_0_ : *δ* = 0 versus the alternative *H*_1_ : *δ* = Δ for some target effect size with odds ratio, exp(Δ). Conditional on a specific design **Ω**, for a prescribed type I error rate *ϵ*_1_ and type II error rate *ϵ*_2_, the required number of clusters based on a *t*-test to achieve 100(1 − *ϵ*_2_)% power satisfies the following generic inequality

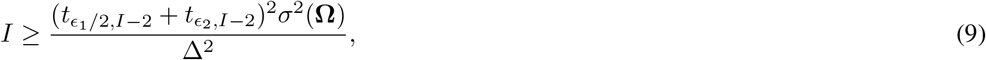

where *t*_*ϵ,I*−2_ is the *ϵ*-quantile of the *t*-distribution with *I* − 2 degree of freedom, and 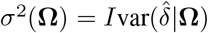 is the scaled variance of the intervention effect estimator. While other choices of the degrees of freedom are possible, we focus on the *I* − 2 degrees of freedom because a number of previous simulation studies indicated adequate control of test size with a small number of clusters (Li, 2020; Ford and Westgate, 2020; Li et al., 2021). Given Δ, the required number of clusters *I* is the smallest number such that (9) holds. Equivalently, (9) can be represented based on the minumum detectable effect size,

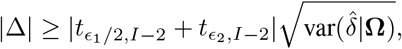

where 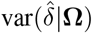 is implicitly a function of *I*. Therefore, sample size determination boils down to the determination of *σ*^2^(**Ω**) or 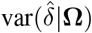, which can be a complicated nonlinear function of design and model parameters. Because the scalar expression of 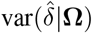 is generally difficult to obtain with binary outcomes and unequal cluster sizes, we propose the following Monte Carlo approach to compute the required sample size for cross-sectional SW-CRTs:

1. Given an initial choice of number of cluster *I*_0_, number of periods *J*, specify the treatment-by-period diagram with the desired number of treatment sequences, and number of clusters per sequence. For the cases that *J* − 1 is not a divisor of *I*_0_, (i.e, the clusters cannot be evenly distributed across the treatment sequences), one can decide a priori the steps with more clusters crossing over.
2. Given the model parameters including the baseline prevalence, anticipated secular trend, intervention effect, obtain the prevalence of outcomes for each cluster (per sequence) during each period according to the marginal mean model, i.e, *µ*_*ij*_ = *g*^−1^(*β*_*j*_ + *X*_*ij*_*δ*).
3. Specify the degree of between-cluster imbalance and/or within-cluster imbalance (for example, following the strategies in Section 4), and simulate the cluster-period sizes *n*_*ij*_ for all *I × J* cluster-periods such that the mean cluster-period size equals to 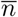. Each simulation replicate corresponds to a possible design with unequal cluster sizes, **Ω**_unequal_. Repeat this steps for *R* times, and record each design as 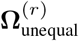 for *r* = 1, …, *R*.
4. Given the assumptions on the ICC parameters, and each simulated design 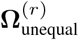, numerically compute 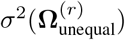 Based on Theorem 2.1, 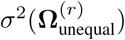 is the (*J* + 1, *J* + 1)-th element of the sandwich variance matrix 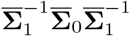 when independence working correlation structure is used, and is the (*J* + 1, *J* + 1)-th element of the sandwich variance matrix of 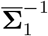 when the correct NEX or ED working correlation structure is used.
5. Obtain the mean variance as 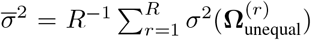 and plug it into the Equation (9). Check to see if *I*_0_ satisfies the inequality. If so, then *I*_0_ clusters already provides adequate power, and one can try to see whether a smaller *I*_1_ *< I*_0_ satisfies the inequality and further reduce the sample size. If the inequality fails to hold with *I*_0_, set 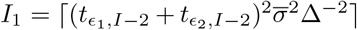 and repeat the above steps. This iterative process is repeated until the smallest *I* is identified to satisfy the inequality in Equation (9).

In principle, the above Monte Carlo procedure is iterative as the variance of treatment effect estimator depends on the current number of clusters, and one needs to search for the smallest number of clusters to provide adequate power. However, because the variance under a specific design 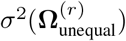 is computed based on the cluster-period mean model according to Theorem 2.1, we only need to invert *J × J* matrices and the computational burden can be dramatically reduced, as evidenced by the feasibility of our simulation study. Additionally, the computational efficiency of the sample size procedure also depends on appropriate choice of an initial value *I*_0_. For example, one could first assume equal cluster sizes and use the existing sample size procedure in Li et al. (2018b) to obtain the required number of clusters *I*_equal_, and then set *I*_0_ = *I*_equal_ to reduce the iterations needed for convergence. We provide an illustrative sample size calculation using this approach below.

### 7.1 Application to the Washington State EPT trial

As shown in Figure 1, the Washington State EPT trial randomized 22 LHJs over 4 steps and 5 periods. This is a cross-sectional design and the primary outcome was Chlamydia test positivity among women tested in sentinel clinics (Golden et al., 2015). Based on the cluster-period sizes, the CV of mean cluster-period sizes within each cluster can be computed as 0.99, which is considerable even in the range of CV tried in the simulation analysis. Given the set of design and model parameters, we aim to compute the required number of clusters *I* such that the trial has 80% power. We assume the mean cluster-period size is 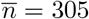, as informed by Figure 1. In the absence of intervention, we assume the marginal prevalence of chlamydia positivity is approximately 7.6% at baseline and no secular trend. This is concordant with the empirical reanalysis of the Washington State EPT study in Li et al. (2021), which suggested minimum secular trend for this outcome. To further illustrate potential differences due to assumptions on ICC, we consider the NEX and ED true correlation structures discussed in Table 1 as well as a simple exchangeable correlation model with equal WP-ICC and BP-ICC. For all three correlation structures, we assume the WP-ICC, *α*_0_ = 0.007. Under the NEX and ED correlation structures, we assume *α*_1_ = *α*_0_/2 = 0.0035 and *ρ* = 0.7. These values are informed by the analysis results in Li et al. (2021). Similar to Golden et al. (2015), assuming a 0.05 type I error rate and target effect size in OR of exp(Δ) = 0.7, we estimate the required number of LHJs to achieve at least 80% power. When the estimated *I* is not divisible by 4, we try to have balanced an allocation as possible, but prioritize the first and last steps over the middle steps as suggested in Lawrie et al. (2015) and Li et al. (2018a) for efficiency considerations. To implement our sample size procedure, we choose the initial value *I*_0_ using the equal cluster size method in Li et al. (2018b). We consider four levels of between-cluster imbalance measured by CV ∈ {0, 0.25, 0.75, 1.25}, as well as three levels of within-cluster imbalance as introduced in our simulation design. The estimated sample size assuming a correctly specified working correlation and its counterpart assuming an independence working correlation (in parentheses) are presented in Table 3. Of note, the Monte Carlo procedure converged in seconds.

**Table 3.** Estimated number of clusters for the Washington State Expedited Partner Therapy trial as a function of between-cluster imbalance measured by coefficient of variation (CV) and three different patterns of within-cluster imbalance, when the true correlation structure is exchangeable, nested exchangeable or exponential decay. The first number in each cell is the sample size estimate under correctly specified correlation structure, while the number in the parenthesis corresponds to the sample size estimate assuming working independence.

| True correlation structure | CV | No within-cluster imbalance | Within-cluster imbalance pattern 2 | Within-cluster imbalance pattern 4 |
| --- | --- | --- | --- | --- |
| Exchangeable | 0 | 11 (31) | 11 (32) | 11 (33) |
|  | 0.25 | 11 (33) | 12 (33) | 12 (33) |
|  | 0.75 | 12 (43) | 13 (38) | 13 (38) |
|  | 1.25 | 13 (64) | 17 (48) | 17 (48) |
| Nested exchangeable | 0 | 18 (25) | 19 (26) | 19 (27) |
|  | 0.25 | 18 (26) | 19 (27) | 19 (27) |
|  | 0.75 | 20 (34) | 21 (32) | 21 (32) |
|  | 1.25 | 24 (50) | 26 (42) | 26 (42) |
| Exponential decay | 0 | 17 (27) | 18 (28) | 18 (29) |
|  | 0.25 | 18 (28) | 18 (29) | 18 (29) |
|  | 0.75 | 19 (37) | 21 (34) | 21 (34) |
|  | 1.25 | 22 (54) | 26 (43) | 26 (43) |

Under the correctly specified working correlation structure, the sample size estimates are reasonably insensitive to between-cluster imbalance as long as the CV does not exceed 0.75. The patterns for within-cluster imbalance also have negligible impact on the sample size. Among different working correlation structures, the exchangeable working structure corresponds to the smallest sample size, which is expected because Li et al. (2018b) showed that larger BP-ICC generally increases study power. In contrast, if the independence working correlation is considered, the sample size estimates can be substantially larger than their counterparts obtained under the correct working correlation structure. This is especially true when the BP-ICC equals to the WP-ICC. In fact, even with equal cluster-period sizes, the sample size obtained under the independence working correlation structure is at least three-fold of that obtained under the correctly-specified exchangeable working correlation structure. The sample size estimates under working independence also rapidly inflate with larger between-cluster imbalance, and appear less robust to unequal cluster sizes compared to those obtained under the correct correlation model. In the presence of within-cluster imbalance or when the BP-ICC deviates from the WP-ICC, the sample size estimates based on independence GEE become smaller. Overall, assuming a correct working correlation model, a maximum of 17 LHJs (corresponding to the most extreme imbalance scenario) are needed to ensure 80% power under the working exchangeable structure. If the true correlation structure is nested exchangeable or exponential decay, a maximum of 26 LHJs are needed. To conclude, modeling the correlation structure in this trial protects against dramatic efficiency loss due to unequal cluster sizes, and leads to much smaller sample sizes compared to assuming working independence.

## 8 Discussion

In this article, we investigate the RE of the GEE treatment effect estimator under unequal versus equal cluster sizes in SW-CRTs with binary outcomes. Because all prior studies assumed a linear mixed model with continuous outcomes, our results complement existing knowledge by exploring the properties and caveats for marginal analysis of SW-CRTs with binary outcomes. We assume a cross-sectional design as in the Washington State EPT study, and consider two popular multilevel correlation structures: the nested exchangeable and exponential decay structures. Both correlation structures include the simple exchangeable structure (e.g. as implied by the Hussey and Hughes (2007) model) as a special case, but are considered more realistic (Taljaard et al., 2016).

The main message of our simulation findings can be summarized as follows. First, the GEE analysis with the correct working correlation structure is much less prone to efficiency loss under between-cluster imbalance compared to the independence GEE, whose RE sharply decreases with a larger degree of between-cluster imbalance. Second, the RE of GEE analysis with the true working correlation structure critically depends on the magnitude of the WP-ICC as well as the amount of BP-ICC decay from WP-ICC. In particular, when BP-ICC equals WP-ICC, the efficiency loss due to between-cluster imbalance is minimal but can be larger when within-cluster imbalance is introduced. However, the efficiency loss due to between-cluster imbalance becomes more notable once BP-ICC deviates from WP-ICC. The statement in Kristunas et al. (2017) that “(between-cluster) imbalance in cluster size was not found to have a notable effect on the power of SW-CRTs” belies the dependence of RE on ICC as they assumed the random intercept linear mixed model Hussey and Hughes (2007), where BP-ICC equals WP-ICC. Third, the RE of the independence GEE estimator is particularly sensitive to values of WP-ICC. Values of the BP-ICC does not substantially affect the RE of the independence GEE, except with the introduction of within-cluster imbalance. This finding is expected given our analytical result in Theorem 3.1 for three-period designs. More intuitively, the independence GEE analysis heavily depends on between-cluster comparisons (or “vertical analysis” as defined in Matthews and Forbes (2017)) instead of within-cluster comparisons (or “horizontal analysis”), and therefore its variability is more dependent on the WP-ICC than the BP-ICC. Fourth, whereas a larger SW-CRT with more clusters and larger mean cluster-period sizes generally has a small effect on the RE of GEE analysis when the working correlation structure is correctly specified, a larger SW-CRT is associated with greater efficiency loss due to between-cluster imbalance for independence GEE. Finally, while the number of periods has minimum effect on the RE of the GEE analysis when the working correlation structure is correctly specified, the RE of the independence GEE estimator increases with a larger number of periods as long as there is within-cluster imbalance. We checked to confirm that introducing within-cluster imbalance in our simulation configuration can somewhat reduce the between-cluster imbalance within each period (even though maintaining the overall between-cluster imbalance on mean cluster-period sizes), and therefore improves the stability of vertical analysis which is the dominating component of the independence GEE.

While we have mainly focused on the RE of GEE analysis under unequal versus equal cluster sizes, there remains interest in understanding the RE of GEE analysis under the true versus independence working correlation model in SW-CRTs with binary outcomes. This is defined by

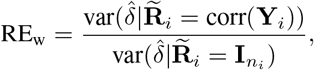

which is strictly below one unless all ICC parameters are zero. In the context of the Washington State EPT study, Table 3 implies that RE_w_ can substantially deviate from one even with a small WP-ICC. To provide a more complete perspective, in Web Appendix E, we present RE_w_ as a function of ICC parameters under equal cluster-period sizes. Evidently, RE_w_ is a monotonically decreasing function of both WP-ICC and BP-ICC, and the efficiency loss due to the incorrect working independence assumption becomes maximum when WP-ICC equals BP-ICC. These findings on RE_w_ re-interpret those of Mancl and Leroux (1996); Wang and Carey (2003) in the context of SW-CRTs, and confirm the notable efficiency loss under the independence GEE for estimating the regression coefficient of a covariate that varies within clusters. This is in contrast to parallel CRTs, where there can be no efficiency loss for independence GEE with equal cluster sizes (Pan, 2001; Li and Tong, 2021a,b). From the efficiency perspective, our results favor the GEE analysis coupled with an appropriate working multilevel correlation structure in SW-CRTs, possibly through the matrix-adjusted estimating equations (MAEE) approach developed in Preisser et al. (2008), and Li et al. (2018b, 2019, 2021). The MAEE approach has been validated in previous simulations with SW-CRTs, and is recently implemented in the `geeCRT` R package. Relatedly, reporting ICC estimates is also recommended practice in SW-CRTs per the CONSORT extension to SW-CRTs (Hemming et al., 2018), as those values provide evidence for designing future trials with a similar endpoints.

To assist with sample size determination in SW-CRTs with unequal cluster sizes, we further developed a Monte Carlo search algorithm in Section 7. This approach is computationally efficient since it only requires numerical inversion of *J* × *J* matrices regardless of the actual cluster-period sizes (Theorem 2.1). Alternatively, for sample size calculation, Girling (2018) suggested (1 + CV^2^) as a conservative variance inflation factor due to between-cluster imbalance in SW-CRTs. In our simulations with binary outcomes, we find in Web Appendix F that (1 + CV^2^)^−1^ can be substantially smaller than the median RE under a correct working correlation model, and therefore the variance inflation factor remains highly conservative. However, when the independence working assumption is adopted, (1 + CV^2^)^−1^ is still lower than but much closer to the median RE curve, regardless of the ICC parameters. This comparison confirms that (1+CV^2^) may be a crude upper bound for variance inflation due to unequal cluster sizes for GEE analysis of SW-CRTs with binary outcomes, and underscores the utility of our sample size search algorithm for more accurate sample size determination.

There are a few limitations of our study. First, while our evaluations assumed that the independence working correlation structure is misspecified in SW-CRTs, we have not investigated the efficiency implications when the exchangeable, nested exchangeable or exponential decay correlation structure deviates from the underlying true correlation structure. Asymptotic efficiency evaluation under misspecified non-identity correlation structure is generally challenging because the probability limits of the misspecified ICC estimators are not easy to identify analytically. Therefore, additional simulation studies are required to address this more complex question, possibly by summarizing the empirical variance of the GEE estimator under alternative data generating processes. Second, we have restricted consideration to cross-sectional designs, as motivated by the Washington State EPT trial. On the other hand, unequal cluster sizes can also arise in closed-cohort or open-cohort SW-CRTs (Copas et al., 2015). These alternative designs require slightly different formulations of the within-cluster correlation structures due to repeated outcome measurements for the same subject (Li et al., 2018b; Kasza et al., 2020; Li et al., 2020). It remains to be explored whether the current findings are generalizable to cohort SW-CRTs. Finally, our simulation design parameters are not exhaustive. However, our comprehensive evaluation identified important factors that affect the efficiency patterns of the GEE estimators in cross-sectional SW-CRTs. We have also articulated the critical need to account for correlations in SW-CRTs from an efficiency perspective, providing a rigorous justification for estimating and reporting ICCs, as recommended by the CONSORT extension to SW-CRTs (Hemming et al., 2018).

## Data Availability

The study focuses on statistical methodology and does not involve any analysis of primary data. R code to replicate the simulations can be found in the Data Availability Links.

https://github.com/Zebedial/SWD-variable-cluster-sizes

## Acknowledgements

Research in this article was partially funded by a Patient-Centered Outcomes Research Institute^®^(PCORI^®^ Award ME-2019C1-16196) as well as by CTSA Grant Number UL1 TR000142 from the National Center for Advancing Translational Science (NCATS), a component of the National Institutes of Health (NIH). The statements presented in this article are solely the responsibility of the authors and do not necessarily represent the views of PCORI^®^, its Board of Governors or Methodology Committee, or the National Institutes of Health. Dr. Preisser has received a stipend for service as a merit reviewer from PCORI^®^. Dr. Preisser did not serve on the Merit Review panel that reviewed his project.

