## Supplementary Materials for "Impact of unequal cluster sizes for GEE analyses of stepped wedge cluster randomized trials with binary outcomes"

### Web Appendix A. Analytical inverse for nested exchangeable correlation structure under unequal cluster-period sizes

As noted in Section 2 of the main article, the nested exchangeable correlation structure characterizing the $i$-th cluster over $J$ periods can be expressed by

$$\mathbf{R}_i(\boldsymbol{\alpha}_{\text{NEX}}) = (1-\alpha_0)\mathbf{I}_{n_i} + (\alpha_0-\alpha_1)\oplus_{j=1}^{J}\mathbf{J}_{n_{ij}} + \alpha_1\mathbf{J}_{n_i},$$

where $\boldsymbol{\alpha}_{\text{NEX}} = (\alpha_0,\alpha_1)^T$, $\alpha_0$ is the within-period ICC, $\alpha_1$ is the between-period ICC, $\mathbf{I}_s$ is the $s\times s$ identity matrix, $\mathbf{J}_s$ is the $s\times s$ matrix of ones, $n_i=\sum_{j=1}^{J} n_{ij}$ is the $i$-th cluster size, and '$\oplus$' is the block diagonal operator. We provide a derivation of the closed-form inverse of $\mathbf{R}_i$, extending the analytical results in Li et al. (2019) for $J=2$. Let $\mathbf{A}=(1-\alpha_0)\mathbf{I}_{n_i}+(\alpha_0-\alpha_1)\oplus_{j=1}^{J}\mathbf{J}_{n_{ij}}$ and $\mathbf{B}=\alpha_1\mathbf{J}_{n_i}$, by Henderson and Searle (1981), the inverse is

$$\mathbf{R}_i^{-1} = (\mathbf{A}+\mathbf{B})^{-1} = \mathbf{A}^{-1} - \mathbf{A}^{-1}\mathbf{B}(\mathbf{I}_{n_i}+\mathbf{A}^{-1}\mathbf{B})\mathbf{A}^{-1}.$$

It is easy to verify that the $\mathbf{A}^{-1}$ has similar basis matrices as $\mathbf{A}$ and can be expanded as $\mathbf{A}^{-1}=x\mathbf{I}_{n_i}+\oplus_{j=1}^{J}y_j\mathbf{J}_{n_{ij}}$. Since

$$\mathbf{I}_{n_i}=\mathbf{A}\mathbf{A}^{-1}=(1-\alpha_0)x\mathbf{I}_{n_i}+(\alpha_0-\alpha_1)x\oplus_{j=1}^{J}\mathbf{J}_{n_{ij}}+(1-\alpha_0)\oplus_{j=1}^{J}y_j\mathbf{J}_{n_{ij}}+(\alpha_0-\alpha_1)\oplus_{j=1}^{J}y_j n_{ij}\mathbf{J}_{n_{ij}},$$

we must have

$$x=\frac{1}{1-\alpha_0}\quad y_j=-\frac{\alpha_0-\alpha_1}{(1-\alpha_0)\psi_j},$$

where $\psi_j = 1+(n_{ij}-1)\alpha_0-n_{ij}\alpha_1$. Since

$$\mathbf{A}^{-1}=\frac{1}{1-\alpha_0}\mathbf{I}_{n_i}-\frac{\alpha_0-\alpha_1}{(1-\alpha_0)}\oplus_{j=1}^{J}\frac{1}{\psi_j}\mathbf{J}_{n_{ij}},$$

$$\mathbf{A}^{-1}\mathbf{B} = \frac{\alpha_1}{1-\alpha_0}\mathbf{J}_{n_i} - \frac{(\alpha_0-\alpha_1)\alpha_1}{(1-\alpha_0)}\Big(\oplus_{j=1}^{J}\frac{n_{ij}}{\psi_j}\mathbf{I}_{n_{ij}}\Big)\mathbf{J}_{ni} = \Big(\oplus_{j=1}^{J}\frac{\alpha_1}{\psi_j}\mathbf{I}_{n_{ij}}\Big)\mathbf{J}_{n_i}.$$

Observe that

$$\mathbf{I}_{n_i} + \mathbf{A}^{-1}\mathbf{B} = \mathbf{I}_{n_i} + \Big(\oplus_{j=1}^{J}\frac{\alpha_1}{\psi_j}\mathbf{I}_{n_{ij}}\Big)\mathbf{J}_{n_i},$$

whose inverse shares the same basis matrices, and are denoted by $(\mathbf{I}_{n_i}+\mathbf{A}^{-1}\mathbf{B})^{-1} = \mathbf{I}_{n_i}+[\oplus_{j=1}^{J}z_j\mathbf{I}_{n_{ij}}]\mathbf{J}_{n_i}$. Observe that

$$\begin{aligned}\mathbf{I}_{n_i} &= (\mathbf{I}_{n_i}+\mathbf{A}^{-1}\mathbf{B})(\mathbf{I}_{n_i}+\mathbf{A}^{-1}\mathbf{B})^{-1}\\ &= \mathbf{I}_{n_i} + \Big(\oplus_{j=1}^{J} z_j\mathbf{I}_{n_{ij}}\Big)\mathbf{J}_{n_i} + \alpha_1\Big(\oplus_{j=1}^{J}\frac{1}{\psi_j}\mathbf{I}_{n_{ij}}\Big)\mathbf{J}_{n_i} + \alpha_1\Big(\oplus_{j=1}^{J}\frac{1}{\psi_j}\mathbf{I}_{n_{ij}}\Big)\mathbf{J}_{n_i}\Big(\oplus_{j=1}^{J} z_j\mathbf{I}_{n_{ij}}\Big)\mathbf{J}_{n_i}\\ &= \mathbf{I}_{n_i}+\mathbf{C}.\end{aligned}$$

The $(j,j)$-th block of $\mathbf{C}$ is

$$0 = z_j\mathbf{J}_{n_{ij}} + \frac{\alpha_1}{\psi_j}\mathbf{J}_{n_{ij}} + \frac{\alpha_1}{\psi_j}\Big(\sum_{s=1}^{J} n_{is}z_s\Big)\mathbf{J}_{n_{ij}},$$

which implies

$$z_j + \frac{\alpha_1}{\psi_j} + \frac{\alpha_1}{\psi_j}\Big(\sum_{s=1}^{J} n_{is}z_s\Big) = 0.$$

Although the above condition is derived solely based on the diagonal block information, it turns out to be sufficient to ensure that $\mathbf{C}=0$, and we can solve for $z_j$ by noting that

$$\sum_{j=1}^{J} n_{ij}z_j + \sum_{j=1}^{J}\frac{n_{ij}\alpha_1}{\psi_j} + \Big(\sum_{j=1}^{J}\frac{n_{ij}\alpha_1}{\psi_j}\Big)\Big(\sum_{s=1}^{J} n_{is}z_s\Big) = 0,$$

and

$$\sum_{j=1}^{J} n_{ij}z_j = -\sum_{j=1}^{J}\frac{n_{ij}\alpha_1}{\psi_j}\Big/\Big(1+\sum_{j=1}^{J}\frac{n_{ij}\alpha_1}{\psi_j}\Big).$$

Define

$$\gamma_j = \psi_j\Big(1+\sum_{j=1}^{J}\frac{n_{ij}\alpha_1}{\psi_j}\Big),$$

we have,

$$z_j = -\frac{\alpha_1}{\psi_j}\Big(1+\sum_{j=1}^{J} n_{ij}z_j\Big) = -\frac{\alpha_1}{\psi_j}\Big/\Big(1+\sum_{j=1}^{J}\frac{n_{ij}\alpha_1}{\psi_j}\Big) = -\frac{\alpha_1}{\gamma_j}.$$

Then

$$\begin{aligned}(\mathbf{I}_{n_i}+\mathbf{A}^{-1}\mathbf{B})^{-1}\mathbf{A}^{-1} &= \Big[\mathbf{I}_{n_i} - \Big(\oplus_{j=1}^{J}\frac{\alpha_1}{\gamma_j}\mathbf{I}_{n_{ij}}\Big)\mathbf{J}_{n_i}\Big]\Big[\frac{1}{1-\alpha_0}\mathbf{I}_{n_i} - \oplus_{j=1}^{J}\frac{\alpha_0-\alpha_1}{(1-\alpha_0)\psi_j}\mathbf{J}_{n_{ij}}\Big]\\ &= \frac{1}{1-\alpha_0}\mathbf{I}_{n_i} - \Big(\oplus_{j=1}^{J}\frac{\alpha_1}{(1-\alpha_0)\gamma_j}\mathbf{I}_{n_{ij}}\Big)\mathbf{J}_{n_i} - \oplus_{j=1}^{J}\frac{\alpha_0-\alpha_1}{(1-\alpha_0)\psi_j}\mathbf{J}_{n_{ij}}\\ &\quad +\Big(\oplus_{j=1}^{J}\frac{\alpha_1}{(1-\alpha_0)\gamma_j}\mathbf{I}_{n_{ij}}\Big)\mathbf{J}_{n_i}\Big(\oplus_{j=1}^{J}\frac{n_{ij}(\alpha_0-\alpha_1)}{\psi_j}\mathbf{I}_{n_{ij}}\Big)\\ &= \frac{1}{1-\alpha_0}\mathbf{I}_{n_i} - \oplus_{j=1}^{J}\frac{\alpha_0-\alpha_1}{(1-\alpha_0)\psi_j}\mathbf{J}_{n_{ij}} - \Big(\oplus_{j=1}^{J}\frac{\alpha_1}{\gamma_j}\mathbf{I}_{n_{ij}}\Big)\mathbf{J}_{n_i}\Big(\oplus_{j=1}^{J}\frac{1}{\psi_j}\mathbf{I}_{n_{ij}}\Big).\end{aligned}$$

Further, routine calculations show that

$$\begin{aligned}
\mathbf{A}^{-1}\mathbf{B}(\mathbf{I}_{n_i}+\mathbf{A}^{-1}\mathbf{B})^{-1}\mathbf{A}^{-1} &= \left(\oplus_{j=1}^{J}\frac{\alpha_1}{(1-\alpha_0)\psi_j}\mathbf{I}_{n_{ij}}\right)\mathbf{J}_{n_i} - \left(\oplus_{j=1}^{J}\frac{\alpha_1}{\psi_j}\mathbf{I}_{n_{ij}}\right)\mathbf{J}_{n_i}\left(\oplus_{j=1}^{J}\frac{n_{ij}(\alpha_0-\alpha_1)}{(1-\alpha_0)\psi_j}\mathbf{I}_{n_{ij}}\right) \\
&\quad -\left(\oplus_{j=1}^{J}\frac{\alpha_1}{\psi_j}\mathbf{I}_{n_{ij}}\right)\mathbf{J}_{n_i}\left(\oplus_{j=1}^{J}\frac{\alpha_1}{\gamma_j}\mathbf{I}_{n_{ij}}\right)\mathbf{J}_{n_i}\left(\oplus_{j=1}^{J}\frac{1}{\psi_j}\mathbf{I}_{n_{ij}}\right) \\
&= \left(\oplus_{j=1}^{J}\frac{\alpha_1}{(1-\alpha_0)\psi_j}\mathbf{I}_{n_{ij}}\right)\mathbf{J}_{n_i} - \left(\oplus_{j=1}^{J}\frac{\alpha_1}{\psi_j}\mathbf{I}_{n_{ij}}\right)\mathbf{J}_{n_i}\left(\oplus_{j=1}^{J}\frac{n_{ij}(\alpha_0-\alpha_1)}{(1-\alpha_0)\psi_j}\mathbf{I}_{n_{ij}}\right) \\
&\quad -\left(\oplus_{j=1}^{J}\frac{\alpha_1}{\psi_j}\mathbf{I}_{n_{ij}}\right)\mathbf{J}_{n_i}\left(\sum_{s=1}^{J}\frac{n_{is}\alpha_1}{\gamma_s}\right)\left(\oplus_{j=1}^{J}\frac{1}{\psi_j}\mathbf{I}_{n_{ij}}\right) \\
&= \left(\oplus_{j=1}^{J}\frac{\alpha_1}{\psi_j}\mathbf{I}_{n_{ij}}\right)\mathbf{J}_{n_i}\left(\oplus_{j=1}^{J}\frac{1-\alpha_1\sum_{s=1}^{J}n_{is}/\gamma_s}{\psi_j}\mathbf{I}_{n_{ij}}\right).
\end{aligned}$$

The inverse is then given in closed form by

$$\mathbf{R}_i^{-1} = \frac{1}{1-\alpha_0}\mathbf{I}_{n_i} - \frac{\alpha_0-\alpha_1}{(1-\alpha_0)}\oplus_{j=1}^{J}\frac{1}{\psi_j}\mathbf{J}_{n_{ij}} - \left(\oplus_{j=1}^{J}\frac{\alpha_1}{\psi_j}\mathbf{I}_{n_{ij}}\right)\mathbf{J}_{n_i}\left(\oplus_{j=1}^{J}\frac{1-\alpha_1\sum_{s=1}^{J}n_{is}/\gamma_s}{\psi_j}\mathbf{I}_{n_{ij}}\right). \quad (1)$$

Li et al. (2019) derived a special case of (1) with $J = 2$ periods, and the above expression is more general and for any positive integer $J$. Furthermore, when $n_{ij} = n_{is} = m$ for all $j \neq s$, we have $\psi_j = \psi = 1 + (m-1)\alpha_0 - m\alpha_1$, and $\gamma_j = \gamma = \psi + Jm\alpha_1 = 1 + (m-1)\alpha_0 + m(J-1)\alpha_1$, which are two eigenvalues of the nested exchangeable correlation matrix with equal cluster-period sizes (Li et al., 2018). In this case, the inverse reduces to the formula presented in Teerenstra et al. (2010).

#### Web Appendix B. Proof of theorem 3.1

In this section, Theorem 3.1 in the main article will be proved. We first consider the case that the true correlation structure is nested exchangeable. The outcomes of interest can be continuous, binary, or count outcomes, which implies the canonical link function in the marginal model (2).

$$g(\mu_{ijk}) = \beta_j + X_{ij}\delta \quad (2)$$

Assume there are $I$ clusters being involved in a cross-sectional stepped wedge trial with 3 periods. $I_1$ clusters are randomized to the first treatment sequence where the clusters receive treatment in the second and the third period, and $I_2$ clusters are randomized to the second treatment sequence where the clusters would receive treatment only in the third period.

Theorem 2.1 in the main article shows the sandwich variance estimator of the parameter set $\boldsymbol{\theta} = (\beta_1, \beta_2, \beta_3, \delta)^T$ has the form

$$\overline{\boldsymbol{\Sigma}}_1^{-1}\overline{\boldsymbol{\Sigma}}_0\overline{\boldsymbol{\Sigma}}_1^{-1} = \left\{\sum_{i=1}^{I}\overline{\mathbf{D}}_i^T\widetilde{\mathbf{V}}_i^{-1}\overline{\mathbf{D}}_i\right\}^{-1}\overline{\boldsymbol{\Sigma}}_0\left\{\sum_{i=1}^{I}\overline{\mathbf{D}}_i^T\widetilde{\mathbf{V}}_i^{-1}\overline{\mathbf{D}}_i\right\}^{-1}$$

where $\overline{\mathbf{D}}_i = \partial\overline{\boldsymbol{\mu}}_i/\partial\boldsymbol{\theta}^T$, $\widetilde{\mathbf{V}}_i$ is the working covariance matrix for the cluster-period means, and

$$\overline{\boldsymbol{\Sigma}}_0 = \sum_{i=1}^{I}\overline{\mathbf{D}}_i^T\widetilde{\mathbf{V}}_i^{-1}\text{cov}(\overline{\mathbf{Y}}_i)\widetilde{\mathbf{V}}_i^{-1}\overline{\mathbf{D}}_i$$

. When the working correlation is independence,

$$\widetilde{\mathbf{V}}_i = \begin{pmatrix} \text{var}(\overline{Y}_{i1}) & 0 & 0 \\ 0 & \text{var}(\overline{Y}_{i2}) & 0 \\ 0 & 0 & \text{var}(\overline{Y}_{i3}) \end{pmatrix} = \begin{pmatrix} \frac{\phi\nu_{i1}}{n_{i1}} & 0 & 0 \\ 0 & \frac{\phi\nu_{i2}}{n_{i2}} & 0 \\ 0 & 0 & \frac{\phi\nu_{i3}}{n_{i3}} \end{pmatrix}$$

where for $j = 1, 2, 3$, $n_{ij}$ is the cluster-period sizes for the $(i, j)$-th cluster, $\phi$ is the dispersion parameter, and $\nu_{ij}$ is the variance function related to the mean for the $(i, j)$-th cluster. For continuous outcomes, $\nu_{ij} = 1$. For binary outcomes, $\nu_{ij} = \mu_{ij}(1 - \mu_{ij})$. For count outcomes, $\nu_{ij} = \mu_{ij}$. We can also write out the inverse of the working covariance matrix as

$$\widetilde{\mathbf{V}}_i^{-1} = \begin{pmatrix} \frac{n_{i1}}{\phi\nu_{i1}} & 0 & 0 \\ 0 & \frac{n_{i2}}{\phi\nu_{i2}} & 0 \\ 0 & 0 & \frac{n_{i3}}{\phi\nu_{i3}} \end{pmatrix}.$$

Besides, the true covariance structure for the cluster-period means is

$$\text{cov}(\overline{\mathbf{Y}}_i) = \begin{pmatrix} \frac{\phi\nu_{i1}}{n_{i1}}[1 + (n_{i1} - 1)\alpha_0] & \sqrt{\nu_{i1}\nu_{i2}}\phi\alpha_1 & \sqrt{\nu_{i1}\nu_{i3}}\phi\alpha_1 \\ \sqrt{\nu_{i1}\nu_{i2}}\phi\alpha_1 & \frac{\phi\nu_{i2}}{n_{i2}}[1 + (n_{i2} - 1)\alpha_0] & \sqrt{\nu_{i2}\nu_{i3}}\phi\alpha_1 \\ \sqrt{\nu_{i1}\nu_{i3}}\phi\alpha_1 & \sqrt{\nu_{i2}\nu_{i3}}\phi\alpha_1 & \frac{\phi\nu_{i3}}{n_{i3}}[1 + (n_{i3} - 1)\alpha_0] \end{pmatrix}.$$

For $i = 1, 2, ..., I_1$, we then have

$$\overline{\mathbf{D}}_i = \begin{pmatrix} \nu_{i1} & 0 & 0 & 0 \\ 0 & \nu_{i2} & 0 & \nu_{i2} \\ 0 & 0 & \nu_{i3} & \nu_{i3} \end{pmatrix}$$

Alternatively, for $i = I_1 + 1, I_1 + 2, ..., I_1 + I_2$,

$$\overline{\mathbf{D}}_i = \begin{pmatrix} \nu_{i1} & 0 & 0 & 0 \\ 0 & \nu_{i2} & 0 & 0 \\ 0 & 0 & \nu_{i3} & \nu_{i3} \end{pmatrix}$$

Thus, the matrix expression of $\overline{\mathbf{D}}_i^T \widetilde{\mathbf{V}}_i^{-1} \overline{\mathbf{D}}_i$ for cluster $i$ is either

$$\begin{pmatrix} n_{i1}\nu_{i1} & 0 & 0 & 0 \\ 0 & n_{i2}\nu_{i2} & 0 & n_{i2}\nu_{i2} \\ 0 & 0 & n_{i3}\nu_{i3} & n_{i3}\nu_{i3} \\ 0 & n_{i2}\nu_{i2} & n_{i3}\nu_{i3} & n_{i2}\nu_{i2} + n_{i3}\nu_{i3} \end{pmatrix}$$

or

$$\begin{pmatrix} n_{i1}\nu_{i1} & 0 & 0 & 0 \\ 0 & n_{i2}\nu_{i2} & 0 & 0 \\ 0 & 0 & n_{i3}\nu_{i3} & n_{i3}\nu_{i3} \\ 0 & 0 & n_{i3}\nu_{i3} & n_{i3}\nu_{i3} \end{pmatrix}$$

determined by the step in which the cluster is randomized.

Similarly, there are two forms of $\overline{\mathbf{D}}_i^T \widetilde{\mathbf{V}}_i^{-1} \text{cov}(\overline{\mathbf{Y}}_i) \widetilde{\mathbf{V}}_i^{-1} \overline{\mathbf{D}}_i$, depending on the value of $i$, namely,

$$\frac{1}{\phi} \begin{pmatrix} \nu_{i1}n_{i1}[1 + (n_{i1} - 1)\alpha_0] & \sqrt{\nu_{i1}\nu_{i2}}n_{i1}n_{i2}\alpha_1 & \sqrt{\nu_{i1}\nu_{i3}}n_{i1}n_{i3}\alpha_1 & A_1 \\ \sqrt{\nu_{i1}\nu_{i2}}n_{i1}n_{i2}\alpha_1 & \nu_{i2}n_{i2}[1 + (n_{i2} - 1)\alpha_0] & \sqrt{\nu_{i2}\nu_{i3}}n_{i2}n_{i3}\alpha_1 & A_2 \\ \sqrt{\nu_{i1}\nu_{i3}}n_{i1}n_{i3}\alpha_1 & \sqrt{\nu_{i2}\nu_{i3}}n_{i2}n_{i3}\alpha_1 & \nu_{i3}n_{i3}[1 + (n_{i3} - 1)\alpha_0] & A_3 \\ A_1 & A_2 & A_3 & A_4 \end{pmatrix}$$

where

$$\begin{aligned} A_1 &= (\sqrt{\nu_{i1}\nu_{i2}}n_{i1}n_{i2} + \sqrt{\nu_{i1}\nu_{i3}}n_{i1}n_{i3})\alpha_1 \\ A_2 &= \nu_{i2}n_{i2}[1 + (n_{i2} - 1)\alpha_0] + \sqrt{\nu_{i2}\nu_{i3}}n_{i2}n_{i3}\alpha_1 \\ A_3 &= \sqrt{\nu_{i2}\nu_{i3}}n_{i2}n_{i3}\alpha_1 + \nu_{i3}n_{i3}[1 + (n_{i3} - 1)\alpha_0] \\ A_4 &= \nu_{i2}n_{i2}[1 + (n_{i2} - 1)\alpha_0] + \nu_{i3}n_{i3}[1 + (n_{i3} - 1)\alpha_0] + 2\sqrt{\nu_{i2}\nu_{i3}}n_{i2}n_{i3}\alpha_1, \end{aligned}$$

or

$$\frac{1}{\phi}\begin{pmatrix} \nu_{i1}n_{i1}[1+(n_{i1}-1)\alpha_0] & \sqrt{\nu_{i1}\nu_{i2}}n_{i1}n_{i2}\alpha_1 & \sqrt{\nu_{i1}\nu_{i3}}n_{i1}n_{i3}\alpha_1 & \sqrt{\nu_{i1}\nu_{i3}}n_{i1}n_{i3}\alpha_1 \\ \sqrt{\nu_{i1}\nu_{i2}}n_{i1}n_{i2}\alpha_1 & \nu_{i2}n_{i2}[1+(n_{i2}-1)\alpha_0] & \sqrt{\nu_{i2}\nu_{i3}}n_{i2}n_{i3}\alpha_1 & \sqrt{\nu_{i2}\nu_{i3}}n_{i2}n_{i3}\alpha_1 \\ \sqrt{\nu_{i1}\nu_{i3}}n_{i1}n_{i3}\alpha_1 & \sqrt{\nu_{i2}\nu_{i3}}n_{i2}n_{i3}\alpha_1 & \nu_{i3}n_{i3}[1+(n_{i3}-1)\alpha_0] & \nu_{i3}n_{i3}[1+(n_{i3}-1)\alpha_0] \\ \sqrt{v_{i1}\nu_{i3}}n_{i1}n_{i3}\alpha_1 & \sqrt{v_{i2}\nu_{i3}}n_{i2}n_{i3}\alpha_1 & \nu_{i3}n_{i3}[1+(n_{i3}-1)\alpha_0] & \nu_{i3}n_{i3}[1+(n_{i3}-1)\alpha_0] \end{pmatrix}.$$

To express $\overline{\boldsymbol{\Sigma}}_1$ and $\overline{\boldsymbol{\Sigma}}_0$ more succinctly, we define a series of notation as followed.

$$\begin{aligned}
a &= \sum_{i=1}^{I}\nu_{i1}n_{i1}/\phi,\ a_1=\sum_{i=1}^{I_1}\nu_{i1}n_{i1}/\phi=\sum_{i=1}^{I}X_{i2}\nu_{i1}n_{i1}/\phi \\
b &= \sum_{i=1}^{I}\frac{\nu_{i2}n_{i2}}{\phi},\ b_1=\sum_{i=1}^{I_1}\nu_{i2}n_{i2}/\phi=\sum_{i=1}^{I}X_{i2}\nu_{i2}n_{i2}/\phi \\
c &= \sum_{i=1}^{I}\frac{\nu_{i3}n_{i3}}{\phi},\ c_1=\sum_{i=1}^{I_1}\nu_{i3}n_{i3}/\phi=\sum_{i=1}^{I}X_{i2}\nu_{i3}n_{i3}/\phi \\
d &= \sum_{i=1}^{I}\nu_{i1}n_{i1}(n_{i1}-1)/\phi,\ d_1=\sum_{i=1}^{I_1}\nu_{i1}n_{i1}(n_{i1}-1)/\phi=\sum_{i=1}^{I}X_{i2}\nu_{i1}n_{i1}(n_{i1}-1)/\phi \\
e &= \sum_{i=1}^{I}\nu_{i2}n_{i2}(n_{i2}-1)/\phi,\ e_1=\sum_{i=1}^{I_1}\nu_{i2}n_{i2}(n_{i2}-1)/\phi=\sum_{i=1}^{I}X_{i2}\nu_{i2}n_{i2}(n_{i2}-1)/\phi \\
f &= \sum_{i=1}^{I}\nu_{i3}n_{i3}(n_{i3}-1)/\phi,\ f_1=\sum_{i=1}^{I_1}\nu_{i3}n_{i3}(n_{i3}-1)/\phi=\sum_{i=1}^{I}X_{i2}\nu_{i3}n_{i3}(n_{i3}-1)/\phi \\
x &= \sum_{i=1}^{I}\sqrt{\nu_{i1}\nu_{i2}}n_{i1}n_{i2}/\phi,\ x_1=\sum_{i=1}^{I_1}\sqrt{\nu_{i1}\nu_{i2}}n_{i1}n_{i2}/\phi=\sum_{i=1}^{I}X_{i2}\sqrt{\nu_{i1}\nu_{i2}}n_{i1}n_{i2}/\phi \\
y &= \sum_{i=1}^{I}\sqrt{\nu_{i1}\nu_{i3}}n_{i1}n_{i3}/\phi,\ y_1=\sum_{i=1}^{I_1}\sqrt{\nu_{i1}\nu_{i3}}n_{i1}n_{i3}/\phi=\sum_{i=1}^{I}X_{i2}\sqrt{\nu_{i1}\nu_{i3}}n_{i1}n_{i3}/\phi \\
z &= \sum_{i=1}^{I}\sqrt{\nu_{i2}\nu_{i3}}n_{i2}n_{i3}/\phi,\ z_1=\sum_{i=1}^{I_1}\sqrt{\nu_{i2}\nu_{i3}}n_{i2}n_{i3}/\phi=\sum_{i=1}^{I}X_{i2}\sqrt{\nu_{i2}\nu_{i3}}n_{i2}n_{i3}/\phi,
\end{aligned}$$

where $X_{i2}$ denotes the indicator of whether the $i$-th cluster receives treatment during the second time period. Then, $\overline{\boldsymbol{\Sigma}}_1$ and $\overline{\boldsymbol{\Sigma}}_0$ can be expressed as

$$\overline{\boldsymbol{\Sigma}}_1=\begin{pmatrix} a & 0 & 0 & 0 \\ 0 & b & 0 & b_1 \\ 0 & 0 & c & c \\ 0 & b_1 & c & b_1+c \end{pmatrix}$$

and

$$\overline{\boldsymbol{\Sigma}}_0=\begin{pmatrix} a+d\alpha_0 & x\alpha_1 & y\alpha_1 & (x_1+y)\alpha_1 \\ x\alpha_1 & b+e\alpha_0 & z\alpha_1 & b_1+e_1\alpha_0+z\alpha_1 \\ y\alpha_1 & z\alpha_1 & c+f\alpha_0 & c+f\alpha_0+z_1\alpha_1 \\ (x_1+y)\alpha_1 & b_1+e_1\alpha_0+z\alpha_1 & c+f\alpha_0+z_1\alpha_1 & b_1+c+e_1\alpha_0+f\alpha_0+2z_1\alpha_1 \end{pmatrix}.$$

Each cells of the matrix product $\overline{\boldsymbol{\Sigma}}_1^{-1}\overline{\boldsymbol{\Sigma}}_0\overline{\boldsymbol{\Sigma}}_1^{-1}$ can be obtained from direct calculation. Therefore, the sandwich variance of $\hat{\delta}$, which is the cell located at the bottom right corner of the obtained matrix can be calculated. The simplified variance of the treatment effect estimator is therefore given by

$$\text{var}(\hat{\delta}) = \frac{(b_1^2 e - 2b_1^2 b e_1 + b^2 e_1)\alpha_0 + b_1 b^2 - b_1^2 b}{(b_1 b - b_1^2)^2} \tag{3}$$

Likewise, when the true correlation structure is exponential decay, the same steps can be followed to derive the variance of the treatment effect estimator. The only difference there is that we have an alternative form of $\overline{\boldsymbol{\Sigma}}_0$ computed as following.

$$\begin{pmatrix} a + d\alpha_0 & x\alpha_0\rho & y\alpha_0\rho^2 & x_1\alpha_0\rho + y\alpha_0\rho^2 \\ x\alpha_0\rho & b + e\alpha_0 & z\alpha_0\rho & b_1 + e_1\alpha_0 + z\alpha_0\rho \\ y\alpha_0\rho^2 & z\alpha_0\rho & c + f\alpha_0 & c + f\alpha_0 + z_1\alpha_0\rho \\ x_1\alpha_0\rho + y\alpha_0\rho^2 & b_1 + e_1\alpha_0 + z\alpha_0\rho & c + f\alpha_0 + z_1\alpha_0\rho & b_1 + c + e_1\alpha_0 + f\alpha_0 + 2z_1\alpha_0\rho \end{pmatrix}$$

The simplified variance of treatment effect is exactly the same as (3) and thus omitted here for brevity.

#### Web Appendix C. Supplementary simulation results under nested exchangeable true correlation structure

In this section, supplementary figures and tables related to the simulation results under the nested exchangeable true correlation structure are showed.

##### Web Appendix C.1. Cluster size variability

The following Figures are the counterparts to Figure 3 in the main text when pattern 1, 2, and 3 within-cluster imbalance are introduced.

**Web Figure 1** The median and interquartile range (IQR) of relative efficiency (RE) as a function of coefficient of variation (CV) measuring between-cluster imbalance, when the true correlation model is nested exchangeable (NEX). Design factors considered are as follows: number of clusters $I = 12$ and 96, number of periods $J = 5$. The within-period intraclass correlation coefficient (WP-ICC) $\alpha_0 = 0.05$, and between-period intraclass correlation coefficient (BP-ICC) $\alpha_1 \in \{0.001, 0.025, 0.05\}$. The working correlation structure is either NEX or independence (IND). Within-cluster imbalance (pattern 1: constant) is introduced.

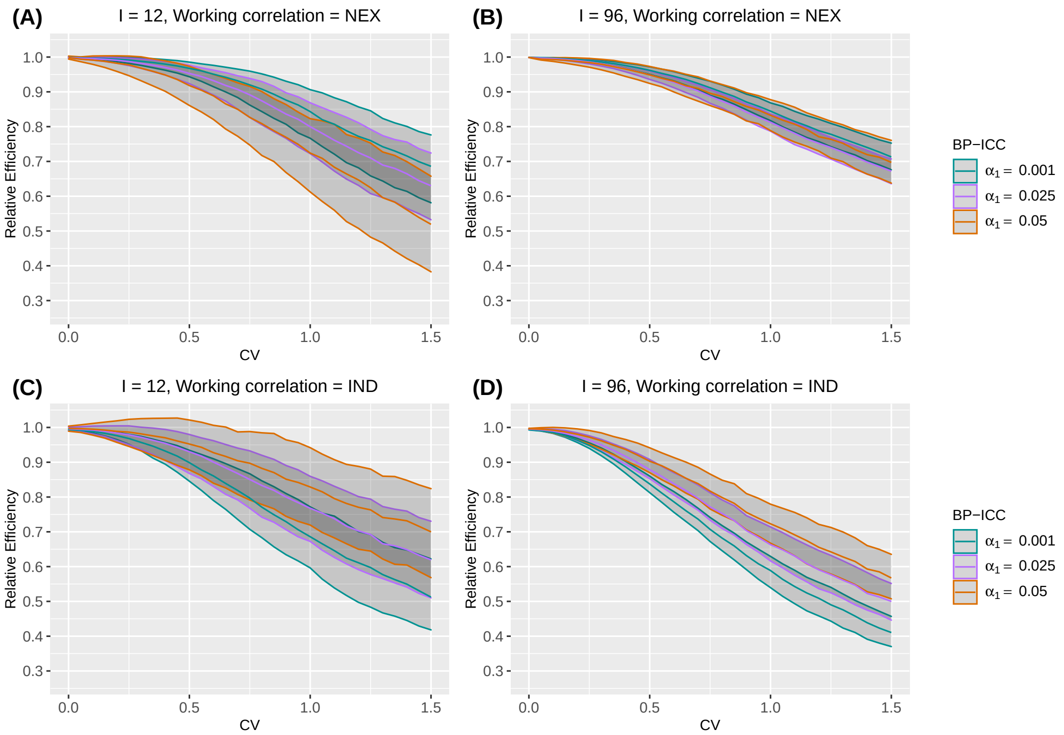

**Web Figure 2** The median and interquartile range (IQR) of relative efficiency (RE) as a function of coefficient of variation (CV) measuring between-cluster imbalance, when the true correlation model is nested exchangeable (NEX). Design factors considered are as follows: number of clusters $I = 12$ and 96, number of periods $J = 5$. The within-period intraclass correlation coefficient (WP-ICC) $\alpha_0 = 0.05$, and between-period intraclass correlation coefficient (BP-ICC) $\alpha_1 \in \{0.001, 0.025, 0.05\}$. The working correlation structure is either NEX or independence (IND). Within-cluster imbalance (pattern 2: monotonically increasing) is introduced.

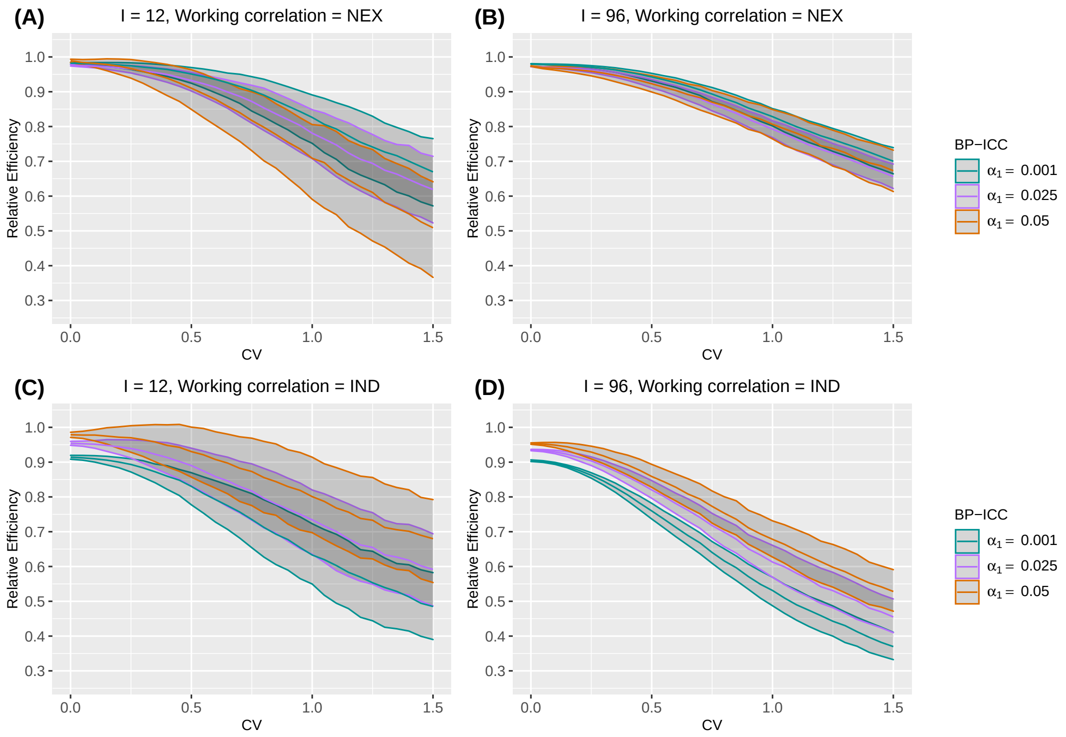

**Web Figure 3** The median and interquartile range (IQR) of relative efficiency (RE) as a function of coefficient of variation (CV) measuring between-cluster imbalance, when the true correlation model is nested exchangeable (NEX). Design factors considered are as follows: number of clusters $I = 12$ and 96, number of periods $J = 5$. The within-period intraclass correlation coefficient (WP-ICC) $\alpha_0 = 0.05$, and between-period intraclass correlation coefficient (BP-ICC) $\alpha_1 \in \{0.001, 0.025, 0.05\}$. The working correlation structure is either NEX or independence (IND). Within-cluster imbalance (pattern 3: monotonically decreasing) is introduced.

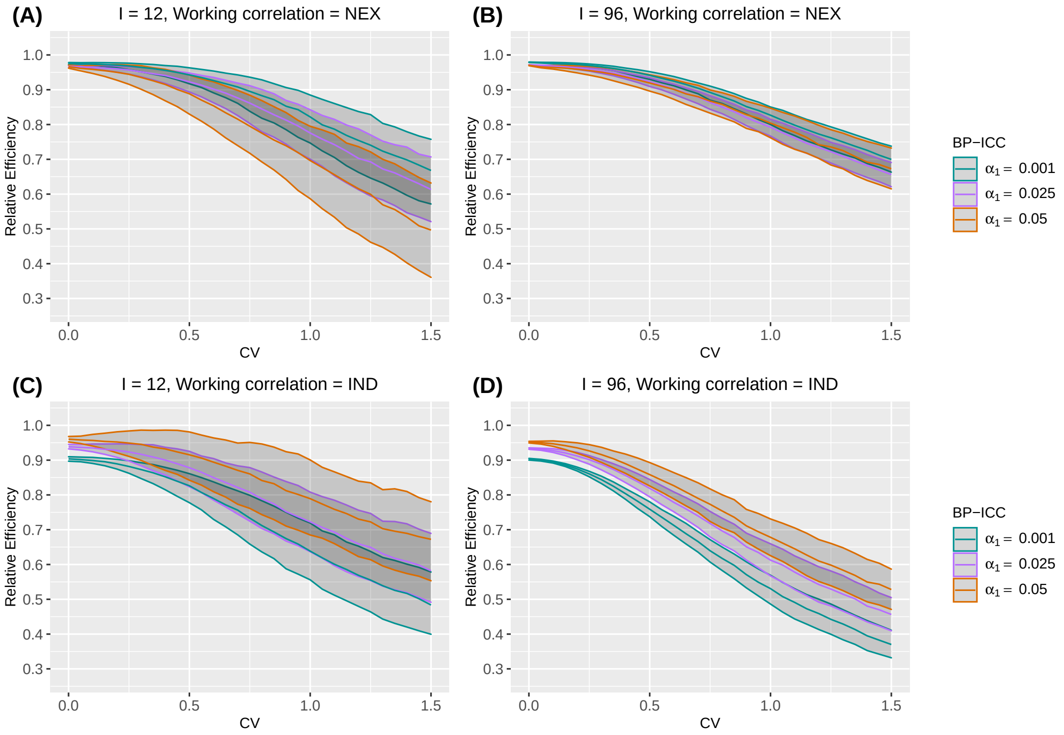

##### Web Appendix C.2. Intraclass correlation coefficients

The following Figures shows the counterparts to Figure 4 and 5 in the main article when pattern 1, 2, 3, and 4 within-cluster imbalance are introduced.

**Web Figure 4** The median of relative efficiency (RE) as a function of the within-period intraclass correlation coefficient (WP-ICC) $\alpha_0 \in \{0.01, 0.05, 0.1, 0.2\}$ and the ratio of between-period intraclass correlation coefficient (BP-ICC) to WP-ICC, $\alpha_1/\alpha_0 \in [0, 1]$, when both the true correlation model and the working correlation model are nested exchangeable (NEX). Design factors considered are as follows: number of clusters $I = 12$ and 96, number of periods $J = 5$, and the degree of between-cluster imbalance is defined by CV $\in \{0.25, 0.75, 1.25\}$. Within-cluster imbalance (pattern 1: constant) is introduced.

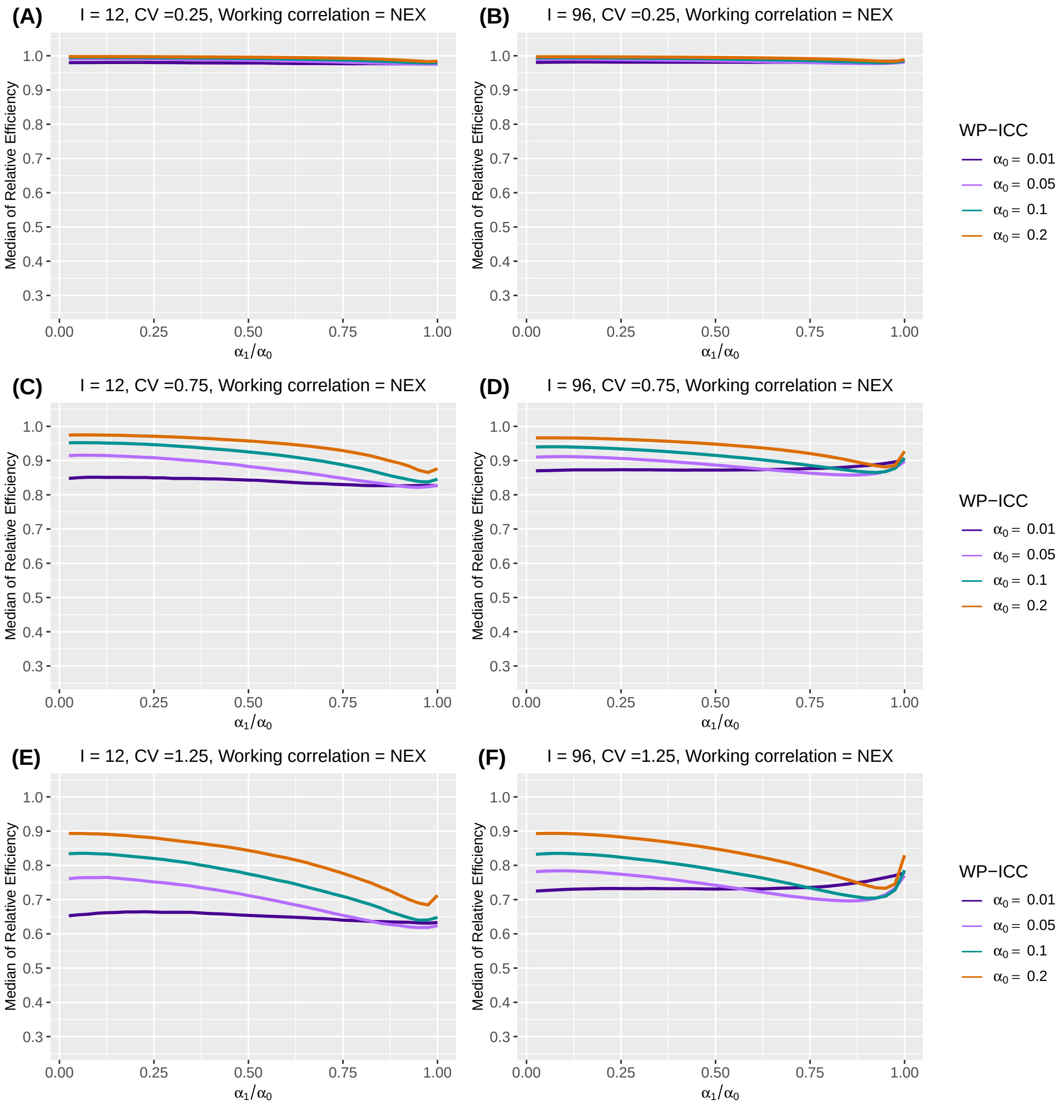

**Web Figure 5** The median of relative efficiency (RE) as a function of the within-period intraclass correlation coefficient (WP-ICC) $\alpha_0 \in \{0.01, 0.05, 0.1, 0.2\}$ and the ratio of between-period intraclass correlation coefficient (BP-ICC) to WP-ICC, $\alpha_1/\alpha_0 \in [0, 1]$, when both the true correlation model and the working correlation model are nested exchangeable (NEX). Design factors considered are as follows: number of clusters $I = 12$ and $96$, number of periods $J = 5$, and the degree of between-cluster imbalance is defined by CV $\in \{0.25, 0.75, 1.25\}$. Within-cluster imbalance (pattern 2: monotonically increasing) is introduced.

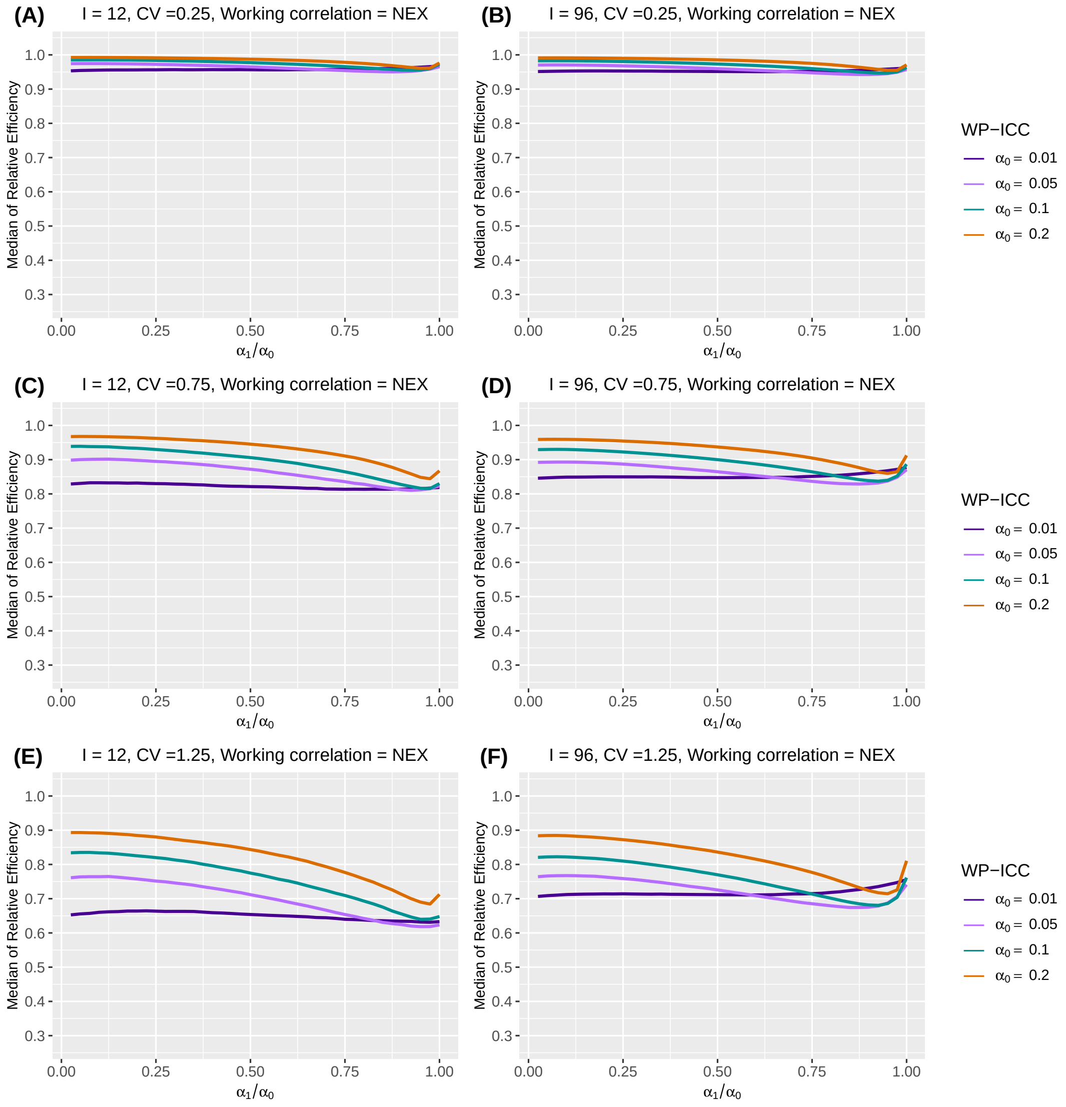

**Web Figure 6** The median of relative efficiency (RE) as a function of the within-period intraclass correlation coefficient (WP-ICC) $\alpha_0 \in \{0.01, 0.05, 0.1, 0.2\}$ and the ratio of between-period intraclass correlation coefficient (BP-ICC) to WP-ICC, $\alpha_1/\alpha_0 \in [0, 1]$, when both the true correlation model and the working correlation model are nested exchangeable (NEX). Design factors considered are as follows: number of clusters $I = 12$ and $96$, number of periods $J = 5$, and the degree of between-cluster imbalance is defined by CV $\in \{0.25, 0.75, 1.25\}$. Within-cluster imbalance (pattern 3: monotonically decreasing) is introduced.

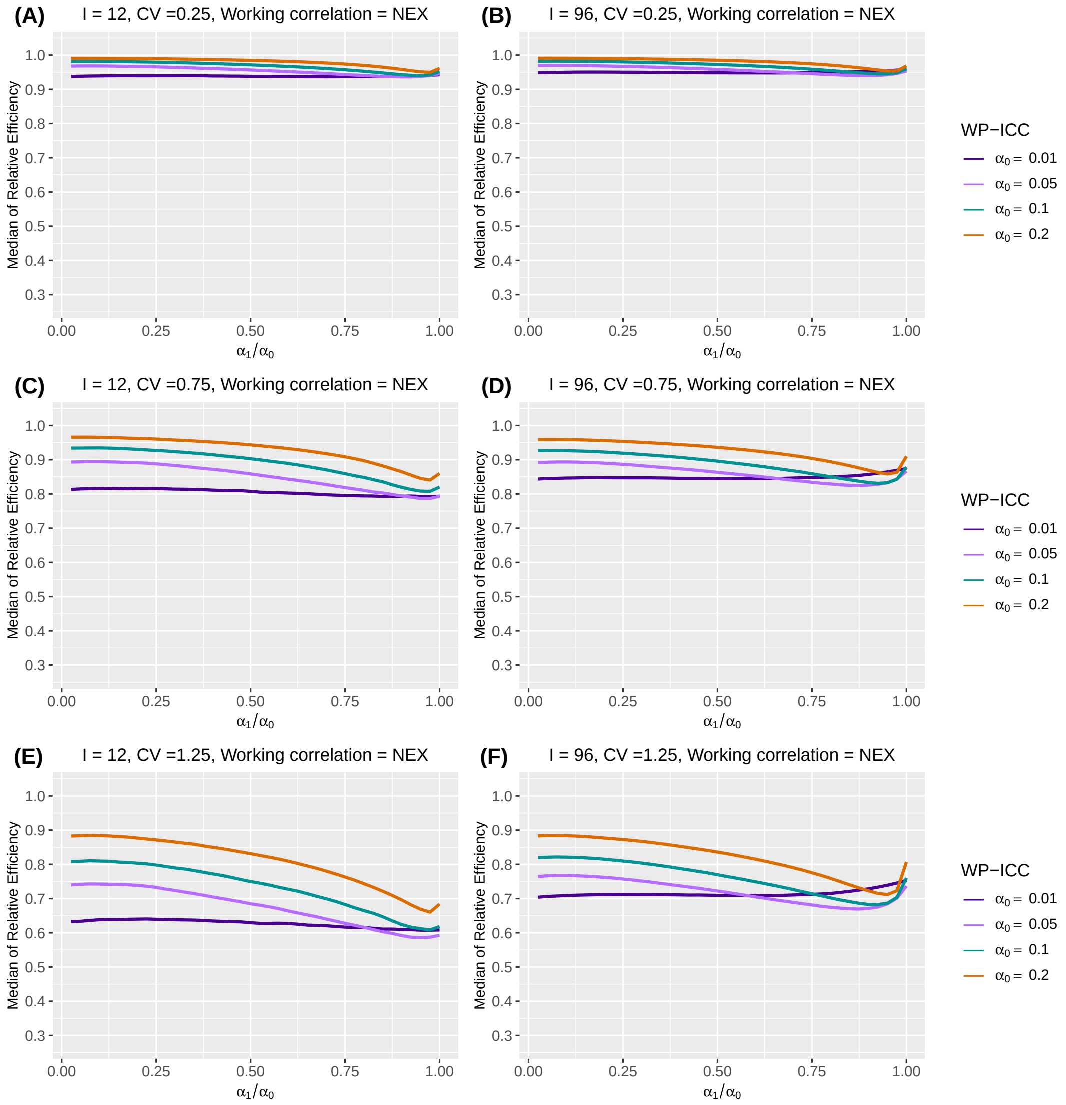

**Web Figure 7** The median of relative efficiency (RE) as a function of the within-period intraclass correlation coefficient (WP-ICC) $\alpha_0 \in \{0.01, 0.05, 0.1, 0.2\}$ and the ratio of between-period intraclass correlation coefficient (BP-ICC) to WP-ICC, $\alpha_1/\alpha_0 \in [0, 1]$, when both the true correlation model and the working correlation model are nested exchangeable (NEX). Design factors considered are as follows: number of clusters $I = 12$ and 96, number of periods $J = 5$, and the degree of between-cluster imbalance is defined by CV $\in \{0.25, 0.75, 1.25\}$. Within-cluster imbalance (pattern 4: randomly permuted) is introduced.

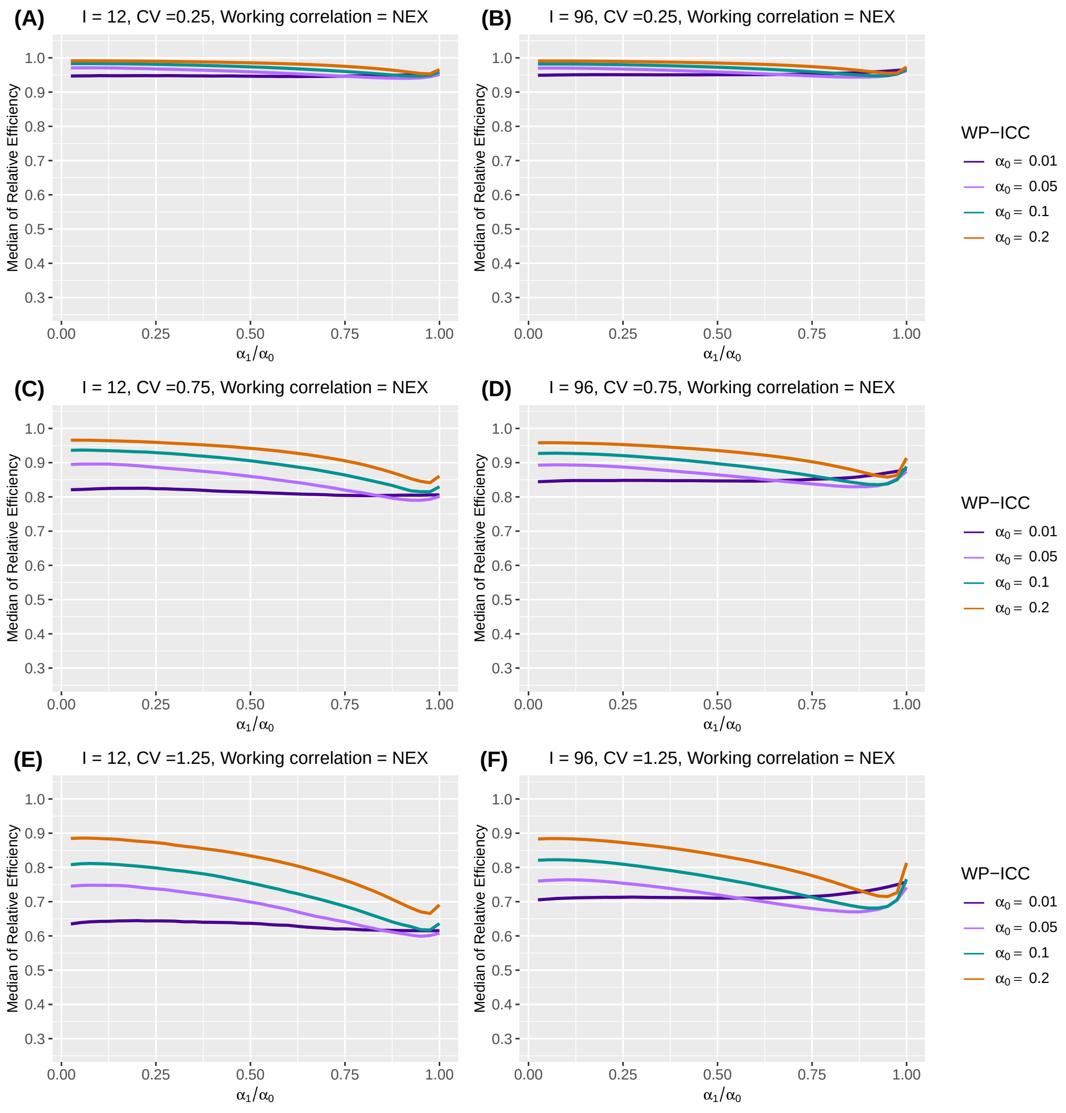

**Web Figure 8** The median of relative efficiency (RE) as a function of the within-period intraclass correlation coefficient (WP-ICC) $\alpha_0 \in \{0.01, 0.05, 0.1, 0.2\}$ and the ratio of between-period intraclass correlation coefficient (BP-ICC) to WP-ICC, $\alpha_1/\alpha_0 \in [0, 1]$, when the true correlation model is nested exchangeable (NEX) and the working correlation model is independence (IND). Design factors considered are as follows: number of clusters $I = 12$ and $96$, number of periods $J = 5$, and the degree of between-cluster imbalance is defined by CV $\in \{0.25, 0.75, 1.25\}$. Within-cluster imbalance (pattern 1: constant) is introduced.

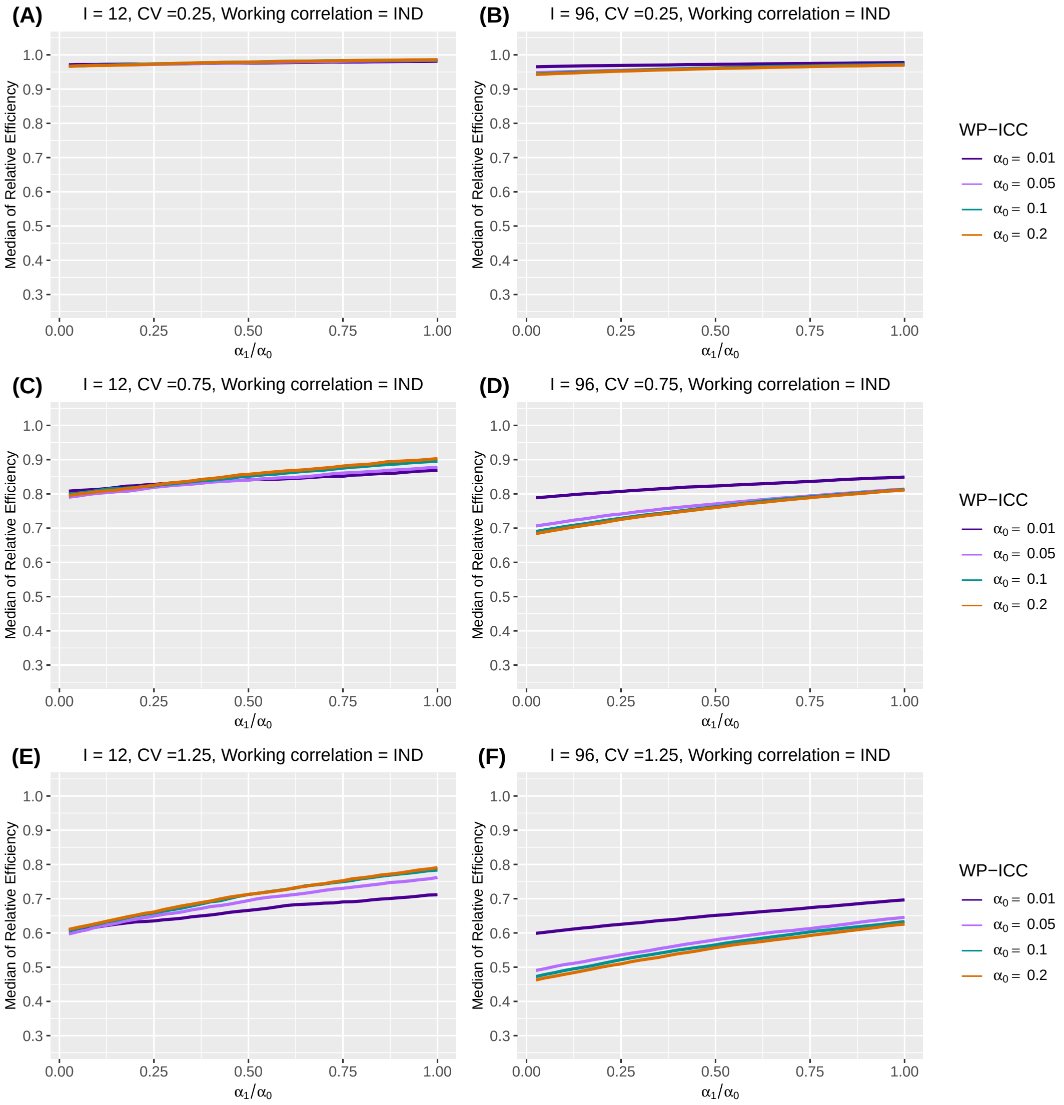

**Web Figure 9** The median of relative efficiency (RE) as a function of the within-period intraclass correlation coefficient (WP-ICC) $\alpha_0 \in \{0.01, 0.05, 0.1, 0.2\}$ and the ratio of between-period intraclass correlation coefficient (BP-ICC) to WP-ICC, $\alpha_1/\alpha_0 \in [0, 1]$, when the true correlation model is nested exchangeable (NEX) and the working correlation model is independence (IND). Design factors considered are as follows: number of clusters $I = 12$ and $96$, number of periods $J = 5$, and the degree of between-cluster imbalance is defined by CV $\in \{0.25, 0.75, 1.25\}$. Within-cluster imbalance (pattern 2: monotonically increasing) is introduced.

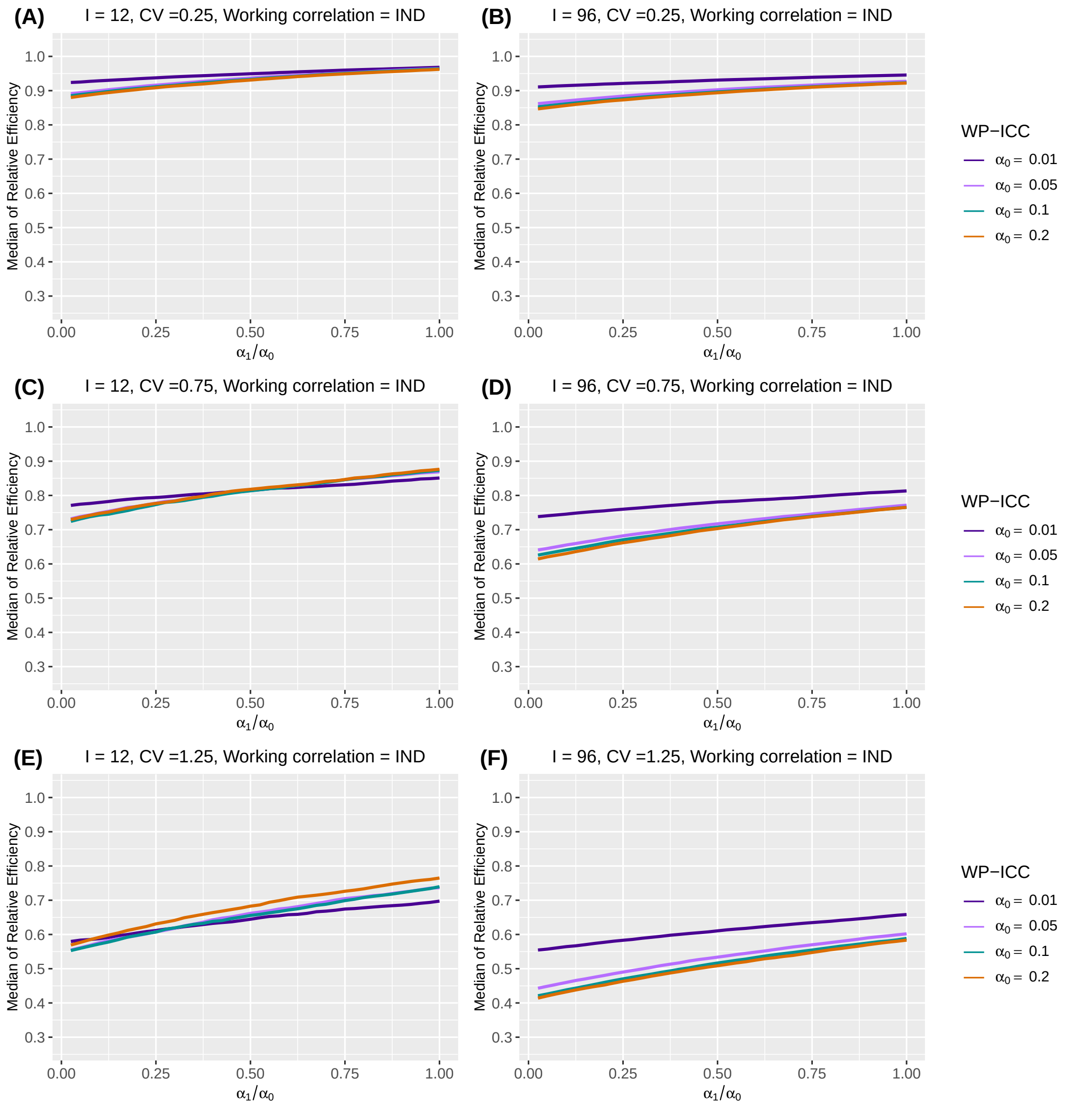

**Web Figure 10** The median of relative efficiency (RE) as a function of the within-period intraclass correlation coefficient (WP-ICC) $\alpha_0 \in \{0.01, 0.05, 0.1, 0.2\}$ and the ratio of between-period intraclass correlation coefficient (BP-ICC) to WP-ICC, $\alpha_1/\alpha_0 \in [0, 1]$, when the true correlation model is nested exchangeable (NEX) and the working correlation model is independence (IND). Design factors considered are as follows: number of clusters $I = 12$ and $96$, number of periods $J = 5$, and the degree of between-cluster imbalance is defined by CV $\in \{0.25, 0.75, 1.25\}$. Within-cluster imbalance (pattern 3: monotonically decreasing) is introduced.

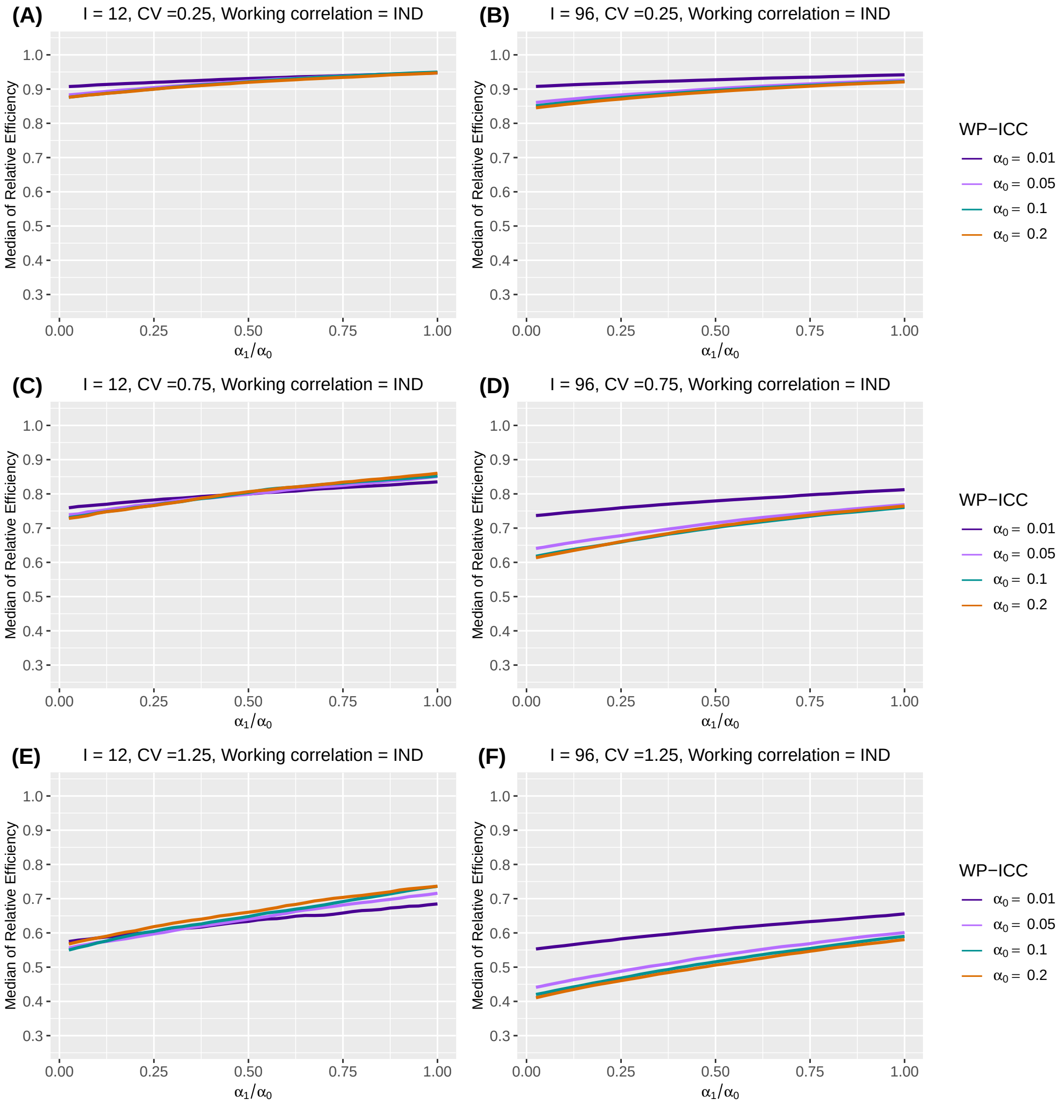

**Web Figure 11** The median of relative efficiency (RE) as a function of the within-period intraclass correlation coefficient (WP-ICC) $\alpha_0 \in \{0.01, 0.05, 0.1, 0.2\}$ and the ratio of between-period intraclass correlation coefficient (BP-ICC) to WP-ICC, $\alpha_1/\alpha_0 \in [0, 1]$, when the true correlation model is nested exchangeable (NEX) and the working correlation model is independence (IND). Design factors considered are as follows: number of clusters $I = 12$ and $96$, number of periods $J = 5$, and the degree of between-cluster imbalance is defined by CV $\in \{0.25, 0.75, 1.25\}$. Within-cluster imbalance (pattern 4: randomly permuted) is introduced.

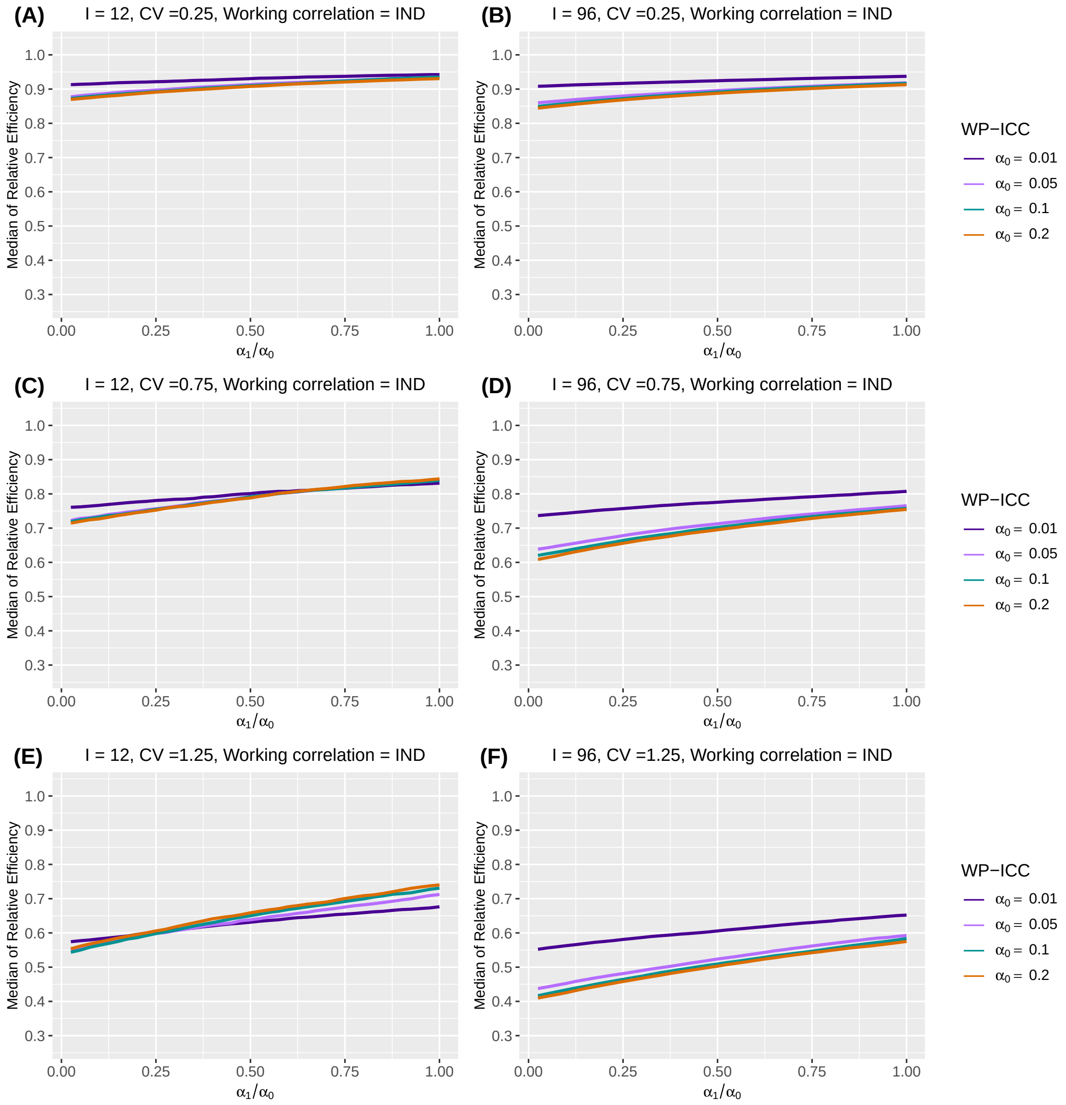

##### Web Appendix C.3. Number of clusters

As mentioned in the first paragraph of Section 5.3 of the main article, the figure below shows the counterpart to Figure 2 in the main article when the number of clusters $I = 24$ and $48$.

**Web Figure 12** The median and interquartile range (IQR) of relative efficiency (RE) as a function of coefficient of variation (CV) measuring between-cluster imbalance, when the true correlation model is nested exchangeable (NEX). Design factors considered are as follows: number of clusters $I = 24$ and $48$, number of periods $J = 5$. The within-period intraclass correlation coefficient (WP-ICC) $\alpha_0 = 0.05$, and between-period intraclass correlation coefficient (BP-ICC) $\alpha_1 \in \{0.001, 0.025, 0.05\}$. The working correlation structure is either NEX or independence (IND). No within-cluster variability in cluster-period sizes is introduced.

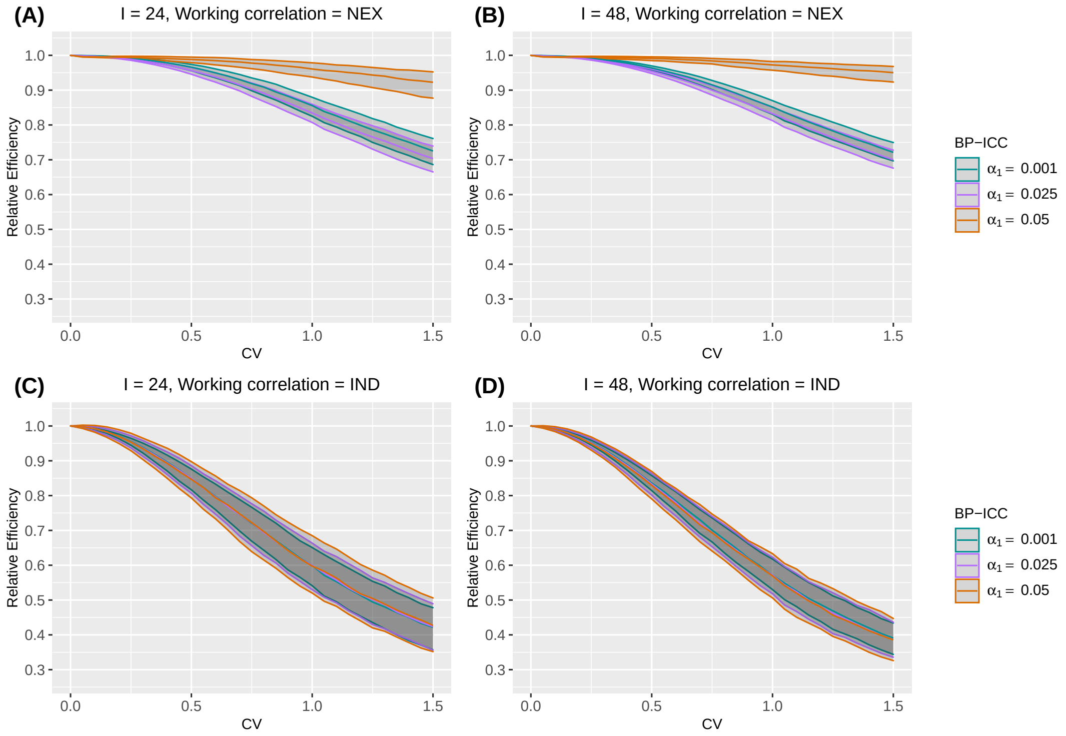

##### Web Appendix C.4. Number of periods

The following tables shows the counterparts of Table 2 in the main article when the number of clusters $I$ is 12, 48 or 96.

**Web Table 1** Median and interquartile range (IQR) (in parentheses) of relative efficiency (RE) as a function of periods $J$, under different degrees of between- and within-cluster imbalance, when the true correlation model is nested exchangeable (NEX). Number of clusters is $I = 12$. The within-period intraclass correlation coefficient (WP-ICC) $\alpha_0$ is $0.05$, and the within-period intraclass correlation coefficient (BP-ICC) $\alpha_1$ is $0.025$. Between cluster imbalance is measured by coefficient of variation, CV $\in \{0.25, 0.75, 1.25\}$. The working correlation structure is either NEX or independence (IND).

| Working correlation | $J$ | CV | No within-cluster imbalance | Within-cluster imbalance pattern 1 | Within-cluster imbalance pattern 2 | Within-cluster imbalance pattern 4 |
|---|---|---|---|---|---|---|
| NEX | 3 | 0.25 | 0.989 (0.985, 0.993) | 0.988 (0.971, 1.002) | 0.967 (0.949, 0.979) | 0.965 (0.949, 0.980) |
| | | 0.75 | 0.906 (0.876, 0.931) | 0.896 (0.816, 0.955) | 0.867 (0.788, 0.926) | 0.873 (0.794, 0.931) |
| | | 1.25 | 0.760 (0.707, 0.807) | 0.726 (0.606, 0.837) | 0.699 (0.589, 0.808) | 0.697 (0.584, 0.821) |
| | 5 | 0.25 | 0.989 (0.985, 0.993) | 0.987 (0.976, 0.997) | 0.964 (0.952, 0.974) | 0.958 (0.946, 0.970) |
| | | 0.75 | 0.907 (0.878, 0.933) | 0.889 (0.832, 0.933) | 0.872 (0.810, 0.914) | 0.865 (0.805, 0.911) |
| | | 1.25 | 0.764 (0.713, 0.813) | 0.711 (0.623, 0.793) | 0.697 (0.611, 0.782) | 0.692 (0.610, 0.776) |
| | 13 | 0.25 | 0.989 (0.986, 0.992) | 0.986 (0.982, 0.990) | 0.973 (0.969, 0.977) | 0.977 (0.972, 0.982) |
| | | 0.75 | 0.909 (0.882, 0.933) | 0.892 (0.854, 0.921) | 0.879 (0.842, 0.905) | 0.880 (0.845, 0.910) |
| | | 1.25 | 0.767 (0.722, 0.815) | 0.728 (0.667, 0.782) | 0.712 (0.650, 0.772) | 0.718 (0.665, 0.774) |
| IND | 3 | 0.25 | 0.961 (0.948, 0.972) | 0.970 (0.951, 0.988) | 0.891 (0.871, 0.908) | 0.889 (0.869, 0.908) |
| | | 0.75 | 0.755 (0.694, 0.806) | 0.825 (0.749, 0.899) | 0.749 (0.674, 0.818) | 0.759 (0.677, 0.821) |
| | | 1.25 | 0.547 (0.474, 0.621) | 0.666 (0.583, 0.769) | 0.611 (0.529, 0.704) | 0.604 (0.532, 0.707) |
| | 5 | 0.25 | 0.962 (0.937, 0.983) | 0.978 (0.952, 1.004) | 0.934 (0.906, 0.960) | 0.911 (0.881, 0.943) |
| | | 0.75 | 0.753 (0.686, 0.826) | 0.847 (0.769, 0.920) | 0.819 (0.740, 0.890) | 0.791 (0.713, 0.863) |
| | | 1.25 | 0.548 (0.467, 0.636) | 0.687 (0.577, 0.784) | 0.660 (0.554, 0.757) | 0.640 (0.554, 0.732) |
| | 13 | 0.25 | 0.961 (0.928, 0.995) | 0.989 (0.982, 0.997) | 0.966 (0.955, 0.975) | 0.977 (0.963, 0.991) |
| | | 0.75 | 0.759 (0.669, 0.838) | 0.924 (0.895, 0.950) | 0.906 (0.864, 0.933) | 0.909 (0.876, 0.939) |
| | | 1.25 | 0.558 (0.457, 0.655) | 0.812 (0.744, 0.863) | 0.794 (0.725, 0.850) | 0.794 (0.726, 0.849) |

**Web Table 2** Median and interquartile range (IQR) (in parentheses) of relative efficiency (RE) as a function of periods $J$, under different degrees of between- and within-cluster imbalance, when the true correlation model is nested exchangeable (NEX). Number of clusters is $I = 48$. The within-period intraclass correlation coefficient (WP-ICC) $\alpha_0$ is $0.05$, and the within-period intraclass correlation coefficient (BP-ICC) $\alpha_1$ is $0.025$. Between cluster imbalance is measured by coefficient of variation, CV $\in \{0.25, 0.75, 1.25\}$. The working correlation structure is either NEX or independence (IND).

| Working correlation | $J$ | CV | No within-cluster imbalance | Within-cluster imbalance pattern 1 | Within-cluster imbalance pattern 2 | Within-cluster imbalance pattern 4 |
|---|---|---|---|---|---|---|
| NEX | 3 | 0.25 | 0.988 (0.985, 0.990) | 0.986 (0.978, 0.993) | 0.965 (0.956, 0.973) | 0.962 (0.954, 0.970) |
| | | 0.75 | 0.898 (0.884, 0.911) | 0.882 (0.847, 0.914) | 0.865 (0.833, 0.895) | 0.861 (0.829, 0.890) |
| | | 1.25 | 0.759 (0.731, 0.784) | 0.733 (0.674, 0.781) | 0.713 (0.660, 0.764) | 0.713 (0.659, 0.760) |
| | 5 | 0.25 | 0.988 (0.986, 0.990) | 0.986 (0.979, 0.992) | 0.960 (0.954, 0.967) | 0.958 (0.951, 0.964) |
| | | 0.75 | 0.900 (0.886, 0.913) | 0.888 (0.860, 0.913) | 0.866 (0.837, 0.889) | 0.864 (0.837, 0.889) |
| | | 1.25 | 0.763 (0.738, 0.786) | 0.735 (0.685, 0.783) | 0.714 (0.673, 0.758) | 0.714 (0.665, 0.762) |
| | 13 | 0.25 | 0.988 (0.986, 0.990) | 0.986 (0.980, 0.990) | 0.976 (0.971, 0.981) | 0.976 (0.971, 0.982) |
| | | 0.75 | 0.902 (0.888, 0.915) | 0.882 (0.858, 0.906) | 0.875 (0.854, 0.898) | 0.875 (0.850, 0.899) |
| | | 1.25 | 0.770 (0.745, 0.790) | 0.725 (0.684, 0.764) | 0.719 (0.678, 0.757) | 0.721 (0.681, 0.757) |
| IND | 3 | 0.25 | 0.952 (0.945, 0.959) | 0.951 (0.939, 0.962) | 0.874 (0.860, 0.884) | 0.871 (0.859, 0.884) |
| | | 0.75 | 0.701 (0.665, 0.734) | 0.725 (0.669, 0.778) | 0.666 (0.619, 0.707) | 0.662 (0.611, 0.711) |
| | | 1.25 | 0.469 (0.419, 0.514) | 0.530 (0.461, 0.590) | 0.481 (0.421, 0.538) | 0.477 (0.415, 0.537) |
| | 5 | 0.25 | 0.951 (0.937, 0.963) | 0.965 (0.947, 0.982) | 0.911 (0.896, 0.927) | 0.899 (0.881, 0.915) |
| | | 0.75 | 0.694 (0.653, 0.740) | 0.792 (0.740, 0.837) | 0.741 (0.692, 0.789) | 0.732 (0.688, 0.778) |
| | | 1.25 | 0.462 (0.403, 0.521) | 0.608 (0.542, 0.676) | 0.568 (0.497, 0.625) | 0.557 (0.487, 0.620) |
| | 13 | 0.25 | 0.949 (0.928, 0.966) | 0.987 (0.964, 1.007) | 0.974 (0.953, 0.994) | 0.974 (0.954, 0.995) |
| | | 0.75 | 0.686 (0.634, 0.742) | 0.903 (0.845, 0.958) | 0.887 (0.834, 0.944) | 0.885 (0.830, 0.937) |
| | | 1.25 | 0.449 (0.390, 0.519) | 0.765 (0.693, 0.839) | 0.761 (0.689, 0.841) | 0.766 (0.694, 0.838) |

**Web Table 3** Median and interquartile range (IQR) (in parentheses) of relative efficiency (RE) as a function of periods $J$, under different degrees of between- and within-cluster imbalance, when the true correlation model is nested exchangeable (NEX). Number of clusters is $I = 96$. The within-period intraclass correlation coefficient (WP-ICC) $\alpha_0$ is $0.05$, and the within-period intraclass correlation coefficient (BP-ICC) $\alpha_1$ is $0.025$. Between cluster imbalance is measured by coefficient of variation, CV $\in \{0.25, 0.75, 1.25\}$. The working correlation structure is either NEX or independence (IND).

| Working correlation | $J$ | CV | No within-cluster imbalance | Within-cluster imbalance pattern 1 | Within-cluster imbalance pattern 2 | Within-cluster imbalance pattern 4 |
|---|---|---|---|---|---|---|
| NEX | 3 | 0.25 | 0.987 (0.986, 0.989) | 0.985 (0.979, 0.991) | 0.964 (0.958, 0.969) | 0.962 (0.956, 0.968) |
| | | 0.75 | 0.896 (0.885, 0.906) | 0.881 (0.858, 0.905) | 0.863 (0.839, 0.886) | 0.862 (0.837, 0.885) |
| | | 1.25 | 0.758 (0.739, 0.775) | 0.725 (0.688, 0.761) | 0.712 (0.677, 0.745) | 0.708 (0.668, 0.742) |
| | 5 | 0.25 | 0.988 (0.986, 0.989) | 0.985 (0.980, 0.990) | 0.960 (0.955, 0.965) | 0.958 (0.954, 0.963) |
| | | 0.75 | 0.899 (0.889, 0.907) | 0.886 (0.867, 0.906) | 0.867 (0.845, 0.885) | 0.864 (0.844, 0.882) |
| | | 1.25 | 0.762 (0.745, 0.780) | 0.738 (0.709, 0.769) | 0.721 (0.693, 0.751) | 0.723 (0.690, 0.752) |
| | 13 | 0.25 | 0.988 (0.987, 0.989) | 0.986 (0.981, 0.990) | 0.977 (0.973, 0.981) | 0.977 (0.972, 0.981) |
| | | 0.75 | 0.901 (0.892, 0.910) | 0.888 (0.870, 0.907) | 0.882 (0.864, 0.899) | 0.881 (0.863, 0.899) |
| | | 1.25 | 0.769 (0.752, 0.785) | 0.740 (0.712, 0.769) | 0.736 (0.709, 0.762) | 0.736 (0.708, 0.763) |
| IND | 3 | 0.25 | 0.951 (0.946, 0.956) | 0.947 (0.938, 0.956) | 0.870 (0.861, 0.879) | 0.870 (0.860, 0.879) |
| | | 0.75 | 0.689 (0.662, 0.714) | 0.706 (0.660, 0.743) | 0.643 (0.604, 0.679) | 0.641 (0.603, 0.681) |
| | | 1.25 | 0.450 (0.414, 0.488) | 0.482 (0.430, 0.534) | 0.441 (0.396, 0.486) | 0.438 (0.389, 0.485) |
| | 5 | 0.25 | 0.949 (0.940, 0.958) | 0.963 (0.951, 0.975) | 0.903 (0.891, 0.915) | 0.896 (0.883, 0.907) |
| | | 0.75 | 0.681 (0.648, 0.715) | 0.772 (0.735, 0.810) | 0.720 (0.682, 0.755) | 0.713 (0.675, 0.747) |
| | | 1.25 | 0.446 (0.401, 0.486) | 0.573 (0.523, 0.627) | 0.526 (0.483, 0.572) | 0.526 (0.474, 0.577) |
| | 13 | 0.25 | 0.947 (0.933, 0.959) | 0.984 (0.967, 1.002) | 0.974 (0.959, 0.992) | 0.970 (0.952, 0.988) |
| | | 0.75 | 0.668 (0.631, 0.711) | 0.890 (0.846, 0.939) | 0.878 (0.835, 0.920) | 0.871 (0.828, 0.921) |
| | | 1.25 | 0.435 (0.384, 0.481) | 0.759 (0.701, 0.820) | 0.745 (0.682, 0.811) | 0.740 (0.685, 0.803) |

##### Web Appendix C.5. Cluster-period size

Figures that illustrate the impact of mean of cluster-period sizes on RE mentioned in section 5.5 of the main article are showed below.

**Web Figure 13** The median of relative efficiency (RE) as a function of the within-period intraclass correlation coefficient (WP-ICC) $\alpha_0 \in \{0.01, 0.05, 0.1, 0.2\}$ and the ratio of between-period intraclass correlation coefficient (BP-ICC) to WP-ICC, $\alpha_1/\alpha_0 \in [0, 1]$, when both the true correlation model and the working correlation model are nested exchangeable (NEX). Design factors considered are as follows: number of clusters $I = 12$, number of periods $J = 5$, mean cluster-period sizes $\overline{n} \in \{50, 300\}$, and the degree of between-cluster imbalance is defined by coefficient of variation CV $\in \{0.25, 0.75, 1.25\}$. No within-cluster imbalance is introduced.

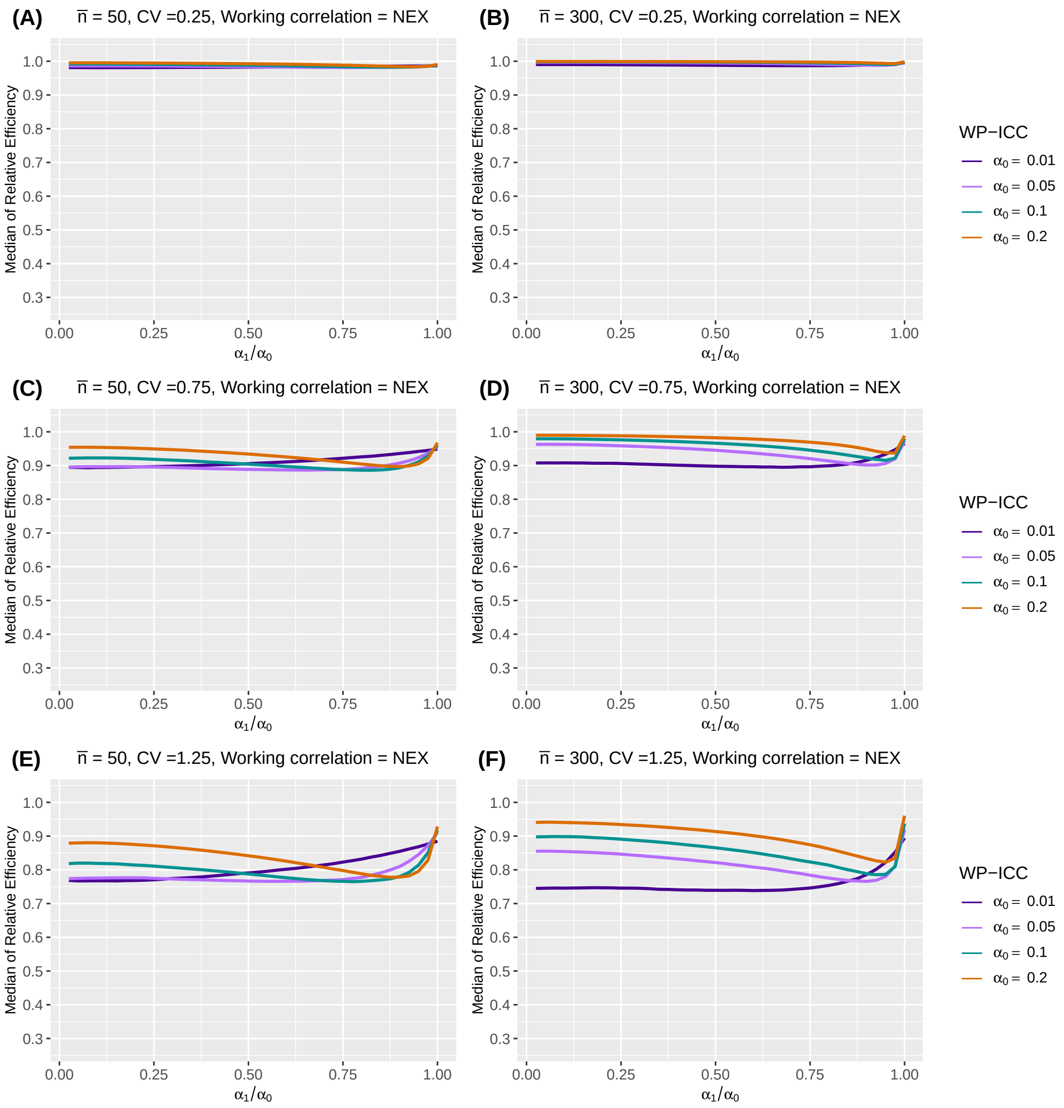

**Web Figure 14** The median of relative efficiency (RE) as a function of the within-period intraclass correlation coefficient (WP-ICC) $\alpha_0 \in \{0.01, 0.05, 0.1, 0.2\}$ and the ratio of between-period intraclass correlation coefficient (BP-ICC) to WP-ICC, $\alpha_1/\alpha_0 \in [0, 1]$, when both the true correlation model and the working correlation model are nested exchangeable (NEX). Design factors considered are as follows: number of clusters $I = 96$, number of periods $J = 5$, mean cluster-period sizes $\overline{n} \in \{50, 300\}$, and the degree of between-cluster imbalance is defined by coefficient of variation CV $\in \{0.25, 0.75, 1.25\}$. No within-cluster imbalance is introduced.

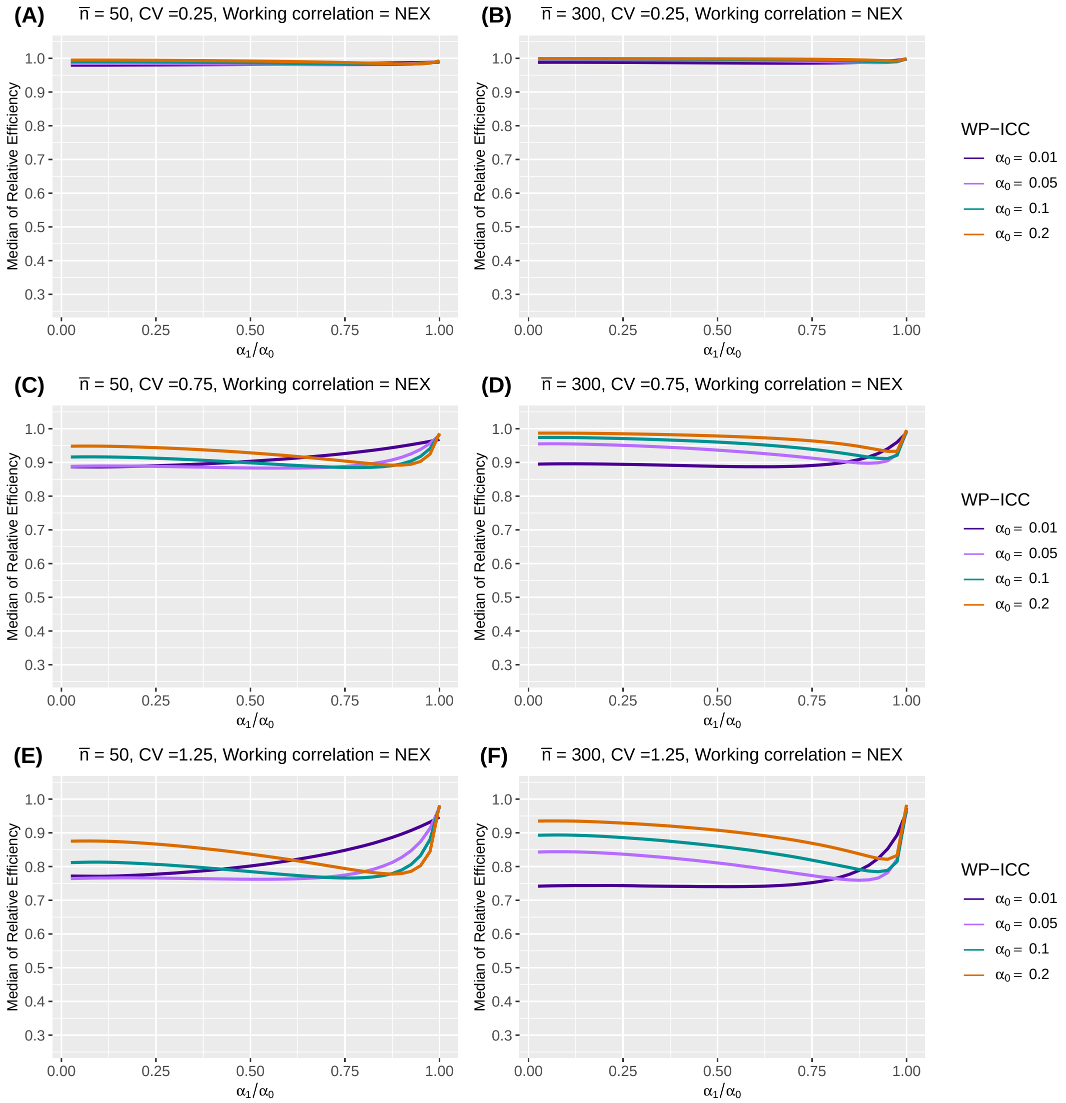

**Web Figure 15** The median of relative efficiency (RE) as a function of the within-period intraclass correlation coefficient (WP-ICC) $\alpha_0 \in \{0.01, 0.05, 0.1, 0.2\}$ and the ratio of between-period intraclass correlation coefficient (BP-ICC) to WP-ICC, $\alpha_1/\alpha_0 \in [0, 1]$, when both the true correlation model and the working correlation model are nested exchangeable (NEX). Design factors considered are as follows: number of clusters $I = 12$, number of periods $J = 5$, mean cluster-period sizes $\overline{n} \in \{50, 300\}$, and the degree of between-cluster imbalance is defined by coefficient of variation CV $\in \{0.25, 0.75, 1.25\}$. Within-cluster imbalance (pattern 4: randomly permuted) is introduced.

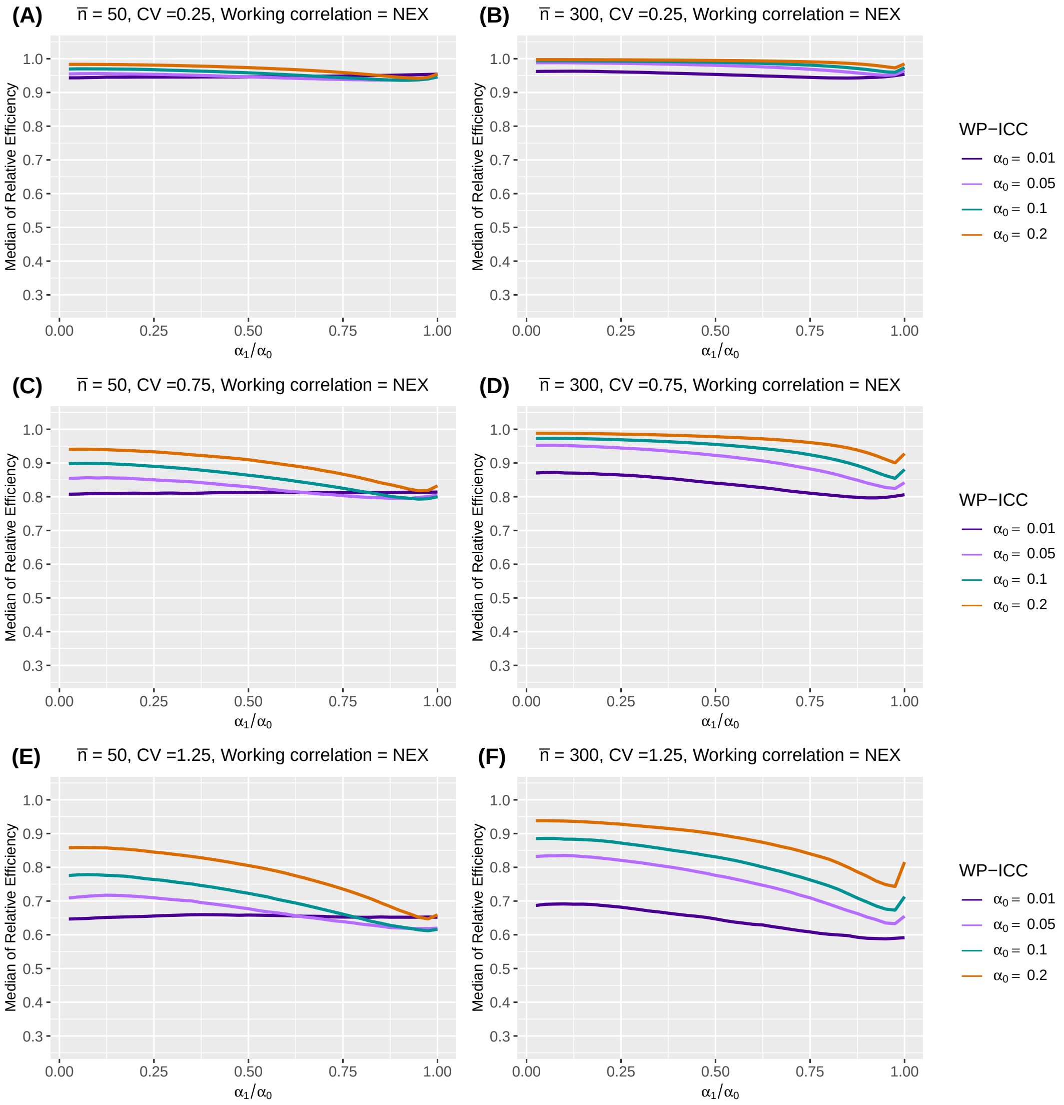

**Web Figure 16** The median of relative efficiency (RE) as a function of the within-period intraclass correlation coefficient (WP-ICC) $\alpha_0 \in \{0.01, 0.05, 0.1, 0.2\}$ and the ratio of between-period intraclass correlation coefficient (BP-ICC) to WP-ICC, $\alpha_1/\alpha_0 \in [0, 1]$, when both the true correlation model and the working correlation model are nested exchangeable (NEX). Design factors considered are as follows: number of clusters $I = 96$, number of periods $J = 5$, mean cluster-period sizes $\overline{n} \in \{50, 300\}$, and the degree of between-cluster imbalance is defined by coefficient of variation CV $\in \{0.25, 0.75, 1.25\}$. Within-cluster imbalance (pattern 4: randomly permuted) is introduced.

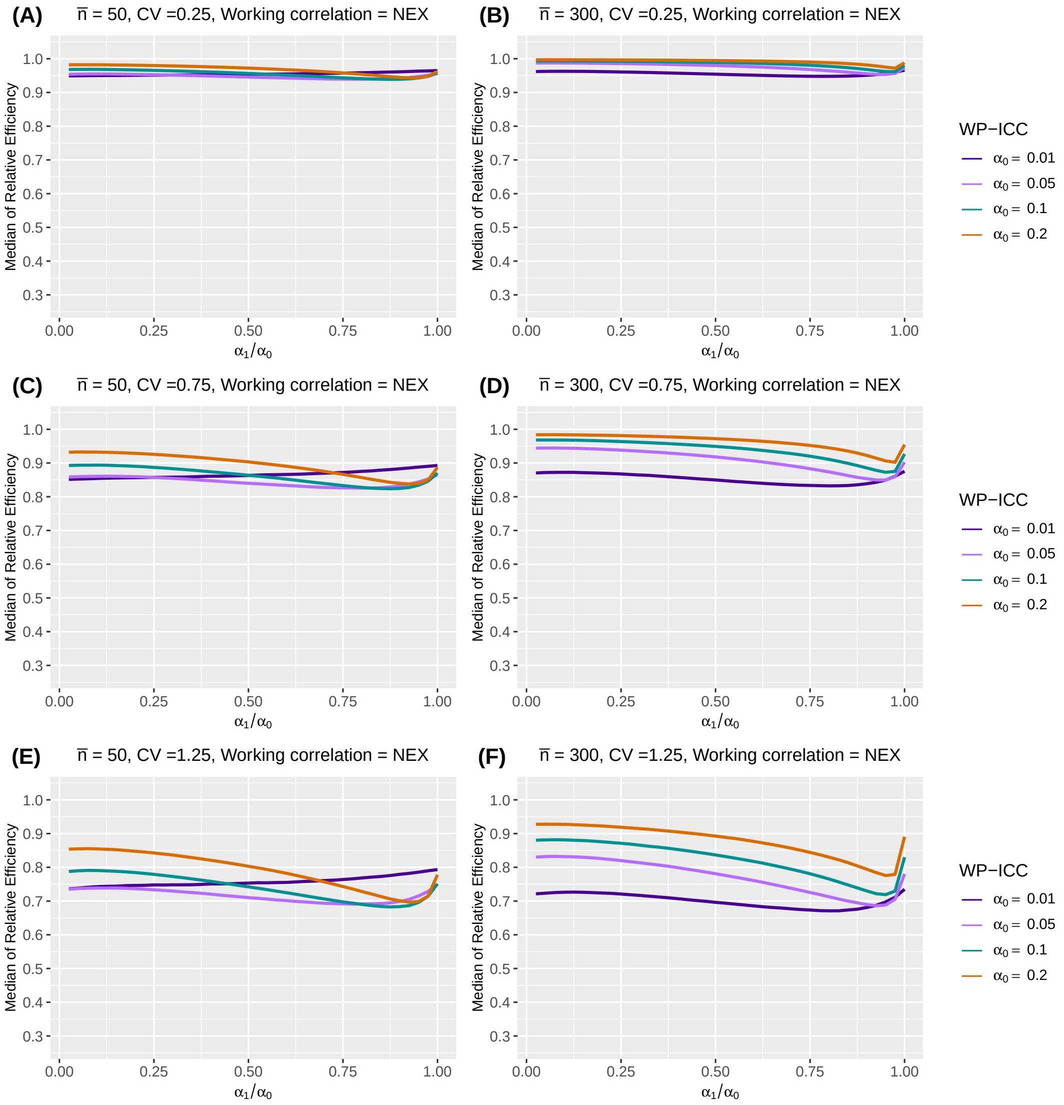

**Web Figure 17** The median of relative efficiency (RE) as a function of the within-period intraclass correlation coefficient (WP-ICC) $\alpha_0 \in \{0.01, 0.05, 0.1, 0.2\}$ and the ratio of between-period intraclass correlation coefficient (BP-ICC) to WP-ICC, $\alpha_1/\alpha_0 \in [0, 1]$, when the true correlation model is nested exchangeable (NEX) and the working correlation model is independence (IND). Design factors considered are as follows: number of clusters $I = 12$, number of periods $J = 5$, mean cluster-period sizes $\overline{n} \in \{50, 300\}$, and the degree of between-cluster imbalance is defined by coefficient of variation CV $\in \{0.25, 0.75, 1.25\}$. No within-cluster imbalance is introduced.

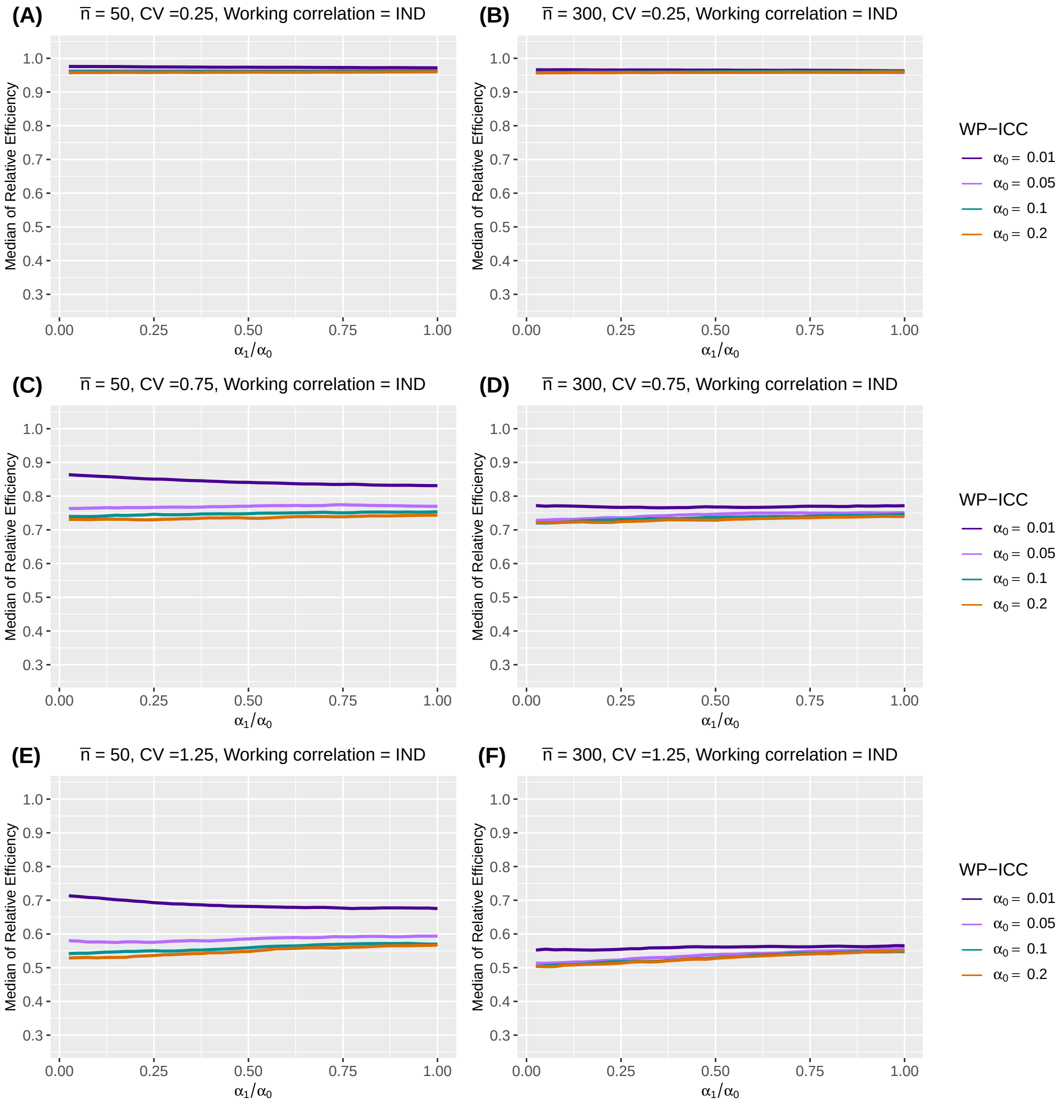

**Web Figure 18** The median of relative efficiency (RE) as a function of the within-period intraclass correlation coefficient (WP-ICC) $\alpha_0 \in \{0.01, 0.05, 0.1, 0.2\}$ and the ratio of between-period intraclass correlation coefficient (BP-ICC) to WP-ICC, $\alpha_1/\alpha_0 \in [0, 1]$, when the true correlation model is nested exchangeable (NEX) and the working correlation model is independence (IND). Design factors considered are as follows: number of clusters $I = 96$, number of periods $J = 5$, mean cluster-period sizes $\overline{n} \in \{50, 300\}$, and the degree of between-cluster imbalance is defined by coefficient of variation CV $\in \{0.25, 0.75, 1.25\}$. No within-cluster imbalance is introduced.

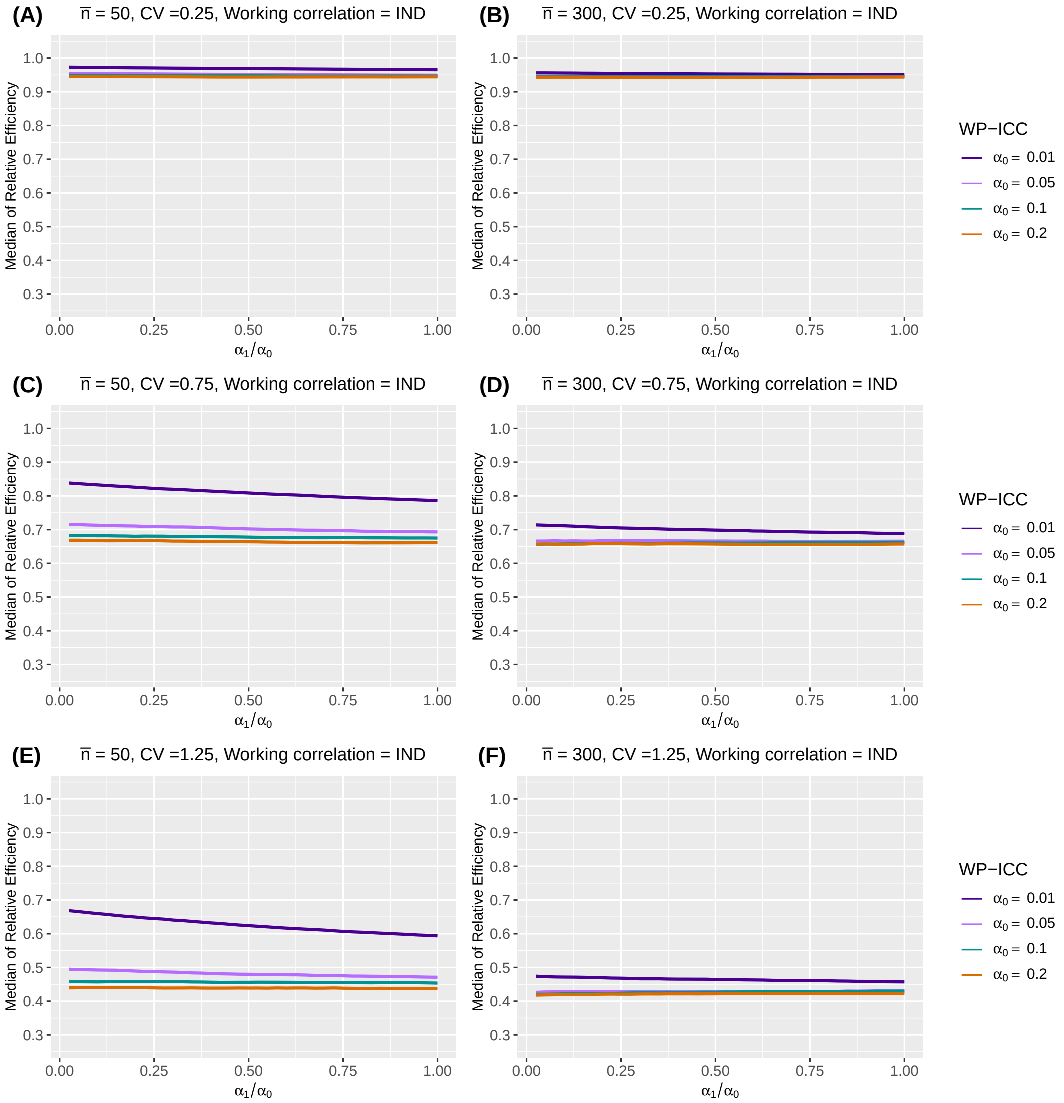

**Web Figure 19** The median of relative efficiency (RE) as a function of the within-period intraclass correlation coefficient (WP-ICC) $\alpha_0 \in \{0.01, 0.05, 0.1, 0.2\}$ and the ratio of between-period intraclass correlation coefficient (BP-ICC) to WP-ICC, $\alpha_1/\alpha_0 \in [0, 1]$, when the true correlation model is nested exchangeable (NEX) and the working correlation model is independence (IND). Design factors considered are as follows: number of clusters $I = 12$, number of periods $J = 5$, mean cluster-period sizes $\overline{n} \in \{50, 300\}$, and the degree of between-cluster imbalance is defined by coefficient of variation CV $\in \{0.25, 0.75, 1.25\}$. Within-cluster imbalance (pattern 4: randomly permuted) is introduced.

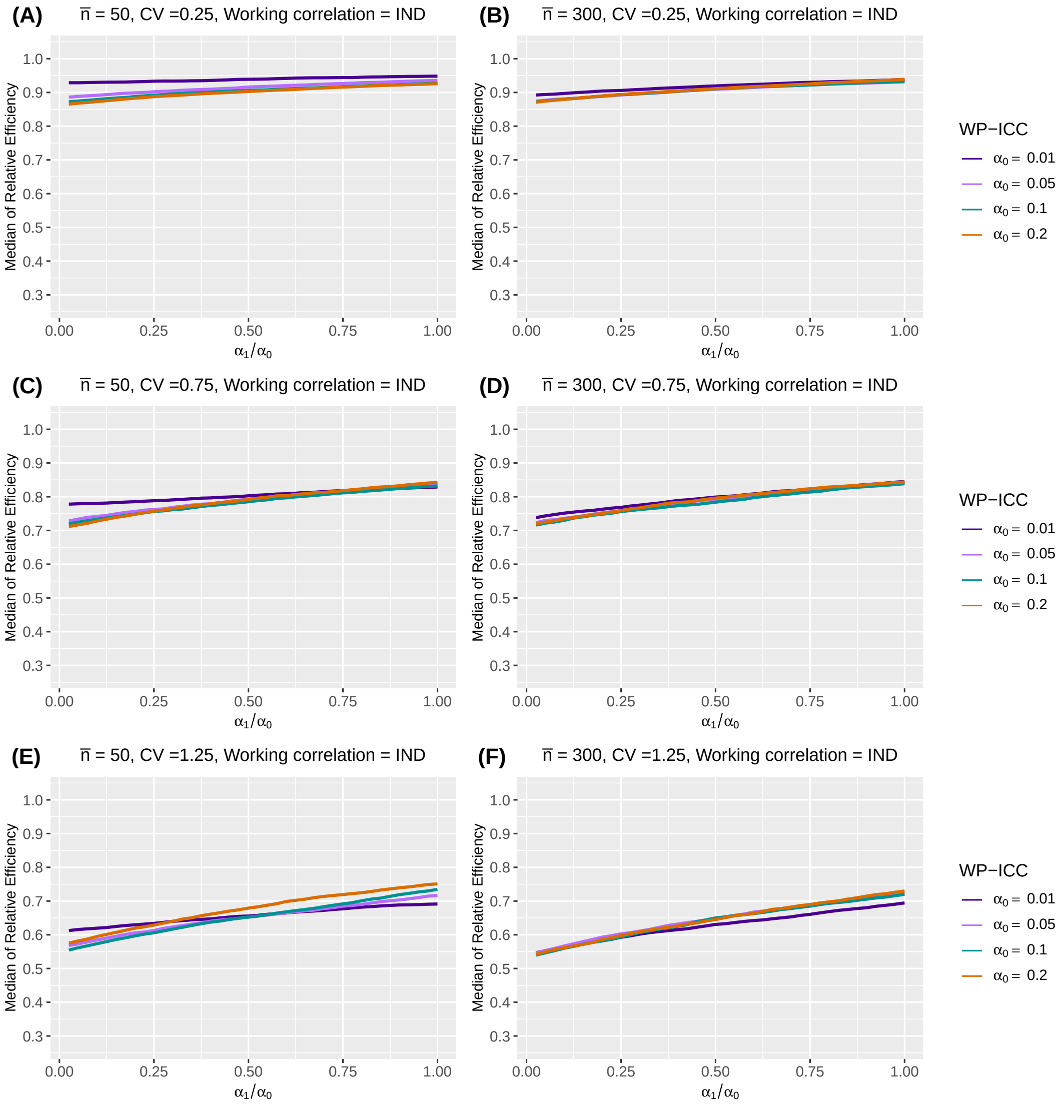

**Web Figure 20** The median of relative efficiency (RE) as a function of the within-period intraclass correlation coefficient (WP-ICC) $\alpha_0 \in \{0.01, 0.05, 0.1, 0.2\}$ and the ratio of between-period intraclass correlation coefficient (BP-ICC) to WP-ICC, $\alpha_1/\alpha_0 \in [0, 1]$, when the true correlation model is nested exchangeable (NEX) and the working correlation model is independence (IND). Design factors considered are as follows: number of clusters $I = 96$, number of periods $J = 5$, mean cluster-period sizes $\overline{n} \in \{50, 300\}$, and the degree of between-cluster imbalance is defined by coefficient of variation CV $\in \{0.25, 0.75, 1.25\}$. Within-cluster imbalance (pattern 4: randomly permuted) is introduced.

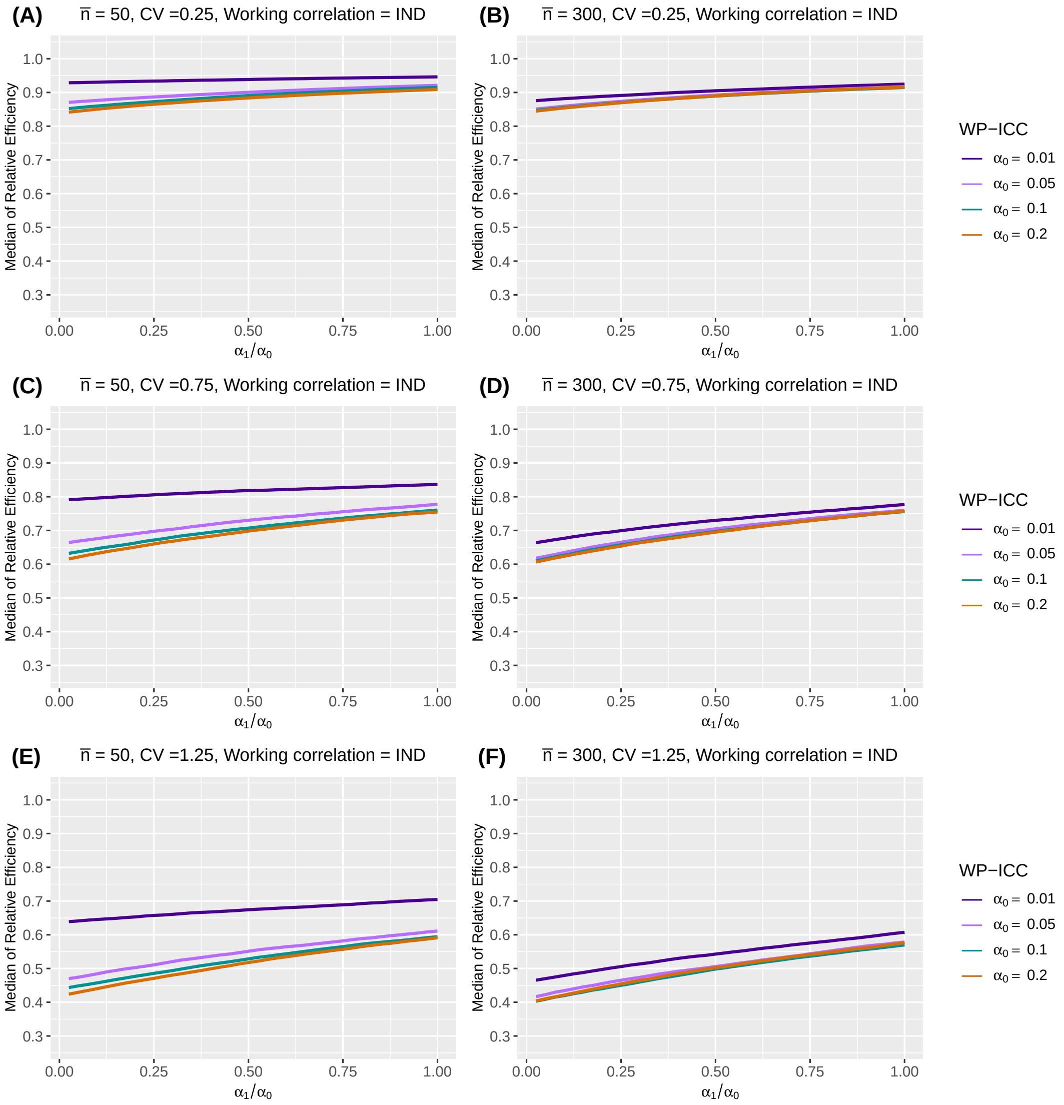

##### Web Appendix C.6. Sensitivity to baseline prevalence, intervention effect and secular trend

As noted in Section 5.6 in the main article, tables below summarize the impact of treatment effect, baseline prevalence and secular trend of the outcomes on RE through a simple sensitivity analysis framework.

**Web Table 4** Median and interquartile range (IQR) (in parentheses) of relative efficiency (RE) as a function of treatment effect $\delta$, baseline prevalence, and different secular trends, when the true correlation model is nested exchangeable and the working correlation correctly specifies the true correlation model. Design factors considered are as follows: number of clusters $I = 24$, number of periods $J = 5$, mean cluster-period size $\overline{n} = 100$. The within-period intraclass correlation coefficient $\alpha_0$ is $0.05$, and the between-period intraclass correlation coefficient $\alpha_1$ is $0.05$. Between cluster imbalance is measured by coefficient of variation, CV $\in \{0.25, 0.75, 1.25\}$. No within-cluster imbalance is introduced.

| $\delta$ | CV | Baseline prevalence | Constant secular trend | Increasing secular trend | Decreasing secular trend |
|---|---|---|---|---|---|
| log(0.35) | 0.25 | 0.1 | 0.995 (0.992, 0.997) | 0.995 (0.991, 0.997) | 0.994 (0.991, 0.998) |
| | | 0.3 | 0.994 (0.990, 0.997) | 0.994 (0.989, 0.998) | 0.994 (0.990, 0.998) |
| | 0.75 | 0.1 | 0.976 (0.963, 0.985) | 0.976 (0.960, 0.984) | 0.974 (0.962, 0.986) |
| | | 0.3 | 0.978 (0.962, 0.988) | 0.977 (0.958, 0.988) | 0.976 (0.961, 0.988) |
| | 1.25 | 0.1 | 0.936 (0.905, 0.954) | 0.933 (0.901, 0.954) | 0.935 (0.902, 0.957) |
| | | 0.3 | 0.943 (0.907, 0.965) | 0.941 (0.903, 0.967) | 0.942 (0.904, 0.967) |
| log(0.75) | 0.25 | 0.1 | 0.993 (0.989, 0.998) | 0.994 (0.988, 0.998) | 0.993 (0.989, 0.998) |
| | | 0.3 | 0.993 (0.988, 0.998) | 0.993 (0.988, 0.998) | 0.993 (0.989, 0.998) |
| | 0.75 | 0.1 | 0.979 (0.960, 0.992) | 0.978 (0.957, 0.991) | 0.978 (0.960, 0.992) |
| | | 0.3 | 0.979 (0.960, 0.993) | 0.978 (0.957, 0.992) | 0.978 (0.960, 0.993) |
| | 1.25 | 0.1 | 0.950 (0.910, 0.976) | 0.947 (0.903, 0.977) | 0.951 (0.908, 0.980) |
| | | 0.3 | 0.952 (0.910, 0.978) | 0.948 (0.903, 0.979) | 0.953 (0.909, 0.981) |

**Web Table 5** Median and interquartile range (IQR) (in parentheses) of relative efficiency (RE) as a function of treatment effect $\delta$, baseline prevalence, and different secular trends, when the true correlation model is nested exchangeable and the working correlation correctly specifies the true correlation model. Design factors considered are as follows: number of clusters $I = 24$, number of periods $J = 5$, mean cluster-period size $\overline{n} = 100$. The within-period intraclass correlation coefficient $\alpha_0$ is $0.05$, and the between-period intraclass correlation coefficient $\alpha_1$ is $0.025$. Between cluster imbalance is measured by coefficient of variation, CV $\in \{0.25, 0.75, 1.25\}$. No within-cluster imbalance is introduced.

| $\delta$ | CV | Baseline prevalence | Constant secular trend | Increasing secular trend | Decreasing secular trend |
|---|---|---|---|---|---|
| log(0.35) | 0.25 | 0.1 | 0.988 (0.986, 0.991) | 0.989 (0.986, 0.991) | 0.989 (0.986, 0.991) |
| | | 0.3 | 0.988 (0.986, 0.991) | 0.988 (0.986, 0.991) | 0.988 (0.985, 0.991) |
| | 0.75 | 0.1 | 0.905 (0.885, 0.922) | 0.900 (0.882, 0.920) | 0.902 (0.883, 0.919) |
| | | 0.3 | 0.903 (0.882, 0.920) | 0.898 (0.879, 0.918) | 0.900 (0.881, 0.917) |
| | 1.25 | 0.1 | 0.768 (0.735, 0.801) | 0.767 (0.731, 0.798) | 0.769 (0.733, 0.803) |
| | | 0.3 | 0.765 (0.730, 0.798) | 0.764 (0.727, 0.795) | 0.765 (0.729, 0.799) |
| log(0.75) | 0.25 | 0.1 | 0.988 (0.985, 0.991) | 0.988 (0.985, 0.990) | 0.988 (0.985, 0.990) |
| | | 0.3 | 0.988 (0.985, 0.991) | 0.988 (0.985, 0.990) | 0.988 (0.985, 0.990) |
| | 0.75 | 0.1 | 0.901 (0.880, 0.919) | 0.897 (0.878, 0.917) | 0.898 (0.879, 0.916) |
| | | 0.3 | 0.901 (0.880, 0.919) | 0.896 (0.878, 0.917) | 0.898 (0.878, 0.916) |
| | 1.25 | 0.1 | 0.763 (0.727, 0.795) | 0.761 (0.725, 0.792) | 0.763 (0.727, 0.796) |
| | | 0.3 | 0.763 (0.727, 0.795) | 0.760 (0.725, 0.792) | 0.763 (0.726, 0.796) |

**Web Table 6** Median and interquartile range (IQR) (in parentheses) of relative efficiency (RE) as a function of treatment effect $\delta$, baseline prevalence, and different secular trends, when the true correlation model is nested exchangeable (NEX) and the working correlation is independence (IND). Design factors considered are as follows: number of clusters $I = 24$, number of periods $J = 5$, mean cluster-period size $\overline{n} = 100$. The within-period intraclass correlation coefficient $\alpha_0$ is $0.05$, and the between-period intraclass correlation coefficient $\alpha_1$ is $0.05$. Between cluster imbalance is measured by coefficient of variation, CV $\in \{0.25, 0.75, 1.25\}$. No within-cluster imbalance is introduced.

| $\delta$ | CV | Baseline prevalence | Constant secular trend | Increasing secular trend | Decreasing secular trend |
|---|---|---|---|---|---|
| log(0.35) | 0.25 | 0.1 | 0.954 (0.929, 0.979) | 0.954 (0.925, 0.979) | 0.954 (0.927, 0.980) |
| | | 0.3 | 0.954 (0.929, 0.979) | 0.954 (0.925, 0.979) | 0.955 (0.927, 0.979) |
| | 0.75 | 0.1 | 0.723 (0.642, 0.793) | 0.714 (0.640, 0.782) | 0.721 (0.642, 0.786) |
| | | 0.3 | 0.722 (0.639, 0.794) | 0.714 (0.639, 0.784) | 0.724 (0.641, 0.785) |
| | 1.25 | 0.1 | 0.503 (0.419, 0.586) | 0.501 (0.421, 0.579) | 0.503 (0.426, 0.591) |
| | | 0.3 | 0.504 (0.420, 0.586) | 0.503 (0.421, 0.582) | 0.506 (0.424, 0.592) |
| log(0.75) | 0.25 | 0.1 | 0.954 (0.928, 0.979) | 0.953 (0.926, 0.979) | 0.954 (0.928, 0.979) |
| | | 0.3 | 0.954 (0.927, 0.978) | 0.953 (0.926, 0.979) | 0.954 (0.928, 0.979) |
| | 0.75 | 0.1 | 0.719 (0.639, 0.795) | 0.717 (0.638, 0.786) | 0.723 (0.641, 0.787) |
| | | 0.3 | 0.717 (0.639, 0.794) | 0.719 (0.639, 0.788) | 0.722 (0.641, 0.788) |
| | 1.25 | 0.1 | 0.505 (0.424, 0.586) | 0.502 (0.424, 0.587) | 0.506 (0.424, 0.593) |
| | | 0.3 | 0.503 (0.424, 0.588) | 0.500 (0.425, 0.588) | 0.508 (0.422, 0.590) |

**Web Table 7** Median and interquartile range (IQR) (in parentheses) of relative efficiency (RE) as a function of treatment effect $\delta$, baseline prevalence, and different secular trends, when the true correlation model is nested exchangeable (NEX) and the working correlation is independence (IND). Design factors considered are as follows: number of clusters $I = 24$, number of periods $J = 5$, mean cluster-period size $\overline{n} = 100$. The within-period intraclass correlation coefficient $\alpha_0$ is $0.05$, and the between-period intraclass correlation coefficient $\alpha_1$ is $0.025$. Between cluster imbalance is measured by coefficient of variation, CV $\in \{0.25, 0.75, 1.25\}$. No within-cluster imbalance is introduced.

| $\delta$ | CV | Baseline prevalence | Constant secular trend | Increasing secular trend | Decreasing secular trend |
|---|---|---|---|---|---|
| log(0.35) | 0.25 | 0.1 | 0.954 (0.937, 0.971) | 0.955 (0.935, 0.971) | 0.954 (0.936, 0.972) |
| | | 0.3 | 0.954 (0.937, 0.972) | 0.955 (0.935, 0.971) | 0.954 (0.936, 0.972) |
| | 0.75 | 0.1 | 0.723 (0.658, 0.775) | 0.712 (0.654, 0.770) | 0.718 (0.659, 0.771) |
| | | 0.3 | 0.722 (0.656, 0.776) | 0.713 (0.653, 0.773) | 0.718 (0.658, 0.770) |
| | 1.25 | 0.1 | 0.501 (0.428, 0.565) | 0.498 (0.428, 0.564) | 0.501 (0.434, 0.576) |
| | | 0.3 | 0.502 (0.430, 0.564) | 0.501 (0.432, 0.563) | 0.501 (0.435, 0.576) |
| log(0.75) | 0.25 | 0.1 | 0.954 (0.936, 0.972) | 0.954 (0.935, 0.971) | 0.953 (0.935, 0.972) |
| | | 0.3 | 0.954 (0.936, 0.971) | 0.955 (0.935, 0.971) | 0.953 (0.935, 0.972) |
| | 0.75 | 0.1 | 0.720 (0.654, 0.778) | 0.714 (0.654, 0.773) | 0.719 (0.656, 0.772) |
| | | 0.3 | 0.720 (0.654, 0.778) | 0.714 (0.654, 0.774) | 0.718 (0.655, 0.771) |
| | 1.25 | 0.1 | 0.503 (0.429, 0.564) | 0.500 (0.432, 0.566) | 0.501 (0.432, 0.573) |
| | | 0.3 | 0.502 (0.431, 0.563) | 0.500 (0.430, 0.567) | 0.502 (0.432, 0.572) |

**Web Table 8** Median and interquartile range (IQR) (in parentheses) of relative efficiency (RE) as a function of treatment effect $\delta$, baseline prevalence, and different secular trends, when the true correlation model is nested exchangeable (NEX) and the working correlation correctly specifies the true one. Design factors considered are as follows: number of clusters $I = 24$, number of periods $J = 5$, mean cluster-period size $\overline{n} = 100$. The within-period intraclass correlation coefficient $\alpha_0$ is $0.05$, and the between-period intraclass correlation coefficient $\alpha_1$ is $0.05$. Between cluster imbalance is measured by coefficient of variation, CV $\in \{0.25, 0.75, 1.25\}$. Within-cluster imbalance (pattern 4: randomly permuted) is introduced.

| $\delta$ | CV | Baseline prevalence | Constant secular trend | Increasing secular trend | Decreasing secular trend |
|---|---|---|---|---|---|
| log(0.35) | 0.25 | 0.1 | 0.963 (0.942, 0.984) | 0.961 (0.941, 0.984) | 0.963 (0.944, 0.984) |
| | | 0.3 | 0.956 (0.931, 0.980) | 0.954 (0.930, 0.98) | 0.958 (0.935, 0.981) |
| | 0.75 | 0.1 | 0.860 (0.793, 0.917) | 0.856 (0.796, 0.913) | 0.857 (0.798, 0.920) |
| | | 0.3 | 0.837 (0.765, 0.903) | 0.833 (0.764, 0.896) | 0.837 (0.771, 0.908) |
| | 1.25 | 0.1 | 0.695 (0.604, 0.785) | 0.688 (0.601, 0.784) | 0.700 (0.613, 0.791) |
| | | 0.3 | 0.659 (0.562, 0.753) | 0.647 (0.554, 0.749) | 0.666 (0.575, 0.764) |
| log(0.75) | 0.25 | 0.1 | 0.950 (0.922, 0.977) | 0.948 (0.921, 0.977) | 0.951 (0.923, 0.977) |
| | | 0.3 | 0.949 (0.921, 0.977) | 0.947 (0.919, 0.978) | 0.950 (0.921, 0.976) |
| | 0.75 | 0.1 | 0.816 (0.741, 0.890) | 0.813 (0.740, 0.887) | 0.814 (0.738, 0.889) |
| | | 0.3 | 0.813 (0.739, 0.888) | 0.812 (0.737, 0.886) | 0.811 (0.736, 0.887) |
| | 1.25 | 0.1 | 0.625 (0.524, 0.724) | 0.621 (0.522, 0.721) | 0.629 (0.532, 0.733) |
| | | 0.3 | 0.622 (0.519, 0.719) | 0.618 (0.518, 0.718) | 0.625 (0.528, 0.729) |

**Web Table 9** Median and interquartile range (IQR) (in parentheses) of relative efficiency (RE) as a function of treatment effect $\delta$, baseline prevalence, and different secular trends, when the true correlation model is nested exchangeable (NEX) and the working correlation correctly specifies the true one. Design factors considered are as follows: number of clusters $I = 24$, number of periods $J = 5$, mean cluster-period size $\overline{n} = 100$. The within-period intraclass correlation coefficient $\alpha_0$ is $0.05$, and the between-period intraclass correlation coefficient $\alpha_1$ is $0.025$. Between cluster imbalance is measured by coefficient of variation, CV $\in \{0.25, 0.75, 1.25\}$. Within-cluster imbalance (pattern 4: randomly permuted) is introduced.

| $\delta$ | CV | Baseline prevalence | Constant secular trend | Increasing secular trend | Decreasing secular trend |
|---|---|---|---|---|---|
| log(0.35) | 0.25 | 0.1 | 0.960 (0.950, 0.969) | 0.960 (0.950, 0.969) | 0.960 (0.951, 0.969) |
| | | 0.3 | 0.959 (0.949, 0.969) | 0.959 (0.949, 0.968) | 0.959 (0.949, 0.968) |
| | 0.75 | 0.1 | 0.868 (0.825, 0.905) | 0.869 (0.828, 0.902) | 0.868 (0.825, 0.904) |
| | | 0.3 | 0.863 (0.822, 0.902) | 0.865 (0.823, 0.899) | 0.864 (0.819, 0.900) |
| | 1.25 | 0.1 | 0.712 (0.646, 0.772) | 0.710 (0.645, 0.772) | 0.715 (0.653, 0.776) |
| | | 0.3 | 0.708 (0.638, 0.768) | 0.704 (0.639, 0.764) | 0.710 (0.646, 0.770) |
| log(0.75) | 0.25 | 0.1 | 0.959 (0.948, 0.968) | 0.958 (0.948, 0.968) | 0.958 (0.948, 0.967) |
| | | 0.3 | 0.958 (0.948, 0.968) | 0.958 (0.948, 0.968) | 0.958 (0.948, 0.967) |
| | 0.75 | 0.1 | 0.860 (0.818, 0.900) | 0.863 (0.820, 0.898) | 0.862 (0.814, 0.898) |
| | | 0.3 | 0.860 (0.819, 0.899) | 0.862 (0.820, 0.897) | 0.861 (0.814, 0.897) |
| | 1.25 | 0.1 | 0.704 (0.630, 0.762) | 0.699 (0.636, 0.761) | 0.706 (0.641, 0.767) |
| | | 0.3 | 0.704 (0.631, 0.762) | 0.698 (0.635, 0.759) | 0.706 (0.640, 0.767) |

**Web Table 10** Median and interquartile range (IQR) (in parentheses) of relative efficiency (RE) as a function of treatment effect $\delta$, baseline prevalence, and different secular trends, when the true correlation model is nested exchangeable (NEX) and the working correlation is independence (IND). Design factors considered are as follows: number of clusters $I = 24$, number of periods $J = 5$, mean cluster-period size $\overline{n} = 100$. The within-period intraclass correlation coefficient $\alpha_0$ is $0.05$, and the between-period intraclass correlation coefficient $\alpha_1$ is $0.05$. Between cluster imbalance is measured by coefficient of variation, CV $\in \{0.25, 0.75, 1.25\}$. Within-cluster imbalance (pattern 4: randomly permuted) is introduced.

| $\delta$ | CV | Baseline prevalence | Constant secular trend | Increasing secular trend | Decreasing secular trend |
|---|---|---|---|---|---|
| log(0.35) | 0.25 | 0.1 | 0.929 (0.894, 0.960) | 0.923 (0.893, 0.958) | 0.928 (0.892, 0.966) |
| | | 0.3 | 0.928 (0.893, 0.961) | 0.923 (0.892, 0.957) | 0.927 (0.892, 0.966) |
| | 0.75 | 0.1 | 0.807 (0.739, 0.893) | 0.800 (0.730, 0.886) | 0.811 (0.739, 0.893) |
| | | 0.3 | 0.806 (0.735, 0.890) | 0.800 (0.730, 0.886) | 0.811 (0.737, 0.894) |
| | 1.25 | 0.1 | 0.657 (0.563, 0.758) | 0.661 (0.574, 0.766) | 0.671 (0.565, 0.767) |
| | | 0.3 | 0.658 (0.562, 0.761) | 0.663 (0.573, 0.764) | 0.668 (0.571, 0.769) |
| log(0.75) | 0.25 | 0.1 | 0.928 (0.892, 0.961) | 0.925 (0.891, 0.957) | 0.927 (0.892, 0.968) |
| | | 0.3 | 0.928 (0.892, 0.960) | 0.924 (0.891, 0.956) | 0.927 (0.892, 0.967) |
| | 0.75 | 0.1 | 0.805 (0.730, 0.889) | 0.801 (0.728, 0.890) | 0.809 (0.730, 0.894) |
| | | 0.3 | 0.806 (0.730, 0.889) | 0.801 (0.729, 0.890) | 0.810 (0.731, 0.894) |
| | 1.25 | 0.1 | 0.657 (0.562, 0.762) | 0.662 (0.575, 0.762) | 0.665 (0.571, 0.767) |
| | | 0.3 | 0.657 (0.563, 0.764) | 0.662 (0.575, 0.761) | 0.664 (0.570, 0.765) |

**Web Table 11** Median and interquartile range (IQR) (in parentheses) of relative efficiency (RE) as a function of treatment effect $\delta$, baseline prevalence, and different secular trends, when the true correlation model is nested exchangeable (NEX) and the working correlation is independence (IND). Design factors considered are as follows: number of clusters $I = 24$, number of periods $J = 5$, mean cluster-period size $\overline{n} = 100$. The within-period intraclass correlation coefficient $\alpha_0$ is $0.05$, and the between-period intraclass correlation coefficient $\alpha_1$ is $0.025$. Between cluster imbalance is measured by coefficient of variation, CV $\in \{0.25, 0.75, 1.25\}$. Within-cluster imbalance (pattern 4: randomly permuted) is introduced.

| $\delta$ | CV | Baseline prevalence | Constant secular trend | Increasing secular trend | Decreasing secular trend |
|---|---|---|---|---|---|
| log(0.35) | 0.25 | 0.1 | 0.907 (0.883, 0.927) | 0.902 (0.881, 0.926) | 0.904 (0.881, 0.930) |
| | | 0.3 | 0.906 (0.882, 0.927) | 0.902 (0.882, 0.926) | 0.904 (0.881, 0.930) |
| | 0.75 | 0.1 | 0.761 (0.695, 0.819) | 0.755 (0.693, 0.816) | 0.765 (0.701, 0.823) |
| | | 0.3 | 0.760 (0.696, 0.820) | 0.755 (0.693, 0.819) | 0.764 (0.697, 0.823) |
| | 1.25 | 0.1 | 0.589 (0.502, 0.675) | 0.595 (0.516, 0.675) | 0.600 (0.519, 0.680) |
| | | 0.3 | 0.590 (0.503, 0.673) | 0.599 (0.515, 0.676) | 0.597 (0.517, 0.683) |
| log(0.75) | 0.25 | 0.1 | 0.906 (0.882, 0.927) | 0.902 (0.881, 0.926) | 0.904 (0.880, 0.930) |
| | | 0.3 | 0.906 (0.881, 0.927) | 0.902 (0.881, 0.925) | 0.905 (0.879, 0.930) |
| | 0.75 | 0.1 | 0.758 (0.697, 0.819) | 0.755 (0.693, 0.820) | 0.764 (0.696, 0.823) |
| | | 0.3 | 0.758 (0.696, 0.821) | 0.756 (0.694, 0.819) | 0.764 (0.695, 0.824) |
| | 1.25 | 0.1 | 0.594 (0.505, 0.672) | 0.596 (0.512, 0.677) | 0.595 (0.518, 0.682) |
| | | 0.3 | 0.594 (0.504, 0.674) | 0.596 (0.511, 0.678) | 0.596 (0.516, 0.680) |

#### Web Appendix D. Supplementary simulation results under exponential decay true correlation structure

Simulation results under exponential decay true correlation model are parallelly showed in this section. As noted in Section 6 in the main article, all $\alpha_0/\alpha_1$ or $\alpha_1$ expressions would be substituted by the decay rate $\rho$ specified in the exponential decay correlation structure.

##### Web Appendix D.1. Cluter size variability and number of clusters

The following figures show the counterparts to all RE versus CV plots that illustrate the impact of cluster size variability and the number of clusters on RE, but under exponential decay true correlation structure.

**Web Figure 21** The median and interquartile range (IQR) of relative efficiency (RE) as a function of coefficient of variation (CV) measuring between-cluster imbalance, when the true correlation model is exponential decay. Design factors considered are as follows: number of clusters $I = 12$ and $96$, number of periods $J = 5$. The within-period intraclass correlation coefficient (WP-ICC) $\alpha_0 = 0.05$, and decay rate $\rho \in \{0.1, 0.5, 1\}$. No within-cluster variability in cluster-period sizes is introduced.

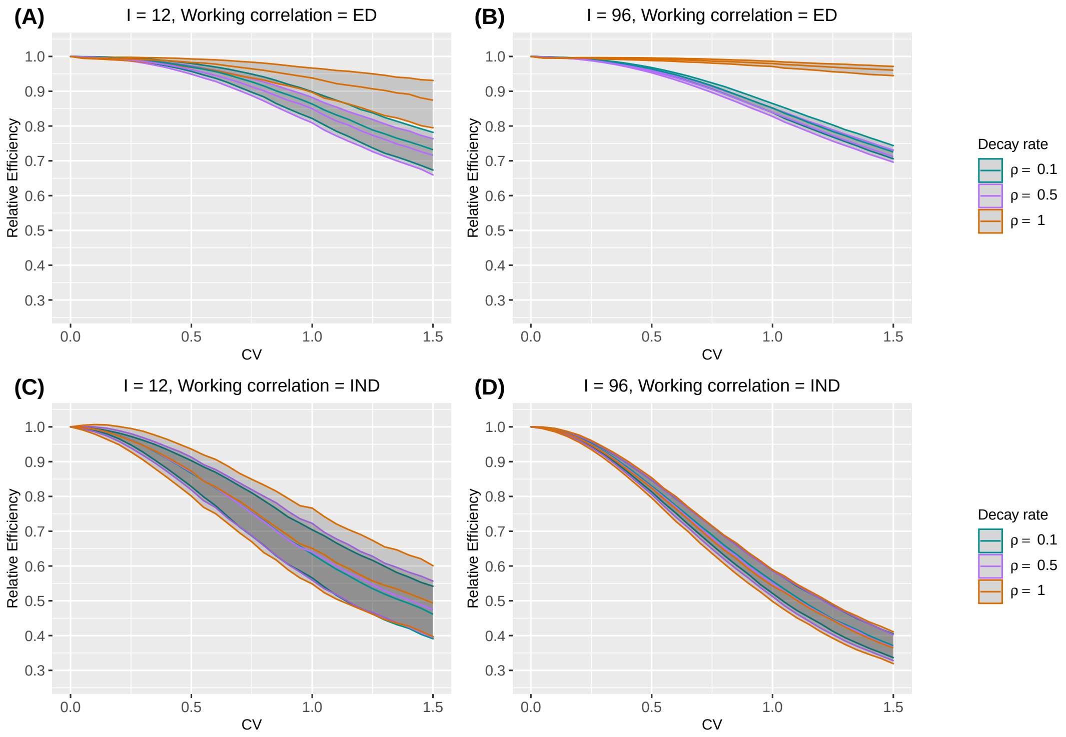

**Web Figure 22** The median and interquartile range (IQR) of relative efficiency (RE) as a function of coefficient of variation (CV) measuring between-cluster imbalance, when the true correlation model is exponential decay. Design factors considered are as follows: number of clusters $I = 24$ and $48$, number of periods $J = 5$. The within-period intraclass correlation coefficient (WP-ICC) $\alpha_0 = 0.05$, and decay rate $\rho \in \{0.1, 0.5, 1\}$. No within-cluster variability in cluster-period sizes is introduced.

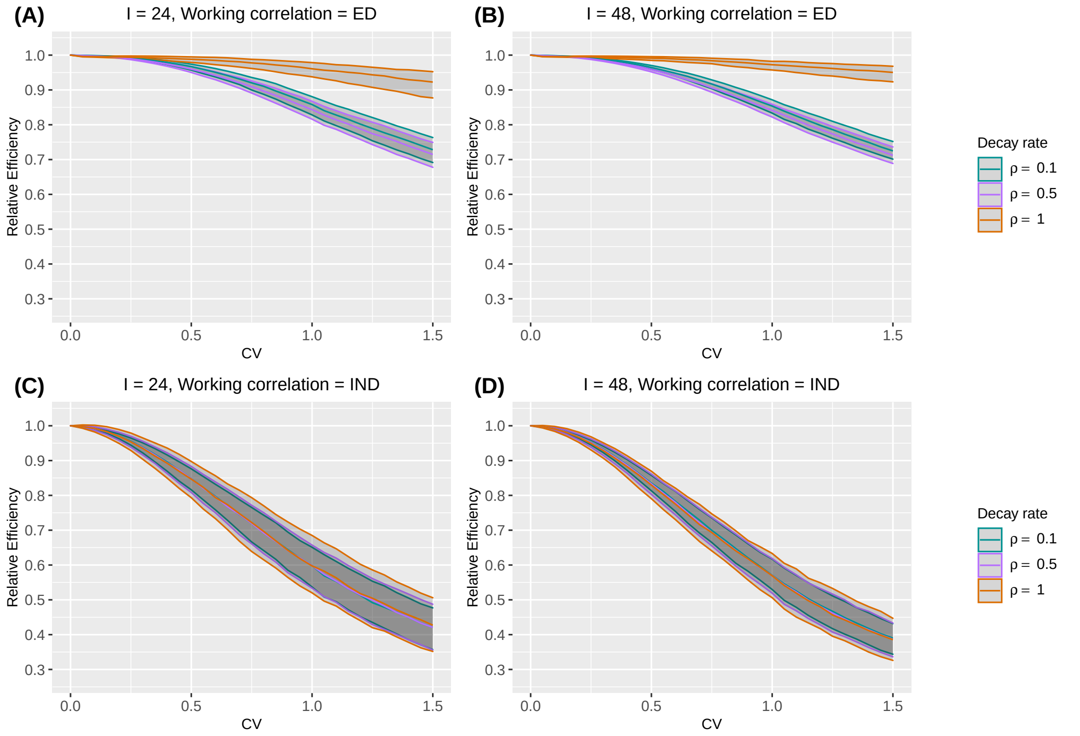

**Web Figure 23** The median and interquartile range (IQR) of relative efficiency (RE) as a function of coefficient of variation (CV) measuring between-cluster imbalance, when the true correlation model is exponential decay. Design factors considered are as follows: number of clusters $I = 12$ and $96$, number of periods $J = 5$. The within-period intraclass correlation coefficient (WP-ICC) $\alpha_0 = 0.05$, and decay rate $\rho \in \{0.1, 0.5, 1\}$. Within-cluster imbalance (pattern 1: constant) is introduced.

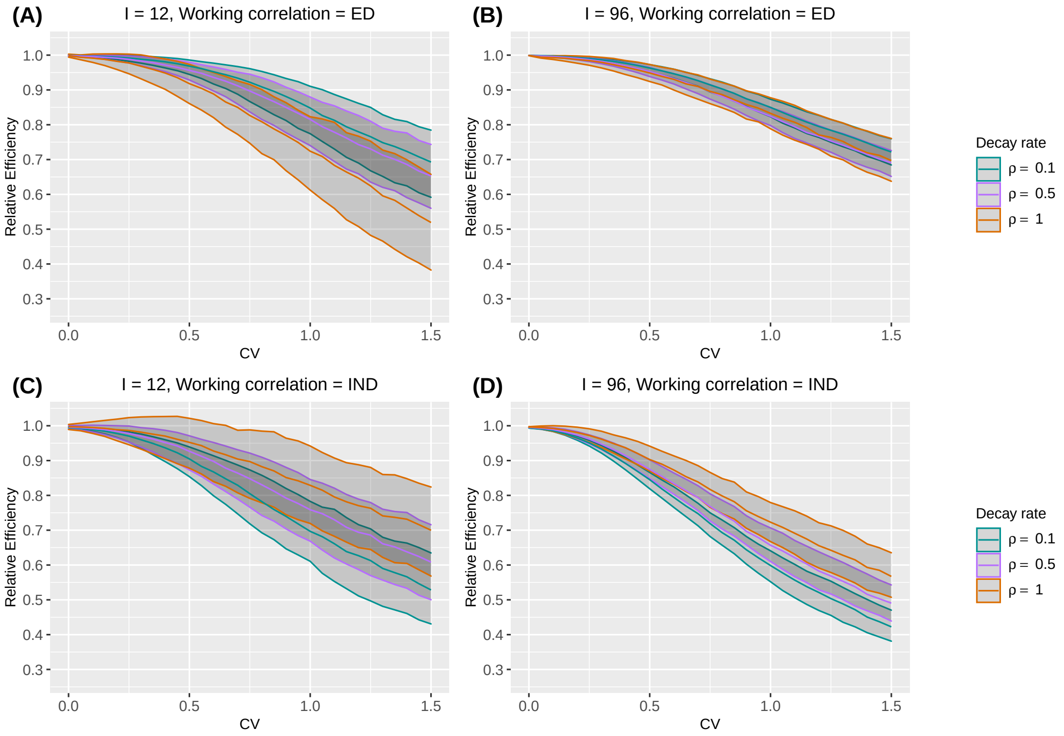

**Web Figure 24** The median and interquartile range (IQR) of relative efficiency (RE) as a function of coefficient of variation (CV) measuring between-cluster imbalance, when the true correlation model is exponential decay. Design factors considered are as follows: number of clusters $I = 12$ and $96$, number of periods $J = 5$. The within-period intraclass correlation coefficient (WP-ICC) $\alpha_0 = 0.05$, and decay rate $\rho \in \{0.1, 0.5, 1\}$. Within-cluster imbalance (pattern 2: monotonically increasing) is introduced.

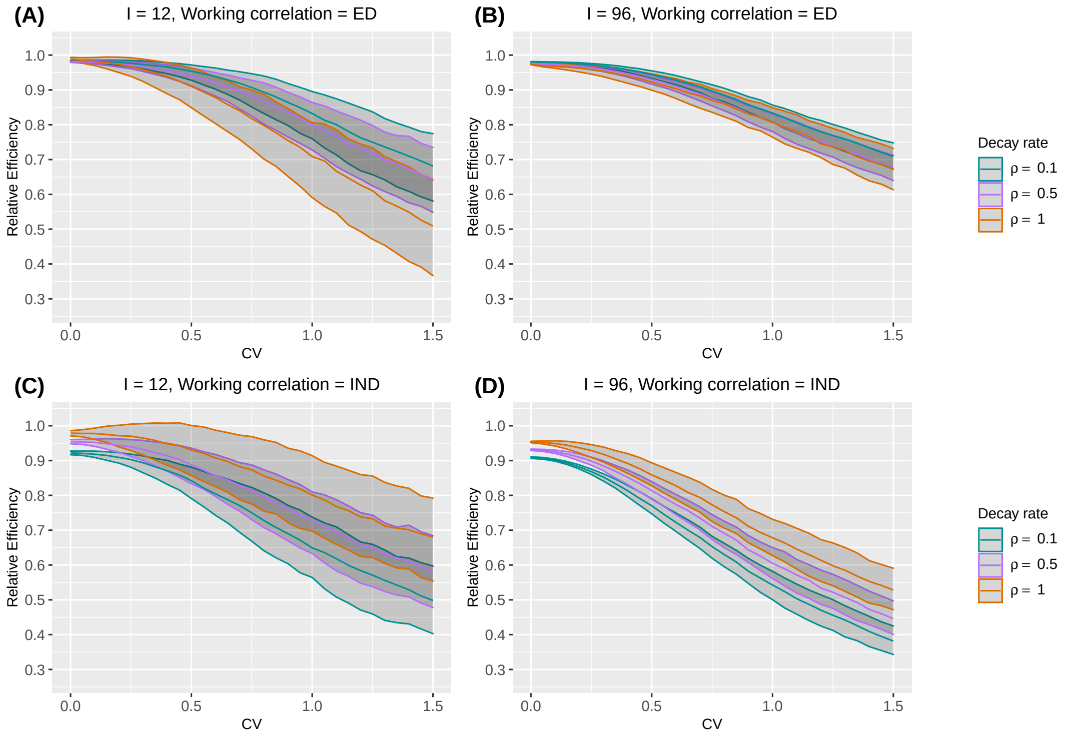

**Web Figure 25** The median and interquartile range (IQR) of relative efficiency (RE) as a function of coefficient of variation (CV) measuring between-cluster imbalance, when the true correlation model is exponential decay. Design factors considered are as follows: number of clusters $I = 12$ and 96, number of periods $J = 5$. The within-period intraclass correlation coefficient (WP-ICC) $\alpha_0 = 0.05$, and decay rate $\rho \in \{0.1, 0.5, 1\}$. Within-cluster imbalance (pattern 3: monotonically decreasing) is introduced.

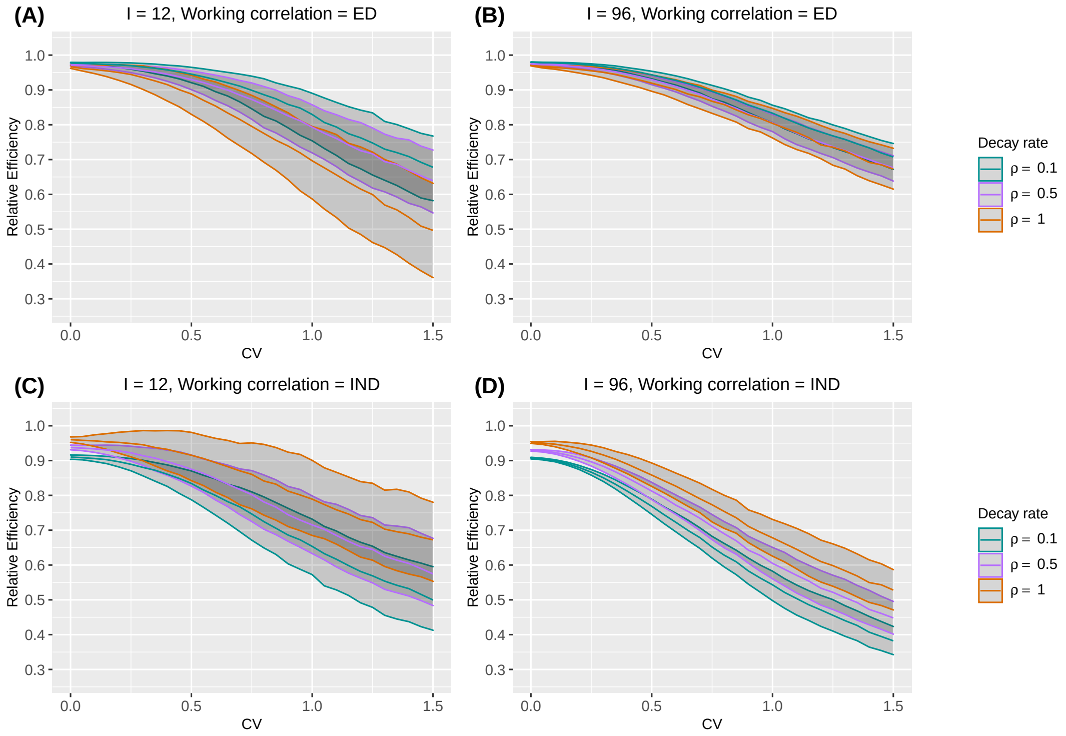

**Web Figure 26** The median and interquartile range (IQR) of relative efficiency (RE) as a function of coefficient of variation (CV) measuring between-cluster imbalance, when the true correlation model is exponential decay. Design factors considered are as follows: number of clusters $I = 12$ and 96, number of periods $J = 5$. The within-period intraclass correlation coefficient (WP-ICC) $\alpha_0 = 0.05$, and decay rate $\rho \in \{0.1, 0.5, 1\}$. Within-cluster imbalance (pattern 4: randomly permuted) is introduced.

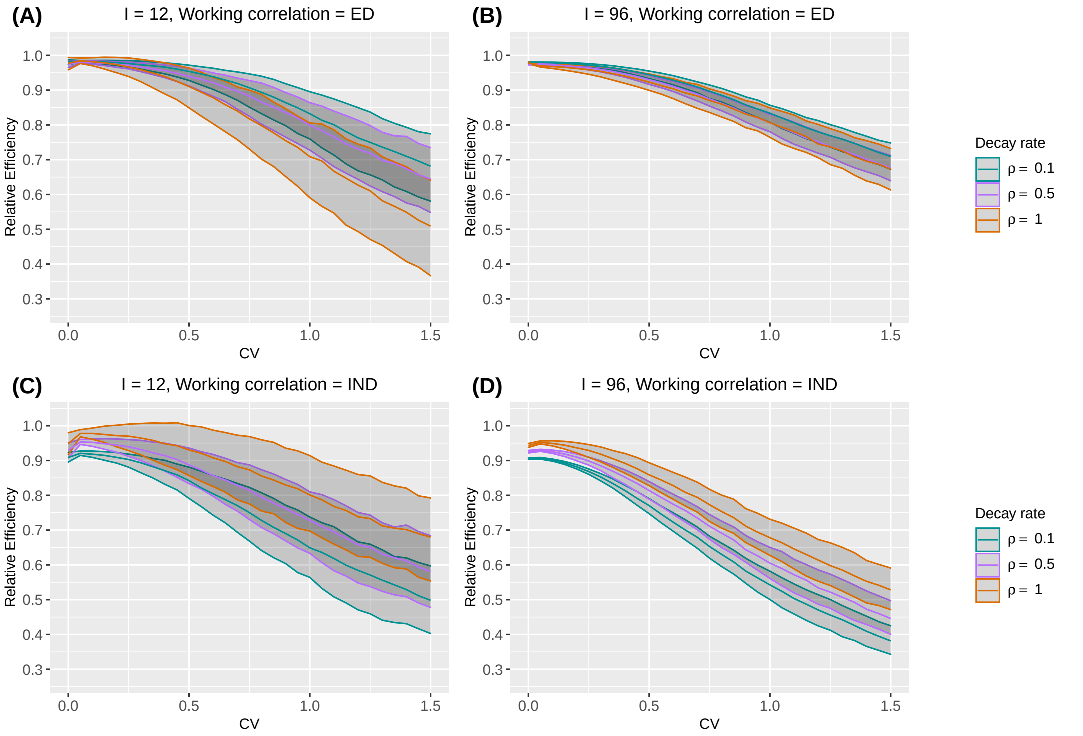

#### Web Appendix D.2. Intraclass correlation coefficients

Figures in this section show the counterparts to plots that illustrate the impact of ICC parameters on RE, but under exponential decay true correlation structure.

**Web Figure 27** The median of relative efficiency (RE) as a function of within-period intraclass correlation coefficient (WP-ICC) $\alpha_0 \in \{0.01, 0.05, 0.1, 0.2\}$ and the decay rate $\rho \in (0, 1)$, when both the true correlation model and the working correlation model are exponential decay (ED). Design factors considered are as follows: number of clusters $I = 12$ and 96, number of periods $J = 5$, mean cluster-period size $\overline{n} = 100$, and the degree of between-cluster imbalance is defined by coefficient of variation CV $\in \{0.25, 0.75, 1.25\}$. No within-cluster imbalance is introduced.

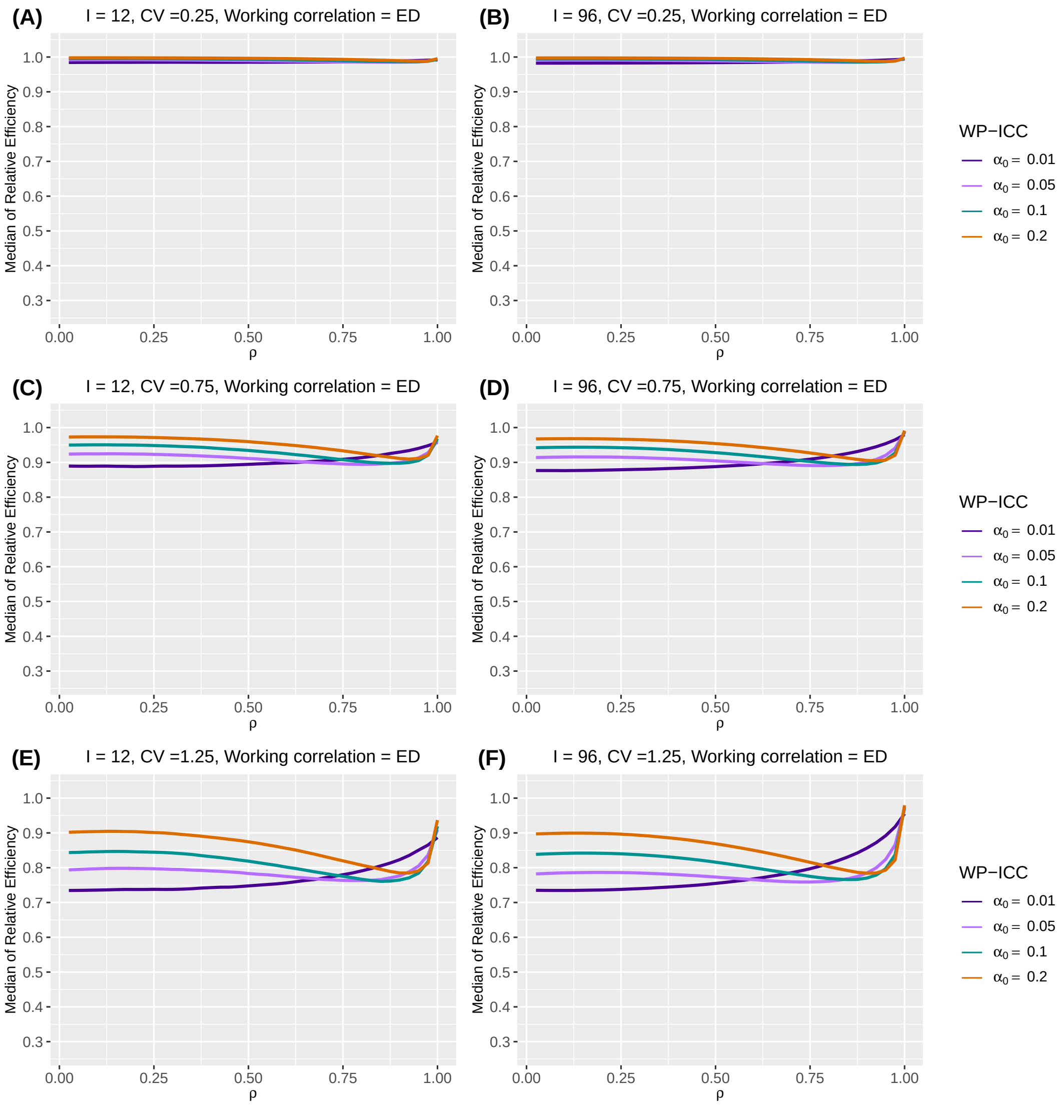

**Web Figure 28** The median of relative efficiency (RE) as a function of within-period intraclass correlation coefficient (WP-ICC) $\alpha_0 \in \{0.01, 0.05, 0.1, 0.2\}$ and the decay rate $\rho \in (0, 1)$, when both the true correlation model and the working correlation model are exponential decay (ED). Design factors considered are as follows: number of clusters $I = 12$ and 96, number of periods $J = 5$, mean cluster-period size $\overline{n} = 100$, and the degree of between-cluster imbalance is defined by coefficient of variation CV $\in \{0.25, 0.75, 1.25\}$. Within-cluster imbalance (pattern 1: constant) is introduced.

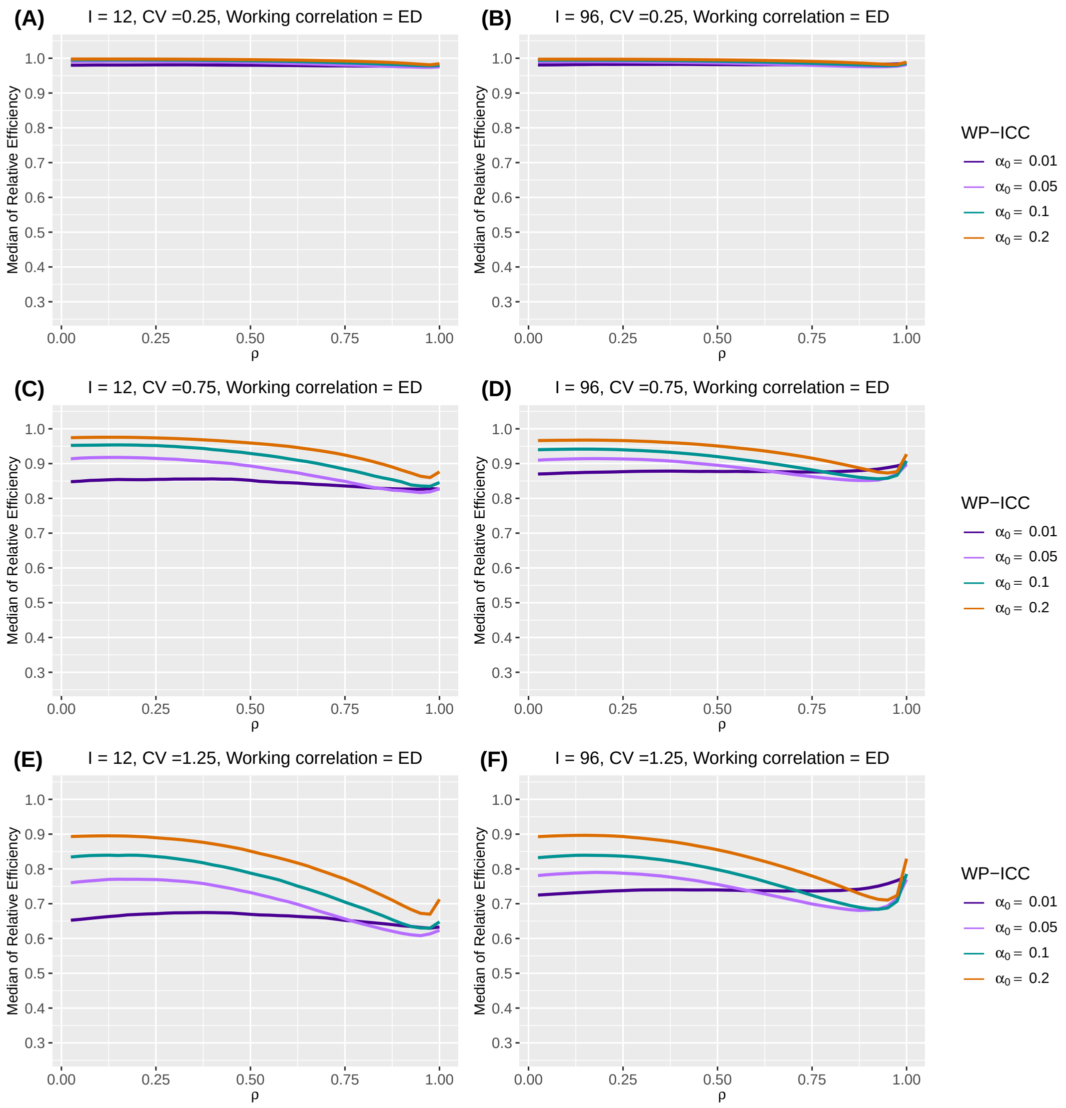

**Web Figure 29** The median of relative efficiency (RE) as a function of within-period intraclass correlation coefficient (WP-ICC) $\alpha_0 \in \{0.01, 0.05, 0.1, 0.2\}$ and the decay rate $\rho \in (0, 1)$, when both the true correlation model and the working correlation model are exponential decay (ED). Design factors considered are as follows: number of clusters $I = 12$ and 96, number of periods $J = 5$, mean cluster-period size $\overline{n} = 100$, and the degree of between-cluster imbalance is defined by coefficient of variation CV $\in \{0.25, 0.75, 1.25\}$. Within-cluster imbalance (pattern 2: monotonically increasing) is introduced.

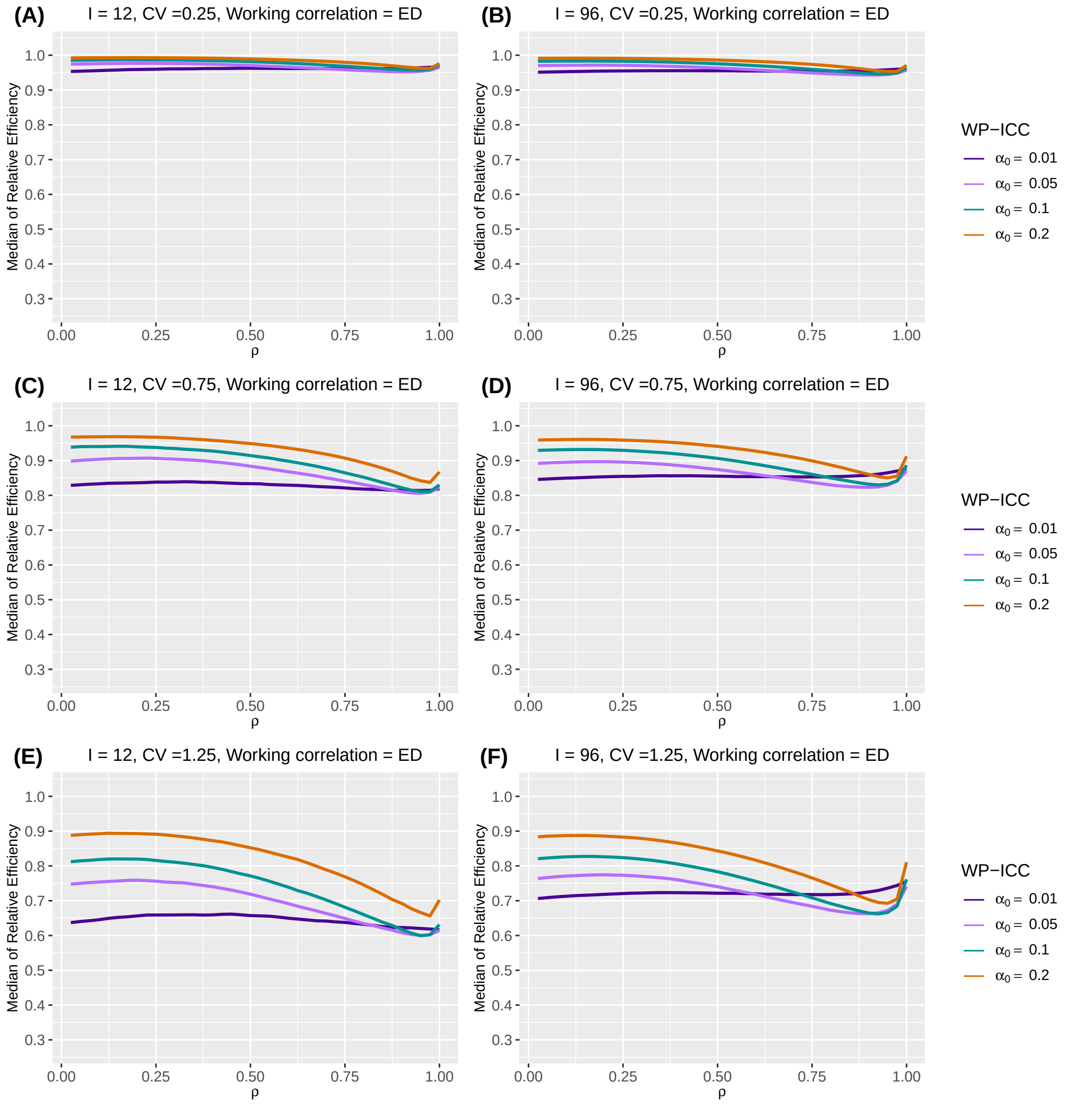

**Web Figure 30** The median of relative efficiency (RE) as a function of within-period intraclass correlation coefficient (WP-ICC) $\alpha_0 \in \{0.01, 0.05, 0.1, 0.2\}$ and the decay rate $\rho \in (0, 1)$, when both the true correlation model and the working correlation model are exponential decay (ED). Design factors considered are as follows: number of clusters $I = 12$ and 96, number of periods $J = 5$, mean cluster-period size $\overline{n} = 100$, and the degree of between-cluster imbalance is defined by coefficient of variation CV $\in \{0.25, 0.75, 1.25\}$. Within-cluster imbalance (pattern 3: monotonically decreasing) is introduced.

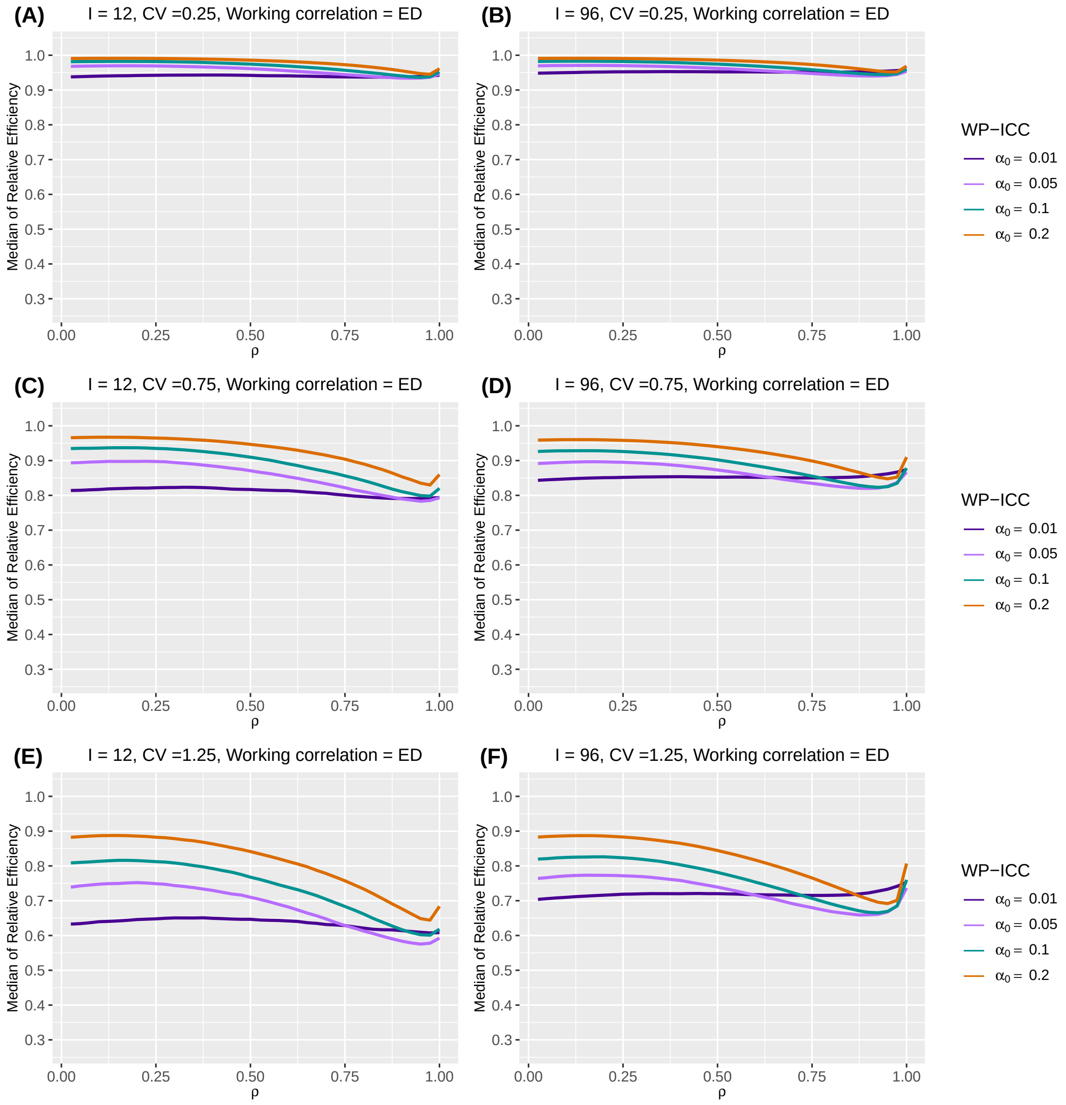

**Web Figure 31** The median of relative efficiency (RE) as a function of within-period intraclass correlation coefficient (WP-ICC) $\alpha_0 \in \{0.01, 0.05, 0.1, 0.2\}$ and the decay rate $\rho \in (0, 1)$, when both the true correlation model and the working correlation model are exponential decay (ED). Design factors considered are as follows: number of clusters $I = 12$ and 96, number of periods $J = 5$, mean cluster-period size $\overline{n} = 100$, and the degree of between-cluster imbalance is defined by coefficient of variation CV $\in \{0.25, 0.75, 1.25\}$. Within-cluster imbalance (pattern 4: randomly permuted) is introduced.

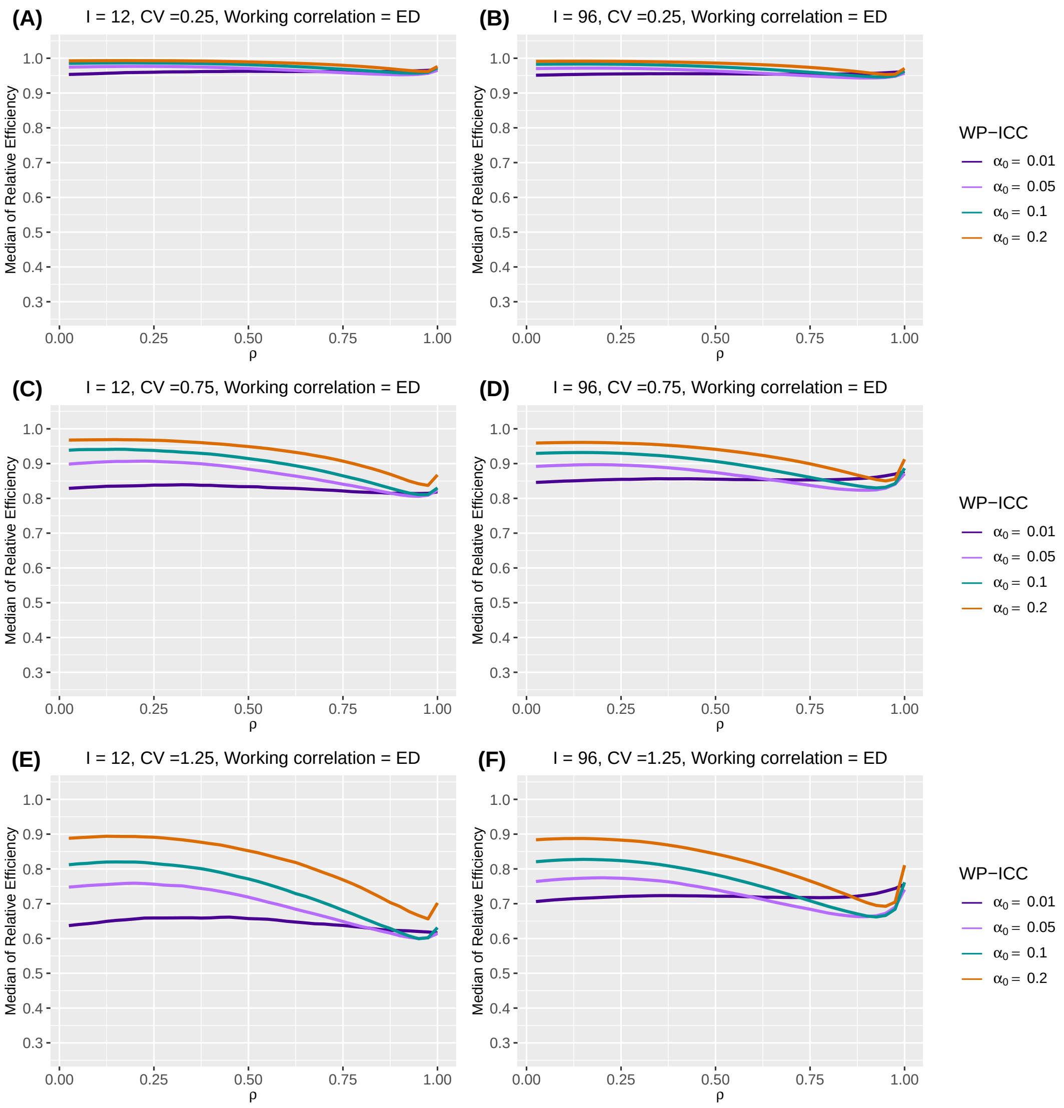

**Web Figure 32** The median of relative efficiency (RE) as a function of within-period intraclass correlation coefficient (WP-ICC) $\alpha_0 \in \{0.01, 0.05, 0.1, 0.2\}$ and the decay rate $\rho \in (0, 1)$, when the true correlation model is exponential decay (ED) and the working correlation model is independence (IND). Design factors considered are as follows: number of clusters $I = 12$ and $96$, number of periods $J = 5$, mean cluster-period size $\overline{n} = 100$, and the degree of between-cluster imbalance is defined by coefficient of variation CV $\in \{0.25, 0.75, 1.25\}$. No within-cluster imbalance is introduced.

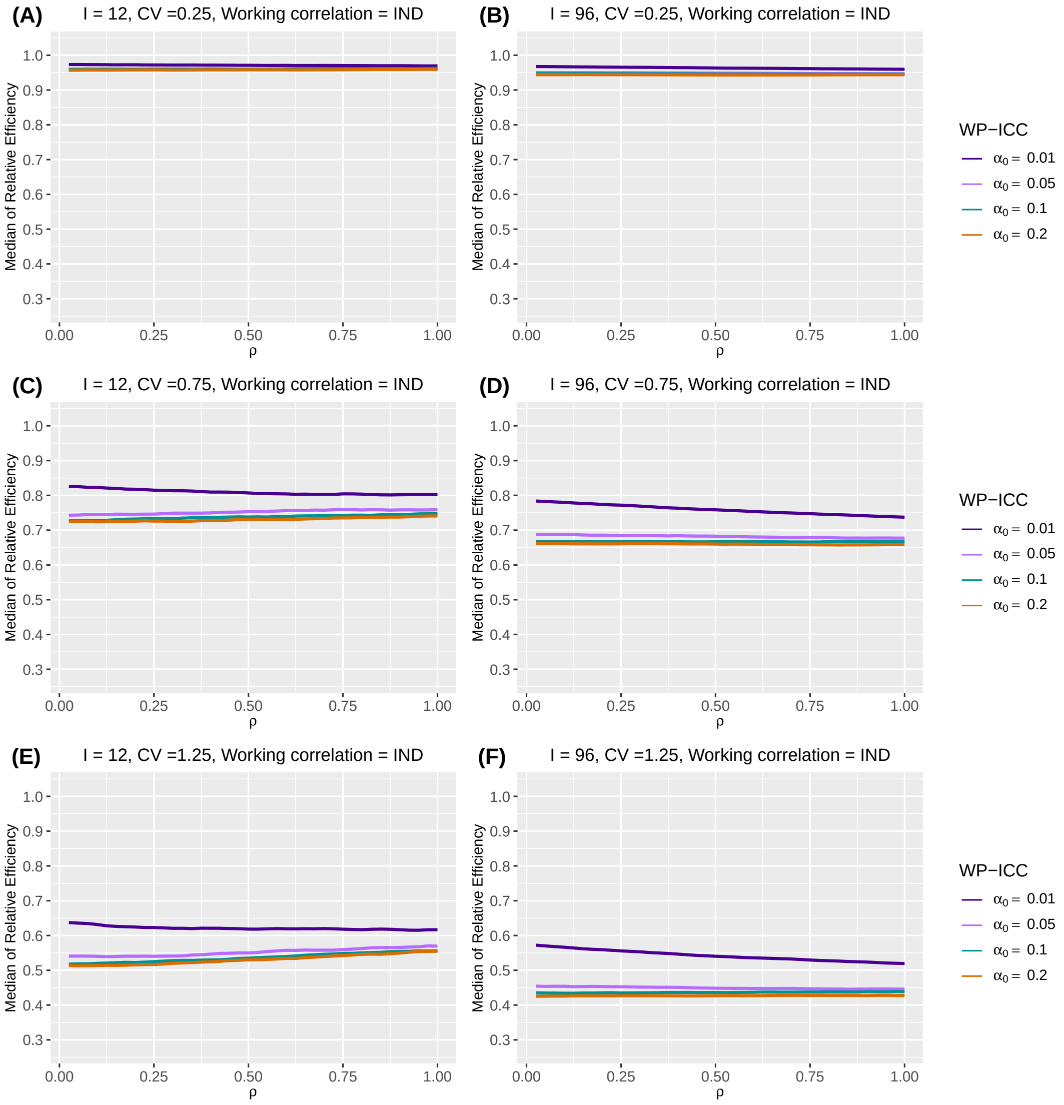

**Web Figure 33** The median of relative efficiency (RE) as a function of within-period intraclass correlation coefficient (WP-ICC) $\alpha_0 \in \{0.01, 0.05, 0.1, 0.2\}$ and the decay rate $\rho \in (0, 1)$, when the true correlation model is exponential decay (ED) and the working correlation model is independence (IND). Design factors considered are as follows: number of clusters $I = 12$ and $96$, number of periods $J = 5$, mean cluster-period size $\overline{n} = 100$, and the degree of between-cluster imbalance is defined by coefficient of variation CV $\in \{0.25, 0.75, 1.25\}$. Within-cluster imbalance (pattern 1: constant) is introduced.

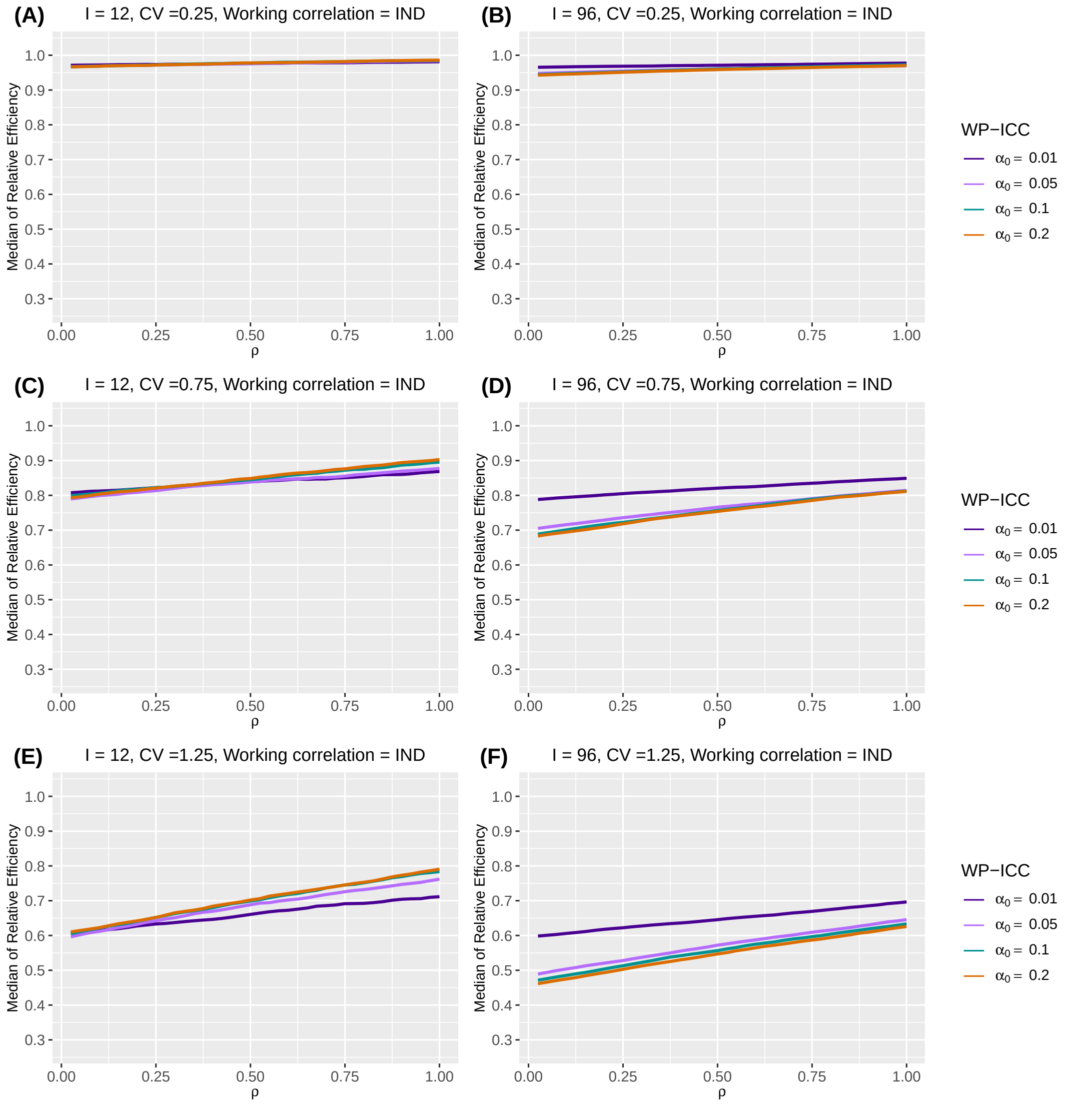

**Web Figure 34** The median of relative efficiency (RE) as a function of within-period intraclass correlation coefficient (WP-ICC) $\alpha_0 \in \{0.01, 0.05, 0.1, 0.2\}$ and the decay rate $\rho \in (0, 1)$, when the true correlation model is exponential decay (ED) and the working correlation model is independence (IND). Design factors considered are as follows: number of clusters $I = 12$ and $96$, number of periods $J = 5$, mean cluster-period size $\overline{n} = 100$, and the degree of between-cluster imbalance is defined by coefficient of variation CV $\in \{0.25, 0.75, 1.25\}$. Within-cluster imbalance (pattern 2: monotonically increasing) is introduced.

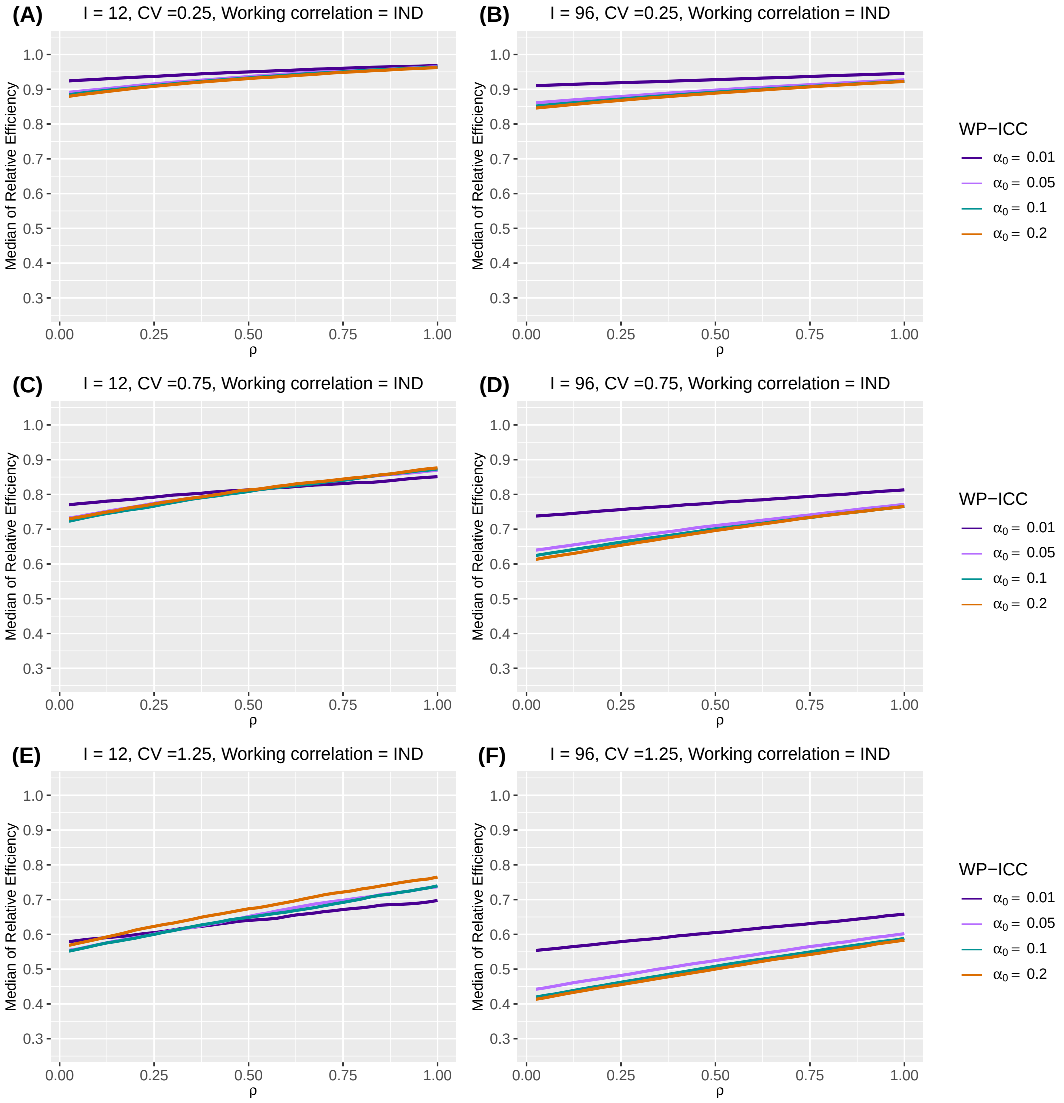

**Web Figure 35** The median of relative efficiency (RE) as a function of within-period intraclass correlation coefficient (WP-ICC) $\alpha_0 \in \{0.01, 0.05, 0.1, 0.2\}$ and the decay rate $\rho \in (0, 1)$, when the true correlation model is exponential decay (ED) and the working correlation model is independence (IND). Design factors considered are as follows: number of clusters $I = 12$ and $96$, number of periods $J = 5$, mean cluster-period size $\overline{n} = 100$, and the degree of between-cluster imbalance is defined by coefficient of variation CV $\in \{0.25, 0.75, 1.25\}$. Within-cluster imbalance (pattern 3: monotonically decreasing) is introduced.

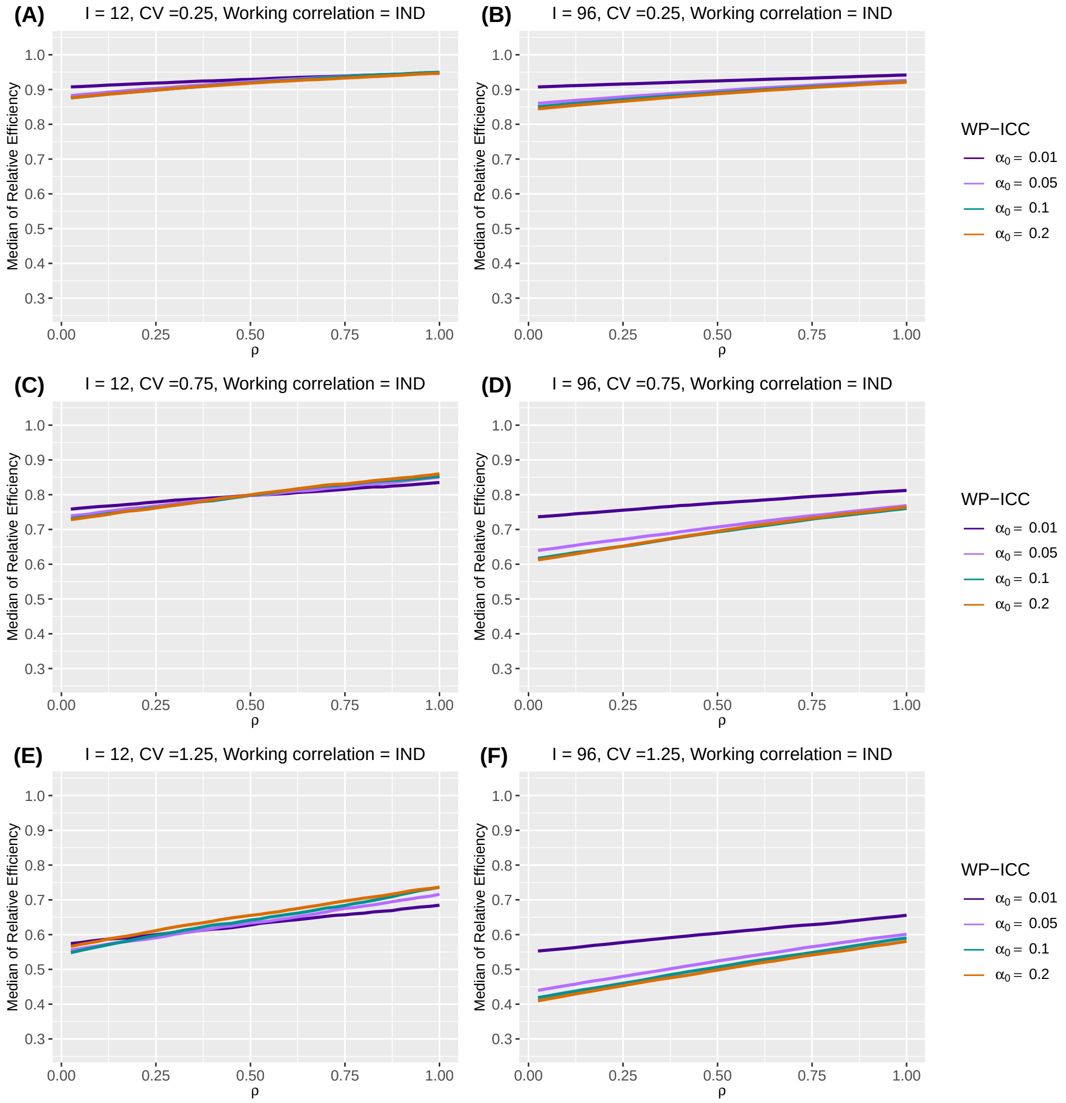

**Web Figure 36** The median of relative efficiency (RE) as a function of within-period intraclass correlation coefficient (WP-ICC) $\alpha_0 \in \{0.01, 0.05, 0.1, 0.2\}$ and the decay rate $\rho \in (0,1)$, when the true correlation model is exponential decay (ED) and the working correlation model is independence (IND). Design factors considered are as follows: number of clusters $I = 12$ and $96$, number of periods $J = 5$, mean cluster-period size $\overline{n} = 100$, and the degree of between-cluster imbalance is defined by coefficient of variation CV $\in \{0.25, 0.75, 1.25\}$. Within-cluster imbalance (pattern 4: randomly permuted) is introduced.

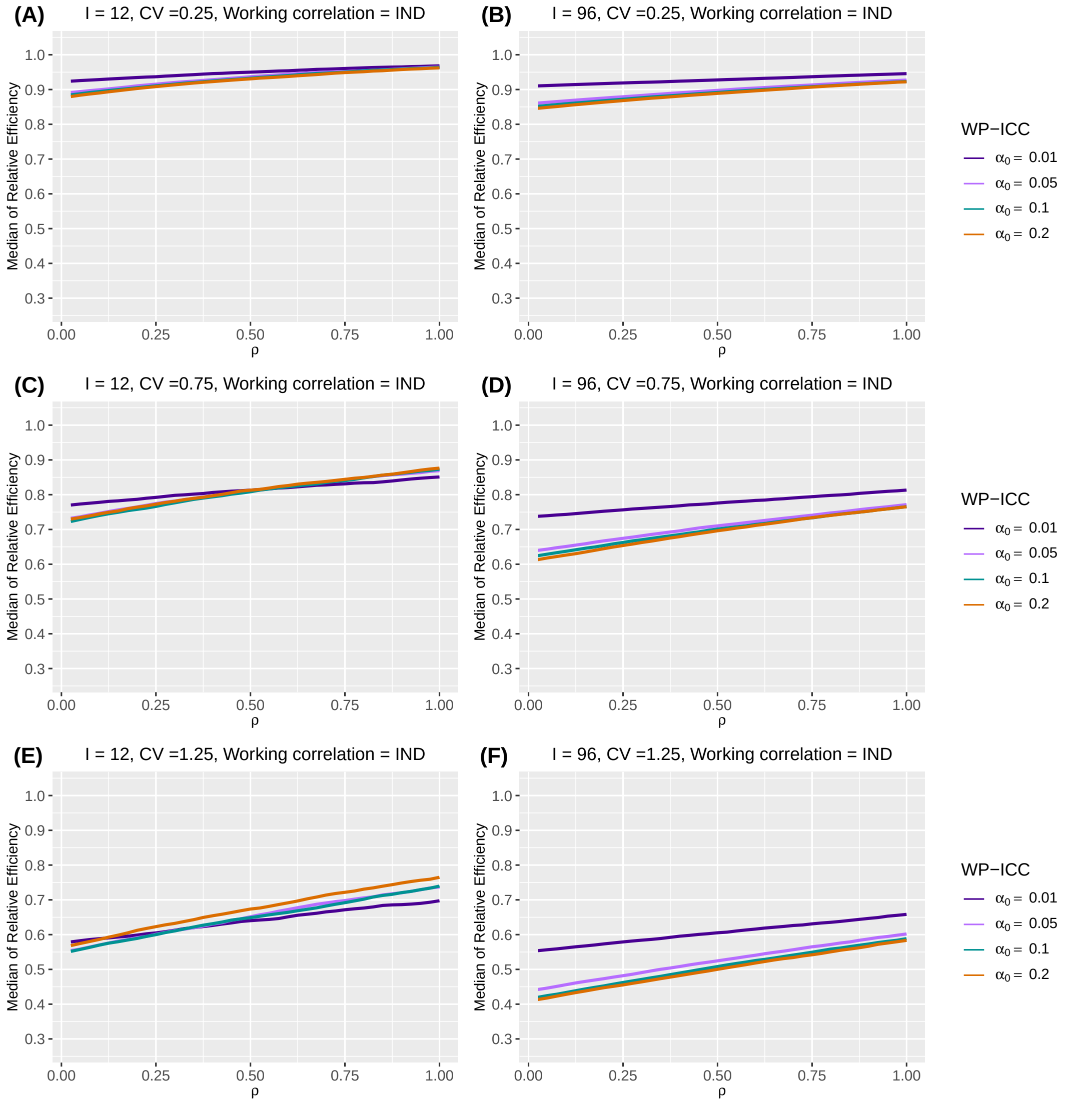

##### Web Appendix D.3. Number of periods

Tables in this section show the counterparts to tables that illustrate the impact of number of clusters on RE, but under exponential decay true correlation structure.

**Web Table 12** Median and interquartile range (IQR) (in parentheses) of relative efficiency (RE) as a function of periods $J$, under different degrees of between- and within-cluster imbalance, when the true correlation model is exponential decay (ED). Number of clusters is $I = 12$. The within-period intraclass correlation coefficient (WP-ICC) $\alpha_0$ is $0.05$, and the decay rate $\rho$ is $0.5$. Between cluster imbalance is measured by coefficient of variation, CV $\in \{0.25, 0.75, 1.25\}$. The working correlation structure is either ED or independence (IND).

| Working correlation | $J$ | CV | No within-cluster imbalance | Within-cluster imbalance pattern 1 | Within-cluster imbalance pattern 2 | Within-cluster imbalance pattern 4 |
|---|---|---|---|---|---|---|
| NEX | 3 | 0.25 | 0.989 (0.984, 0.992) | 0.986 (0.969, 1.001) | 0.964 (0.947, 0.977) | 0.964 (0.948, 0.979) |
| | | 0.75 | 0.902 (0.872, 0.928) | 0.884 (0.803, 0.944) | 0.856 (0.776, 0.915) | 0.863 (0.782, 0.920) |
| | | 1.25 | 0.756 (0.704, 0.804) | 0.704 (0.590, 0.817) | 0.679 (0.571, 0.791) | 0.684 (0.567, 0.800) |
| | 5 | 0.25 | 0.990 (0.987, 0.992) | 0.988 (0.978, 0.997) | 0.970 (0.958, 0.98) | 0.970 (0.959, 0.980) |
| | | 0.75 | 0.913 (0.888, 0.936) | 0.897 (0.842, 0.940) | 0.884 (0.825, 0.925) | 0.881 (0.825, 0.925) |
| | | 1.25 | 0.773 (0.726, 0.819) | 0.729 (0.648, 0.809) | 0.720 (0.637, 0.803) | 0.719 (0.635, 0.796) |
| | 13 | 0.25 | 0.993 (0.990, 0.995) | 0.991 (0.988, 0.994) | 0.978 (0.974, 0.981) | 0.978 (0.975, 0.981) |
| | | 0.75 | 0.933 (0.910, 0.952) | 0.930 (0.903, 0.950) | 0.917 (0.890, 0.936) | 0.916 (0.890, 0.936) |
| | | 1.25 | 0.810 (0.769, 0.852) | 0.811 (0.761, 0.852) | 0.795 (0.744, 0.841) | 0.795 (0.752, 0.84) |
| IND | 3 | 0.25 | 0.961 (0.948, 0.972) | 0.970 (0.951, 0.988) | 0.891 (0.871, 0.908) | 0.890 (0.87, 0.907) |
| | | 0.75 | 0.755 (0.694, 0.806) | 0.825 (0.749, 0.899) | 0.749 (0.674, 0.818) | 0.755 (0.676, 0.823) |
| | | 1.25 | 0.547 (0.474, 0.621) | 0.666 (0.583, 0.769) | 0.611 (0.529, 0.704) | 0.605 (0.531, 0.707) |
| | 5 | 0.25 | 0.962 (0.939, 0.980) | 0.977 (0.954, 0.998) | 0.933 (0.907, 0.956) | 0.936 (0.910, 0.959) |
| | | 0.75 | 0.752 (0.687, 0.819) | 0.841 (0.766, 0.911) | 0.813 (0.738, 0.882) | 0.805 (0.725, 0.883) |
| | | 1.25 | 0.546 (0.466, 0.628) | 0.678 (0.570, 0.771) | 0.651 (0.546, 0.748) | 0.645 (0.551, 0.747) |
| | 13 | 0.25 | 0.961 (0.943, 0.974) | 0.982 (0.976, 0.989) | 0.950 (0.937, 0.961) | 0.950 (0.937, 0.962) |
| | | 0.75 | 0.746 (0.677, 0.805) | 0.873 (0.823, 0.909) | 0.848 (0.786, 0.886) | 0.847 (0.796, 0.889) |
| | | 1.25 | 0.530 (0.452, 0.610) | 0.699 (0.621, 0.772) | 0.681 (0.603, 0.749) | 0.679 (0.597, 0.754) |

**Web Table 13** Median and interquartile range (IQR) (in parentheses) of relative efficiency (RE) as a function of periods $J$, under different degrees of between- and within-cluster imbalance, when the true correlation model is exponential decay (ED). Number of clusters is $I = 24$. The within-period intraclass correlation coefficient (WP-ICC) $\alpha_0$ is $0.05$, and the decay rate $\rho$ is $0.5$. Between cluster imbalance is measured by coefficient of variation, CV $\in \{0.25, 0.75, 1.25\}$. The working correlation structure is either ED or independence (IND).

| Working correlation | $J$ | CV | No within-cluster imbalance | Within-cluster imbalance pattern 1 | Within-cluster imbalance pattern 2 | Within-cluster imbalance pattern 4 |
|---|---|---|---|---|---|---|
| NEX | 3 | 0.25 | 0.987 (0.984, 0.990) | 0.984 (0.972, 0.994) | 0.962 (0.950, 0.973) | 0.962 (0.951, 0.974) |
| | | 0.75 | 0.898 (0.876, 0.916) | 0.877 (0.822, 0.921) | 0.854 (0.806, 0.894) | 0.855 (0.806, 0.897) |
| | | 1.25 | 0.758 (0.720, 0.793) | 0.707 (0.630, 0.776) | 0.696 (0.622, 0.769) | 0.684 (0.605, 0.761) |
| | 5 | 0.25 | 0.989 (0.987, 0.991) | 0.987 (0.977, 0.996) | 0.960 (0.950, 0.969) | 0.960 (0.951, 0.969) |
| | | 0.75 | 0.909 (0.891, 0.926) | 0.897 (0.855, 0.932) | 0.873 (0.830, 0.908) | 0.871 (0.825, 0.916) |
| | | 1.25 | 0.774 (0.743, 0.807) | 0.738 (0.671, 0.801) | 0.719 (0.654, 0.786) | 0.725 (0.658, 0.794) |
| | 13 | 0.25 | 0.992 (0.990, 0.994) | 0.991 (0.987, 0.993) | 0.979 (0.975, 0.982) | 0.979 (0.976, 0.982) |
| | | 0.75 | 0.929 (0.912, 0.942) | 0.921 (0.902, 0.938) | 0.910 (0.890, 0.926) | 0.912 (0.892, 0.930) |
| | | 1.25 | 0.810 (0.780, 0.838) | 0.796 (0.760, 0.830) | 0.788 (0.748, 0.821) | 0.787 (0.750, 0.820) |
| IND | 3 | 0.25 | 0.955 (0.945, 0.964) | 0.958 (0.942, 0.972) | 0.878 (0.863, 0.892) | 0.881 (0.864, 0.894) |
| | | 0.75 | 0.721 (0.673, 0.763) | 0.767 (0.696, 0.831) | 0.693 (0.629, 0.756) | 0.700 (0.642, 0.756) |
| | | 1.25 | 0.501 (0.440, 0.560) | 0.591 (0.508, 0.659) | 0.532 (0.464, 0.611) | 0.525 (0.452, 0.605) |
| | 5 | 0.25 | 0.954 (0.938, 0.970) | 0.970 (0.946, 0.993) | 0.896 (0.875, 0.914) | 0.895 (0.873, 0.914) |
| | | 0.75 | 0.720 (0.661, 0.771) | 0.812 (0.748, 0.878) | 0.753 (0.687, 0.809) | 0.748 (0.683, 0.811) |
| | | 1.25 | 0.499 (0.429, 0.561) | 0.633 (0.545, 0.719) | 0.584 (0.504, 0.663) | 0.582 (0.493, 0.670) |
| | 13 | 0.25 | 0.953 (0.939, 0.965) | 0.981 (0.973, 0.988) | 0.954 (0.944, 0.961) | 0.955 (0.944, 0.962) |
| | | 0.75 | 0.709 (0.652, 0.757) | 0.870 (0.831, 0.897) | 0.843 (0.802, 0.874) | 0.845 (0.803, 0.877) |
| | | 1.25 | 0.480 (0.419, 0.541) | 0.695 (0.622, 0.752) | 0.673 (0.598, 0.736) | 0.675 (0.612, 0.738) |

**Web Table 14** Median and interquartile range (IQR) (in parentheses) of relative efficiency (RE) as a function of periods $J$, under different degrees of between- and within-cluster imbalance, when the true correlation model is exponential decay (ED). Number of clusters is $I = 48$. The within-period intraclass correlation coefficient (WP-ICC) $\alpha_0$ is $0.05$, and the decay rate $\rho$ is $0.5$. Between cluster imbalance is measured by coefficient of variation, CV $\in \{0.25, 0.75, 1.25\}$. The working correlation structure is either ED or independence (IND).

| Working correlation | $J$ | CV | No within-cluster imbalance | Within-cluster imbalance pattern 1 | Within-cluster imbalance pattern 2 | Within-cluster imbalance pattern 4 |
|---|---|---|---|---|---|---|
| NEX | 3 | 0.25 | 0.987 (0.985, 0.989) | 0.984 (0.976, 0.992) | 0.962 (0.954, 0.971) | 0.961 (0.953, 0.969) |
| | | 0.75 | 0.895 (0.880, 0.908) | 0.872 (0.837, 0.903) | 0.855 (0.822, 0.885) | 0.853 (0.821, 0.882) |
| | | 1.25 | 0.756 (0.727, 0.781) | 0.715 (0.658, 0.761) | 0.695 (0.644, 0.746) | 0.698 (0.644, 0.742) |
| | 5 | 0.25 | 0.989 (0.987, 0.990) | 0.987 (0.980, 0.993) | 0.962 (0.955, 0.969) | 0.961 (0.954, 0.968) |
| | | 0.75 | 0.906 (0.893, 0.917) | 0.896 (0.866, 0.922) | 0.875 (0.845, 0.898) | 0.874 (0.845, 0.899) |
| | | 1.25 | 0.774 (0.750, 0.795) | 0.747 (0.699, 0.796) | 0.731 (0.687, 0.778) | 0.732 (0.683, 0.780) |
| | 13 | 0.25 | 0.992 (0.991, 0.993) | 0.991 (0.987, 0.995) | 0.982 (0.978, 0.986) | 0.982 (0.978, 0.987) |
| | | 0.75 | 0.926 (0.915, 0.936) | 0.922 (0.902, 0.939) | 0.914 (0.896, 0.933) | 0.914 (0.896, 0.932) |
| | | 1.25 | 0.809 (0.788, 0.827) | 0.807 (0.773, 0.839) | 0.801 (0.766, 0.833) | 0.802 (0.769, 0.831) |
| IND | 3 | 0.25 | 0.952 (0.945, 0.959) | 0.951 (0.939, 0.962) | 0.874 (0.860, 0.884) | 0.871 (0.859, 0.882) |
| | | 0.75 | 0.701 (0.665, 0.734) | 0.725 (0.669, 0.778) | 0.666 (0.619, 0.707) | 0.661 (0.612, 0.711) |
| | | 1.25 | 0.469 (0.419, 0.514) | 0.530 (0.461, 0.590) | 0.481 (0.421, 0.538) | 0.476 (0.417, 0.536) |
| | 5 | 0.25 | 0.951 (0.938, 0.962) | 0.965 (0.947, 0.980) | 0.908 (0.894, 0.924) | 0.908 (0.893, 0.922) |
| | | 0.75 | 0.694 (0.655, 0.737) | 0.787 (0.734, 0.832) | 0.737 (0.686, 0.781) | 0.736 (0.690, 0.780) |
| | | 1.25 | 0.462 (0.407, 0.516) | 0.597 (0.533, 0.666) | 0.558 (0.494, 0.616) | 0.551 (0.487, 0.615) |
| | 13 | 0.25 | 0.949 (0.939, 0.959) | 0.978 (0.967, 0.988) | 0.961 (0.950, 0.971) | 0.962 (0.951, 0.973) |
| | | 0.75 | 0.689 (0.646, 0.722) | 0.853 (0.814, 0.885) | 0.833 (0.794, 0.872) | 0.836 (0.796, 0.870) |
| | | 1.25 | 0.452 (0.398, 0.499) | 0.682 (0.619, 0.740) | 0.672 (0.610, 0.726) | 0.671 (0.609, 0.722) |

**Web Table 15** Median and interquartile range (IQR) (in parentheses) of relative efficiency (RE) as a function of periods $J$, under different degrees of between- and within-cluster imbalance, when the true correlation model is exponential decay (ED). Number of clusters is $I = 96$. The within-period intraclass correlation coefficient (WP-ICC) $\alpha_0$ is $0.05$, and the decay rate $\rho$ is $0.5$. Between cluster imbalance is measured by coefficient of variation, CV $\in \{0.25, 0.75, 1.25\}$. The working correlation structure is either ED or independence (IND).

| Working correlation | $J$ | CV | No within-cluster imbalance | Within-cluster imbalance pattern 1 | Within-cluster imbalance pattern 2 | Within-cluster imbalance pattern 4 |
|---|---|---|---|---|---|---|
| NEX | 3 | 0.25 | 0.987 (0.985, 0.988) | 0.983 (0.978, 0.989) | 0.961 (0.956, 0.967) | 0.961 (0.955, 0.967) |
| | | 0.75 | 0.893 (0.882, 0.903) | 0.871 (0.847, 0.895) | 0.853 (0.829, 0.875) | 0.854 (0.828, 0.875) |
| | | 1.25 | 0.755 (0.736, 0.772) | 0.708 (0.671, 0.743) | 0.695 (0.661, 0.727) | 0.693 (0.652, 0.726) |
| | 5 | 0.25 | 0.989 (0.988, 0.990) | 0.986 (0.981, 0.992) | 0.963 (0.958, 0.969) | 0.963 (0.958, 0.968) |
| | | 0.75 | 0.905 (0.896, 0.913) | 0.892 (0.874, 0.914) | 0.876 (0.855, 0.896) | 0.874 (0.852, 0.893) |
| | | 1.25 | 0.773 (0.756, 0.789) | 0.753 (0.723, 0.785) | 0.739 (0.707, 0.768) | 0.740 (0.705, 0.771) |
| | 13 | 0.25 | 0.992 (0.991, 0.993) | 0.991 (0.986, 0.995) | 0.985 (0.981, 0.990) | 0.986 (0.981, 0.990) |
| | | 0.75 | 0.925 (0.917, 0.932) | 0.924 (0.905, 0.940) | 0.920 (0.901, 0.936) | 0.920 (0.902, 0.937) |
| | | 1.25 | 0.807 (0.792, 0.823) | 0.813 (0.786, 0.845) | 0.812 (0.782, 0.838) | 0.810 (0.781, 0.841) |
| IND | 3 | 0.25 | 0.951 (0.946, 0.956) | 0.947 (0.938, 0.956) | 0.870 (0.861, 0.879) | 0.870 (0.861, 0.878) |
| | | 0.75 | 0.689 (0.662, 0.714) | 0.706 (0.660, 0.743) | 0.643 (0.604, 0.679) | 0.641 (0.603, 0.680) |
| | | 1.25 | 0.450 (0.414, 0.488) | 0.482 (0.430, 0.534) | 0.441 (0.396, 0.486) | 0.438 (0.390, 0.485) |
| | 5 | 0.25 | 0.949 (0.941, 0.957) | 0.962 (0.950, 0.973) | 0.898 (0.887, 0.910) | 0.898 (0.886, 0.909) |
| | | 0.75 | 0.680 (0.649, 0.714) | 0.767 (0.730, 0.804) | 0.713 (0.676, 0.747) | 0.710 (0.673, 0.743) |
| | | 1.25 | 0.445 (0.403, 0.484) | 0.566 (0.515, 0.617) | 0.516 (0.475, 0.564) | 0.522 (0.470, 0.572) |
| | 13 | 0.25 | 0.947 (0.940, 0.954) | 0.975 (0.964, 0.986) | 0.962 (0.952, 0.972) | 0.962 (0.951, 0.973) |
| | | 0.75 | 0.673 (0.641, 0.700) | 0.835 (0.802, 0.867) | 0.823 (0.789, 0.852) | 0.819 (0.786, 0.853) |
| | | 1.25 | 0.433 (0.394, 0.469) | 0.668 (0.618, 0.715) | 0.650 (0.599, 0.697) | 0.650 (0.602, 0.698) |

##### Web Appendix D.4. Cluster-period size

Figures in this section show the counterparts to plots that illustrate the impact of mean of cluster-period size on RE, but under exponential decay true correlation structure.

**Web Figure 37** The median of relative efficiency (RE) as a function of the within-period intraclass correlation coefficient (WP-ICC) $\alpha_0 \in \{0.01, 0.05, 0.1, 0.2\}$ and the decay rate $\rho \in (0, 1)$, when both the true correlation model and the working correlation model are exponential decay (ED). Design factors considered are as follows: number of clusters $I = 12$, number of periods $J = 5$, mean cluster-period sizes $\overline{n} \in \{50, 300\}$, and the degree of between-cluster imbalance is defined by coefficient of variation CV $\in \{0.25, 0.75, 1.25\}$. No within-cluster imbalance is introduced.

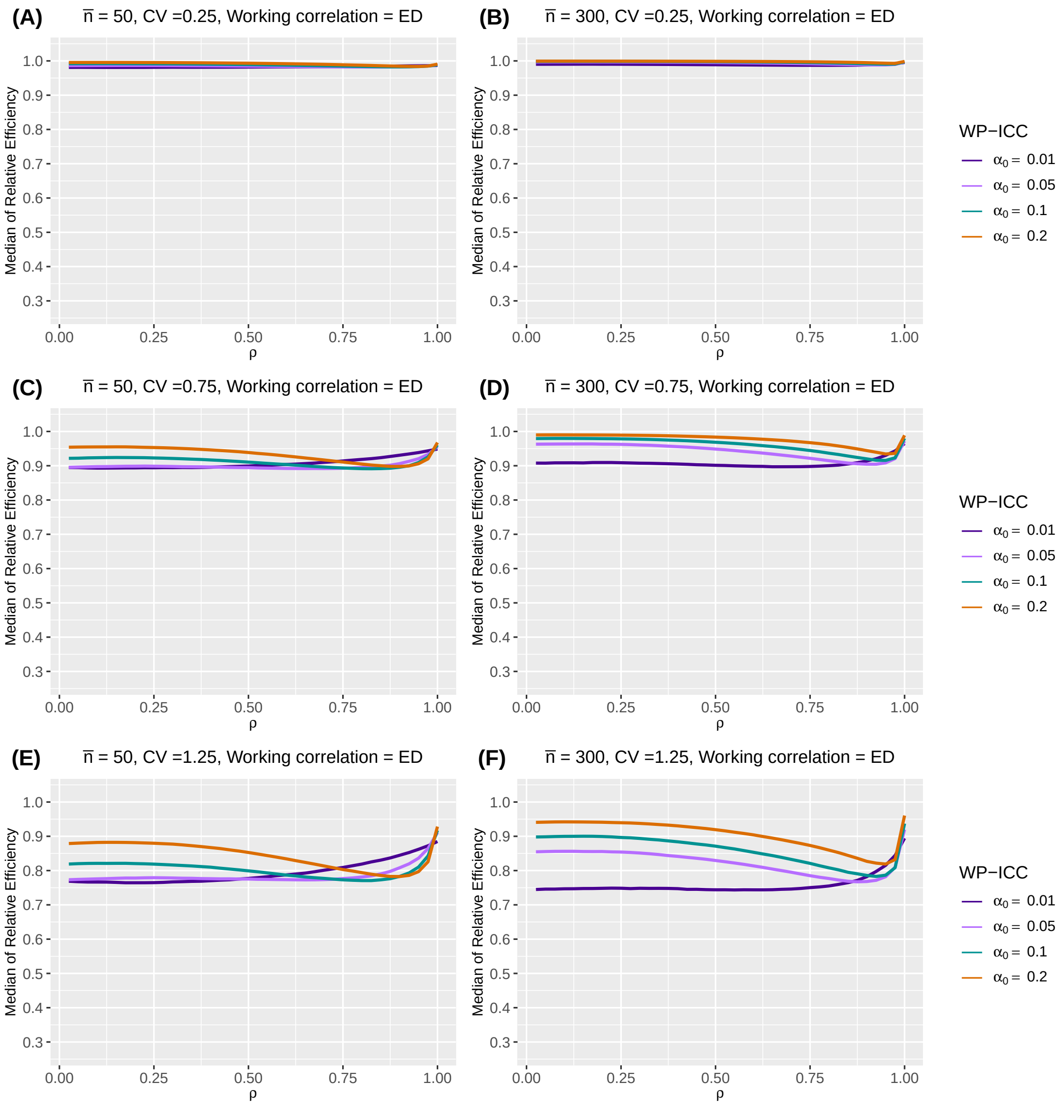

**Web Figure 38** The median of relative efficiency (RE) as a function of the within-period intraclass correlation coefficient (WP-ICC) $\alpha_0 \in \{0.01, 0.05, 0.1, 0.2\}$ and the decay rate $\rho \in (0, 1)$, when both the true correlation model and the working correlation model are exponential decay (ED). Design factors considered are as follows: number of clusters $I = 96$, number of periods $J = 5$, mean cluster-period sizes $\overline{n} \in \{50, 300\}$, and the degree of between-cluster imbalance is defined by coefficient of variation $\text{CV} \in \{0.25, 0.75, 1.25\}$. No within-cluster imbalance is introduced.

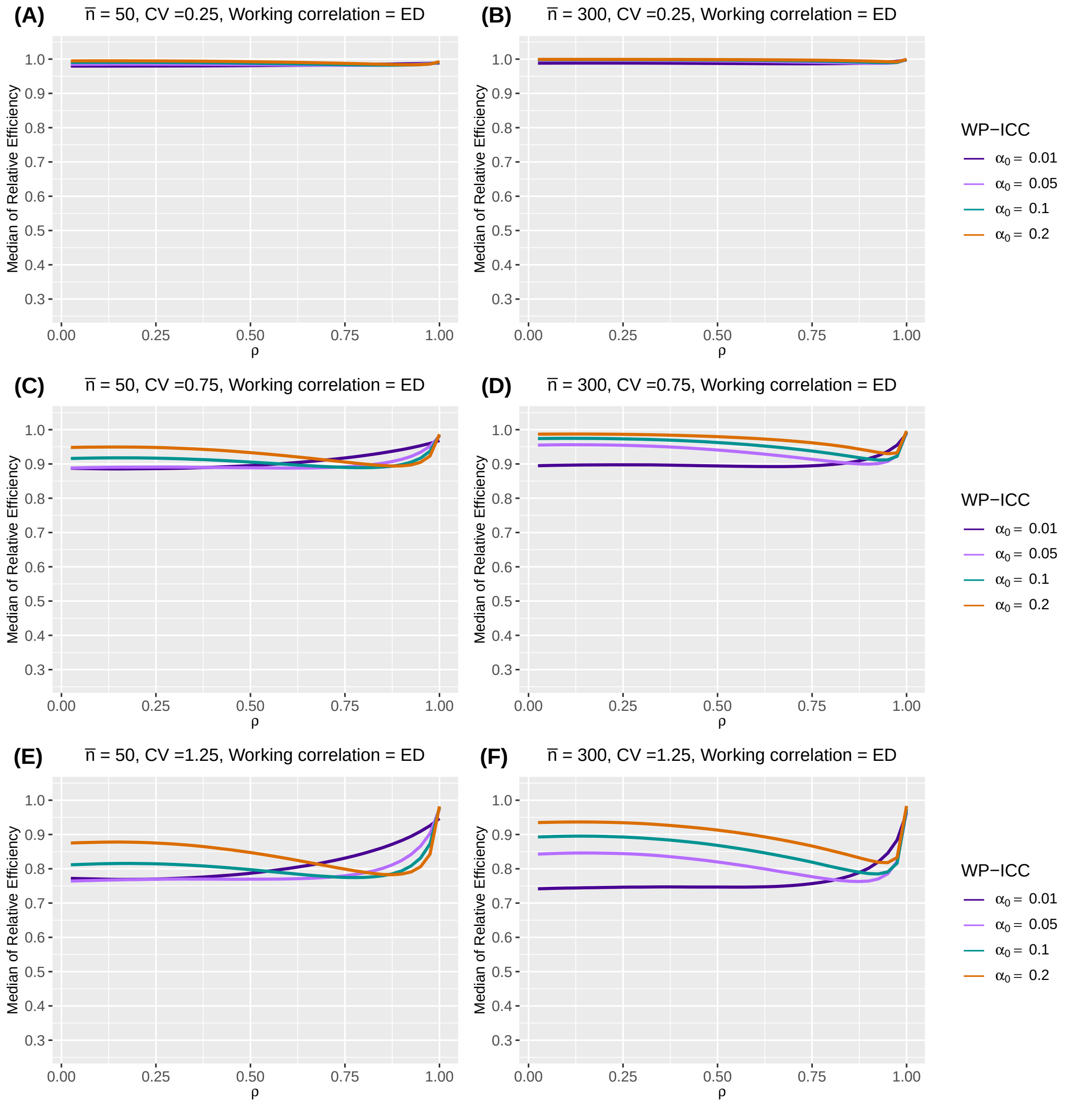

**Web Figure 39** The median of relative efficiency (RE) as a function of the within-period intraclass correlation coefficient (WP-ICC) $\alpha_0 \in \{0.01, 0.05, 0.1, 0.2\}$ and the decay rate $\rho \in (0, 1)$, when both the true correlation model and the working correlation model are exponential decay (ED). Design factors considered are as follows: number of clusters $I = 12$, number of periods $J = 5$, mean cluster-period sizes $\overline{n} \in \{50, 300\}$, and the degree of between-cluster imbalance is defined by coefficient of variation CV $\in \{0.25, 0.75, 1.25\}$. Within-cluster imbalance (pattern 4: randomly permuted) is introduced.

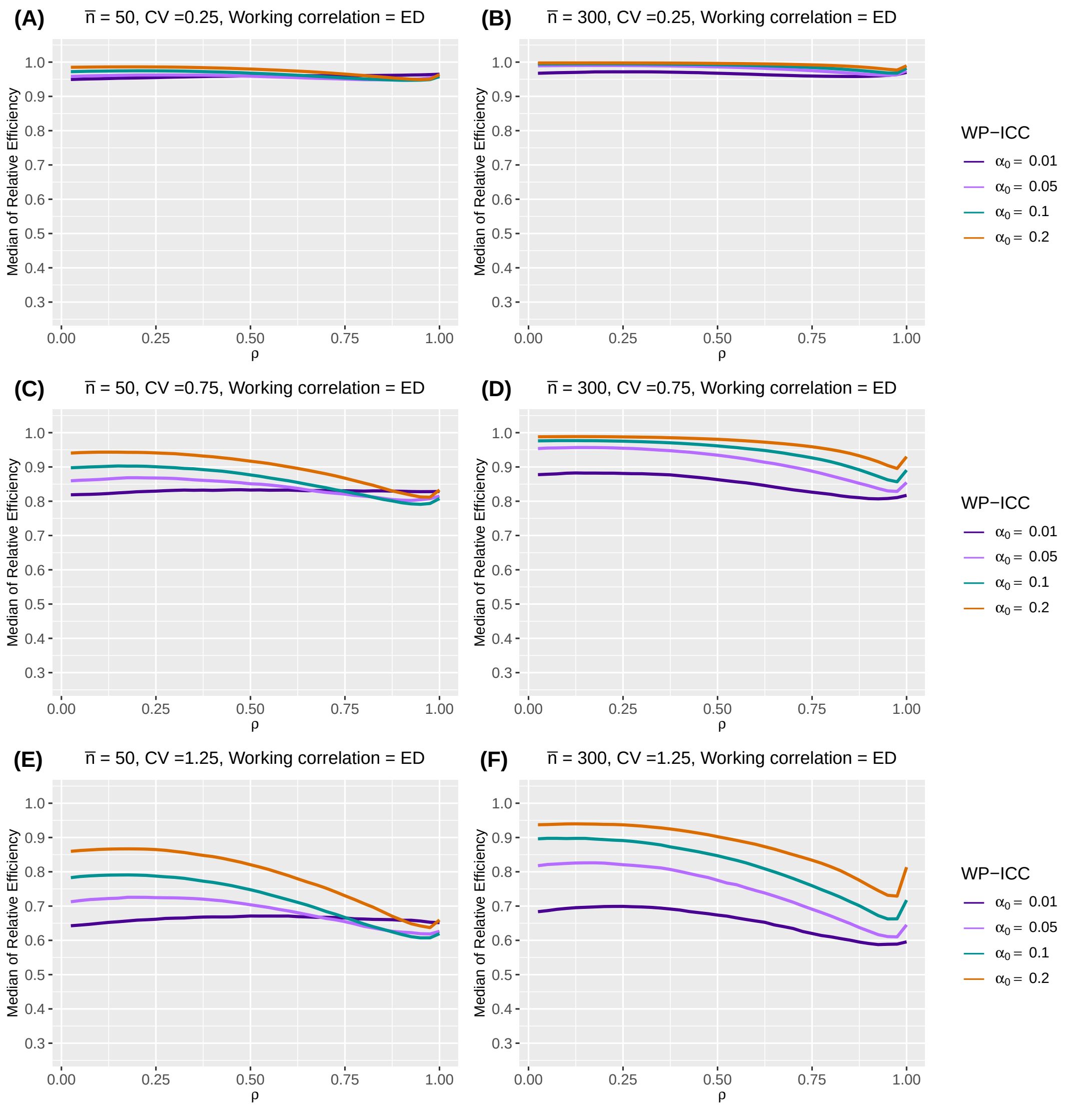

**Web Figure 40** The median of relative efficiency (RE) as a function of the within-period intraclass correlation coefficient (WP-ICC) $\alpha_0 \in \{0.01, 0.05, 0.1, 0.2\}$ and the decay rate $\rho \in (0,1)$, when both the true correlation model and the working correlation model are exponential decay (ED). Design factors considered are as follows: number of clusters $I = 96$, number of periods $J = 5$, mean cluster-period sizes $\overline{n} \in \{50, 300\}$, and the degree of between-cluster imbalance is defined by coefficient of variation CV $\in \{0.25, 0.75, 1.25\}$. Within-cluster imbalance (pattern 4: randomly permuted) is introduced.

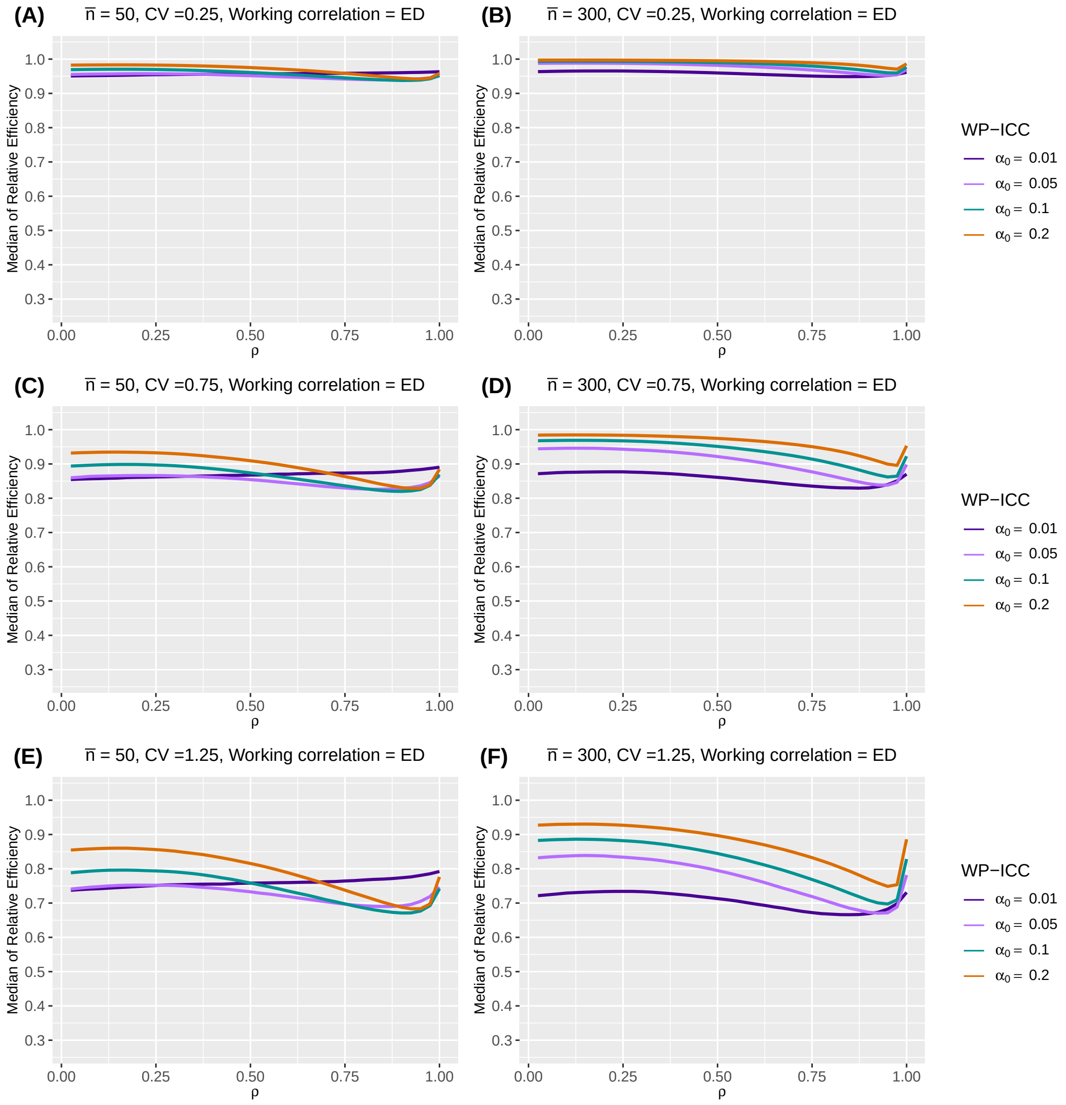

**Web Figure 41** The median of relative efficiency (RE) as a function of the within-period intraclass correlation coefficient (WP-ICC) $\alpha_0 \in \{0.01, 0.05, 0.1, 0.2\}$ and the decay rate $\rho \in (0,1)$, when the true correlation model is exponential decay (ED) and the working correlation model is independence (IND). Design factors considered are as follows: number of clusters $I = 12$, number of periods $J = 5$, mean cluster-period sizes $\overline{n} \in \{50, 300\}$, and the degree of between-cluster imbalance is defined by coefficient of variation CV $\in \{0.25, 0.75, 1.25\}$. No within-cluster imbalance is introduced.

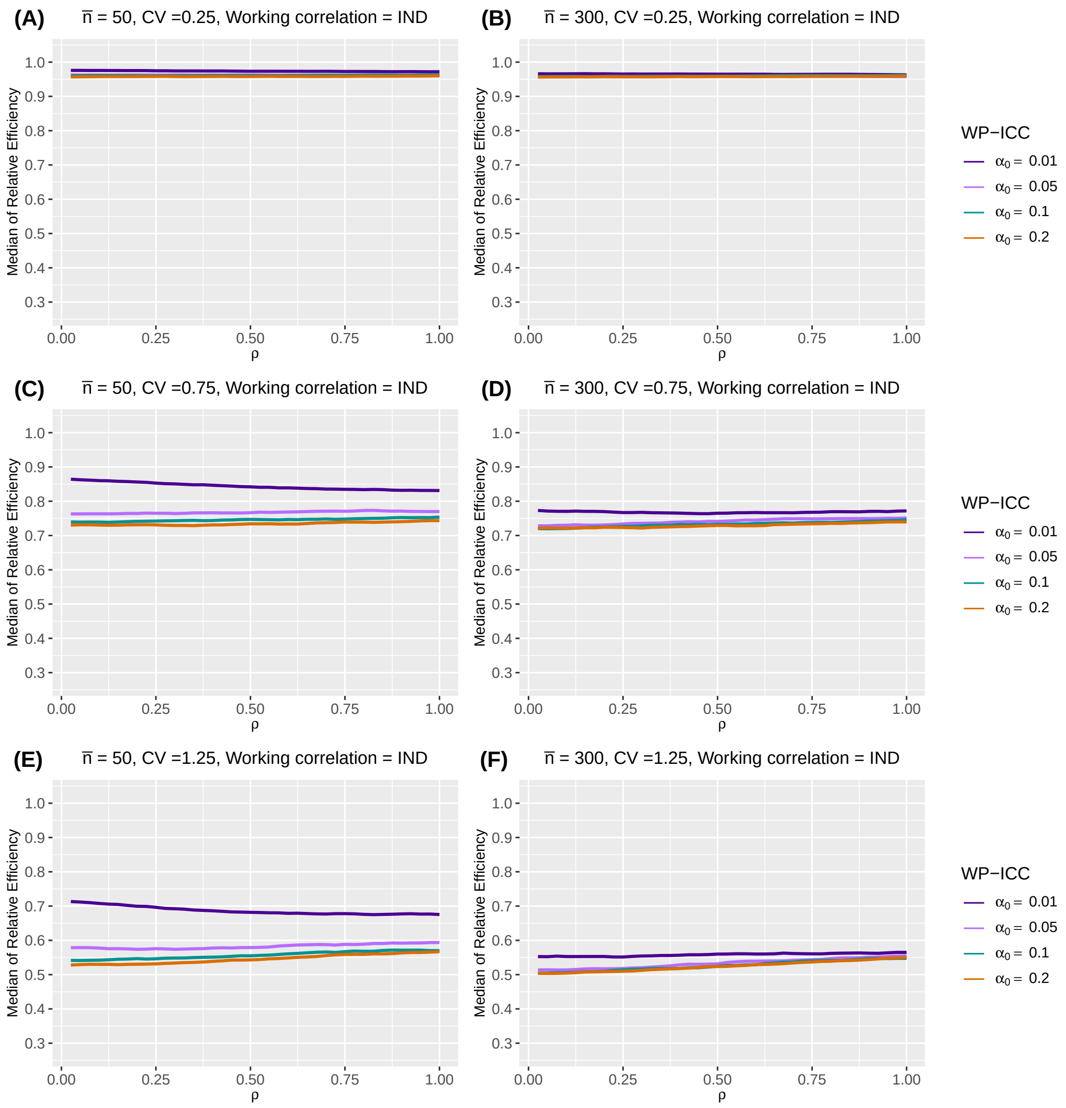

**Web Figure 42** The median of relative efficiency (RE) as a function of the within-period intraclass correlation coefficient (WP-ICC) $\alpha_0 \in \{0.01, 0.05, 0.1, 0.2\}$ and the decay rate $\rho \in (0, 1)$, when the true correlation model is exponential decay (ED) and the working correlation model is independence (IND). Design factors considered are as follows: number of clusters $I = 96$, number of periods $J = 5$, mean cluster-period sizes $\overline{n} \in \{50, 300\}$, and the degree of between-cluster imbalance is defined by coefficient of variation CV $\in \{0.25, 0.75, 1.25\}$. No within-cluster imbalance is introduced.

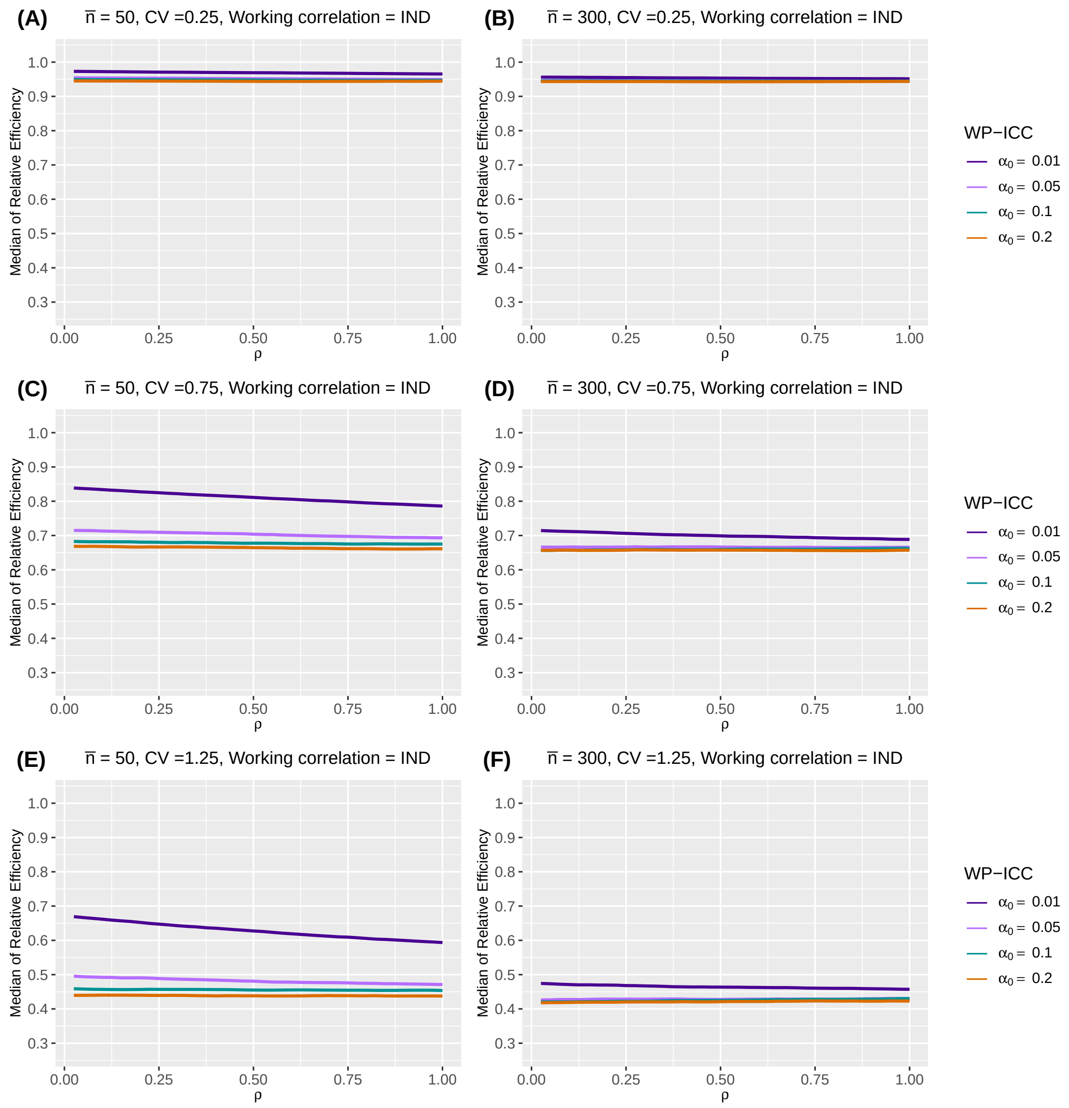

**Web Figure 43** The median of relative efficiency (RE) as a function of the within-period intraclass correlation coefficient (WP-ICC) $\alpha_0 \in \{0.01, 0.05, 0.1, 0.2\}$ and the decay rate $\rho \in (0, 1)$, when the true correlation model is exponential decay (ED) and the working correlation model is independence (IND). Design factors considered are as follows: number of clusters $I = 12$, number of periods $J = 5$, mean cluster-period sizes $\overline{n} \in \{50, 300\}$, and the degree of between-cluster imbalance is defined by coefficient of variation CV $\in \{0.25, 0.75, 1.25\}$. Within-cluster imbalance (pattern 4: randomly permuted) is introduced.

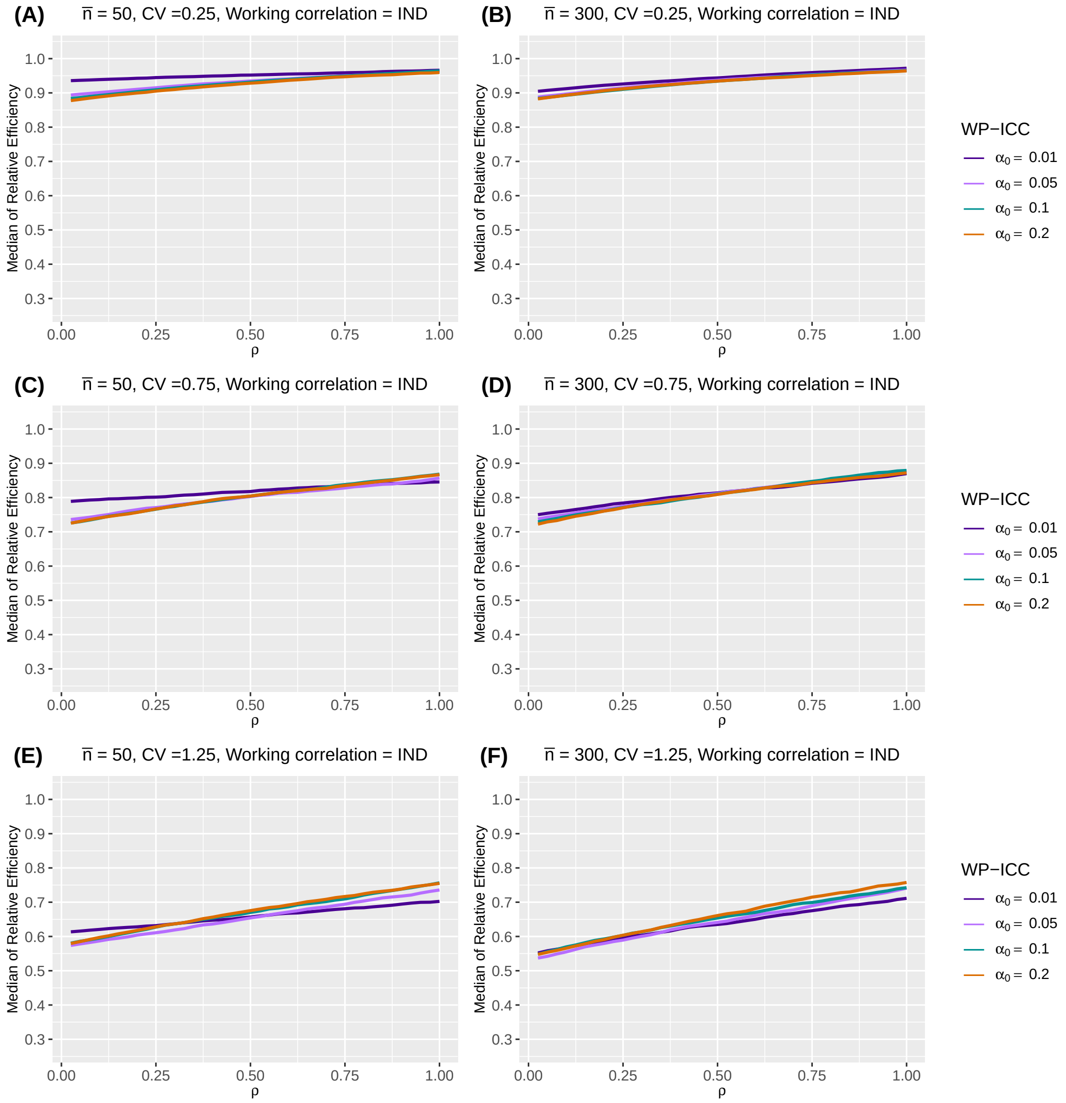

**Web Figure 44** The median of relative efficiency (RE) as a function of the within-period intraclass correlation coefficient (WP-ICC) $\alpha_0 \in \{0.01, 0.05, 0.1, 0.2\}$ and the decay rate $\rho \in (0, 1)$, when the true correlation model is exponential decay (ED) and the working correlation model is independence (IND). Design factors considered are as follows: number of clusters $I = 96$, number of periods $J = 5$, mean cluster-period sizes $\overline{n} \in \{50, 300\}$, and the degree of between-cluster imbalance is defined by coefficient of variation CV $\in \{0.25, 0.75, 1.25\}$. Within-cluster imbalance (pattern 4: randomly permuted) is introduced.

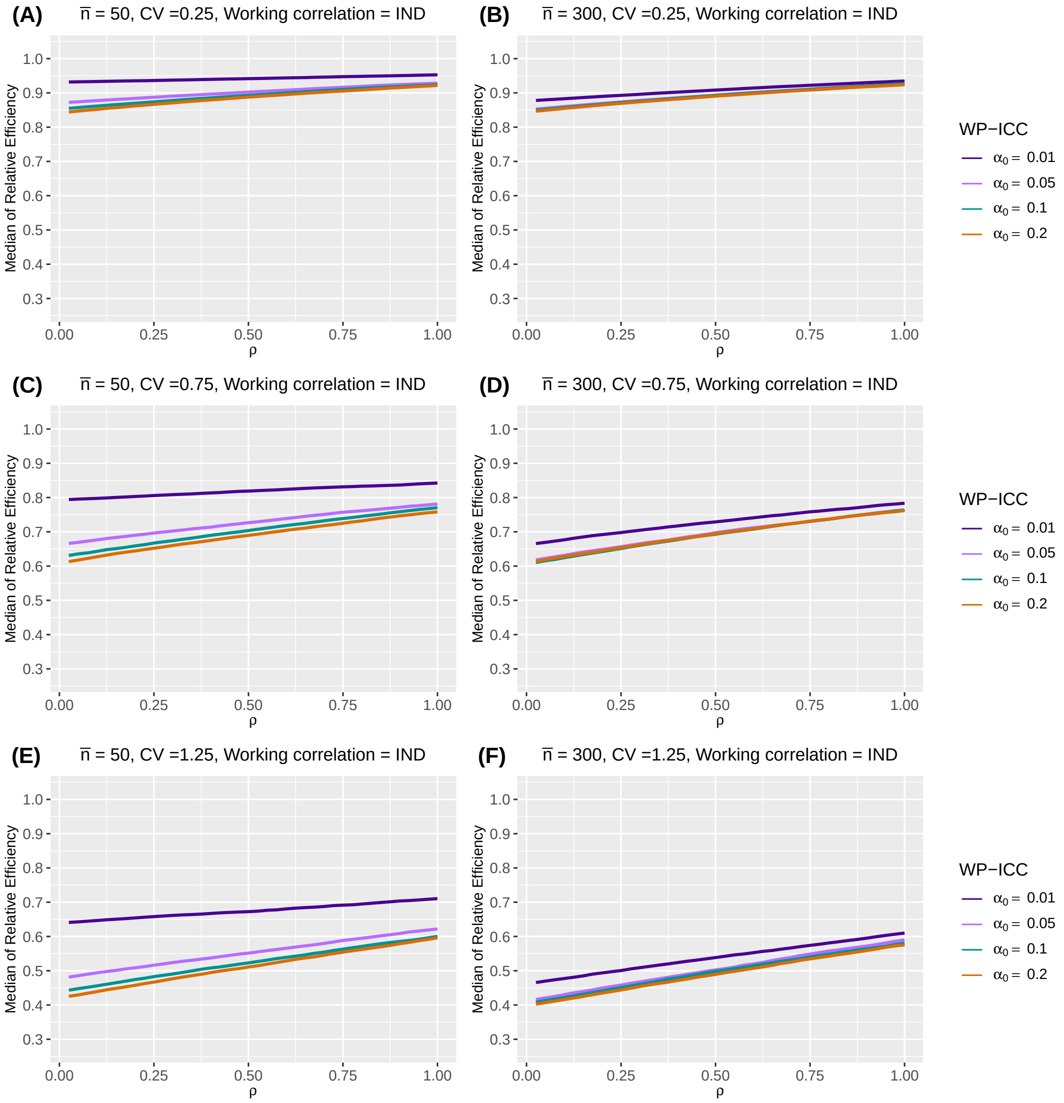

##### Web Appendix D.5. Sensitivity to baseline prevalence, intervention effect and secular trend

Tables in this section show the counterparts to tables that illustrate the impact of baseline prevalence, intervention effect and secular trend of the outcomes on RE, but under exponential decay true correlation structure.

**Web Table 16** Median and interquartile range (IQR) (in parentheses) of relative efficiency (RE) as a function of treatment effect $\delta$, baseline prevalence, and different secular trends, when the true correlation model is exponential decay (ED) and the working correlation correctly specifies the true correlation model. Design factors considered are as follows: number of clusters $I = 24$, number of periods $J = 5$, mean cluster-period size $\overline{n} = 100$. The within-period intraclass correlation coefficient (WP-ICC) $\alpha_0$ is $0.05$, and the decay rate $\rho$ is $0.9$. Between cluster imbalance is measured by coefficient of variation, CV $\in \{0.25, 0.75, 1.25\}$. No within-cluster imbalance is introduced.

| $\delta$ | CV | Baseline prevalence | Constant secular trend | Increasing secular trend | Decreasing secular trend |
|---|---|---|---|---|---|
| log(0.35) | 0.25 | 0.1 | 0.987 (0.984, 0.989) | 0.986 (0.984, 0.989) | 0.987 (0.984, 0.989) |
| | | 0.3 | 0.985 (0.983, 0.988) | 0.985 (0.982, 0.988) | 0.985 (0.983, 0.988) |
| | 0.75 | 0.1 | 0.908 (0.891, 0.924) | 0.905 (0.888, 0.922) | 0.907 (0.889, 0.921) |
| | | 0.3 | 0.902 (0.883, 0.918) | 0.899 (0.879, 0.916) | 0.901 (0.883, 0.917) |
| | 1.25 | 0.1 | 0.790 (0.755, 0.822) | 0.786 (0.754, 0.817) | 0.792 (0.755, 0.822) |
| | | 0.3 | 0.780 (0.744, 0.814) | 0.775 (0.741, 0.808) | 0.784 (0.746, 0.815) |
| log(0.75) | 0.25 | 0.1 | 0.984 (0.981, 0.987) | 0.984 (0.981, 0.987) | 0.984 (0.981, 0.987) |
| | | 0.3 | 0.984 (0.981, 0.987) | 0.984 (0.981, 0.987) | 0.984 (0.981, 0.987) |
| | 0.75 | 0.1 | 0.897 (0.877, 0.914) | 0.894 (0.875, 0.912) | 0.895 (0.876, 0.912) |
| | | 0.3 | 0.897 (0.877, 0.914) | 0.893 (0.873, 0.911) | 0.894 (0.875, 0.911) |
| | 1.25 | 0.1 | 0.771 (0.735, 0.806) | 0.768 (0.733, 0.802) | 0.774 (0.735, 0.806) |
| | | 0.3 | 0.771 (0.733, 0.805) | 0.767 (0.732, 0.801) | 0.773 (0.735, 0.805) |

**Web Table 17** Median and interquartile range (IQR) (in parentheses) of relative efficiency (RE) as a function of treatment effect $\delta$, baseline prevalence, and different secular trends, when the true correlation model is exponential decay (ED) and the working correlation correctly specifies the true correlation model. Design factors considered are as follows: number of clusters $I = 24$, number of periods $J = 5$, mean cluster-period size $\overline{n} = 100$. The within-period intraclass correlation coefficient (WP-ICC) $\alpha_0$ is $0.05$, and the decay rate $\rho$ is $0.5$. Between cluster imbalance is measured by coefficient of variation, CV $\in \{0.25, 0.75, 1.25\}$. No within-cluster imbalance is introduced.

| $\delta$ | CV | Baseline prevalence | Constant secular trend | Increasing secular trend | Decreasing secular trend |
|---|---|---|---|---|---|
| log(0.35) | 0.25 | 0.1 | 0.989 (0.987, 0.991) | 0.989 (0.987, 0.991) | 0.989 (0.987, 0.991) |
| | | 0.3 | 0.989 (0.987, 0.991) | 0.989 (0.987, 0.991) | 0.989 (0.987, 0.991) |
| | 0.75 | 0.1 | 0.910 (0.892, 0.926) | 0.907 (0.889, 0.924) | 0.908 (0.890, 0.925) |
| | | 0.3 | 0.909 (0.891, 0.926) | 0.906 (0.887, 0.923) | 0.907 (0.889, 0.924) |
| | 1.25 | 0.1 | 0.776 (0.745, 0.809) | 0.777 (0.743, 0.805) | 0.777 (0.744, 0.808) |
| | | 0.3 | 0.774 (0.743, 0.807) | 0.775 (0.741, 0.803) | 0.776 (0.741, 0.806) |
| log(0.75) | 0.25 | 0.1 | 0.989 (0.987, 0.991) | 0.989 (0.987, 0.991) | 0.989 (0.987, 0.991) |
| | | 0.3 | 0.989 (0.987, 0.991) | 0.989 (0.987, 0.991) | 0.989 (0.987, 0.991) |
| | 0.75 | 0.1 | 0.908 (0.890, 0.925) | 0.905 (0.886, 0.923) | 0.906 (0.888, 0.922) |
| | | 0.3 | 0.908 (0.890, 0.925) | 0.905 (0.886, 0.923) | 0.906 (0.888, 0.922) |
| | 1.25 | 0.1 | 0.773 (0.741, 0.805) | 0.774 (0.739, 0.802) | 0.775 (0.740, 0.805) |
| | | 0.3 | 0.774 (0.741, 0.805) | 0.773 (0.739, 0.802) | 0.775 (0.740, 0.805) |

**Web Table 18** Median and interquartile range (IQR) (in parentheses) of relative efficiency (RE) as a function of treatment effect $\delta$, baseline prevalence, and different secular trends, when the true correlation model is exponential decay (ED) and the working correlation correctly is independence (IND). Design factors considered are as follows: number of clusters $I = 24$, number of periods $J = 5$, mean cluster-period size $\overline{n} = 100$. The within-period intraclass correlation coefficient (WP-ICC) $\alpha_0$ is $0.05$, and the decay rate $\rho$ is $0.9$. Between cluster imbalance is measured by coefficient of variation, CV $\in \{0.25, 0.75, 1.25\}$. No within-cluster imbalance is introduced.

| $\delta$ | CV | Baseline prevalence | Constant secular trend | Increasing secular trend | Decreasing secular trend |
|---|---|---|---|---|---|
| log(0.35) | 0.25 | 0.1 | 0.954 (0.931, 0.977) | 0.954 (0.927, 0.977) | 0.954 (0.929, 0.978) |
| | | 0.3 | 0.954 (0.931, 0.977) | 0.954 (0.927, 0.977) | 0.955 (0.929, 0.977) |
| | 0.75 | 0.1 | 0.721 (0.647, 0.789) | 0.714 (0.642, 0.778) | 0.721 (0.647, 0.78) |
| | | 0.3 | 0.722 (0.642, 0.791) | 0.714 (0.641, 0.779) | 0.723 (0.645, 0.78) |
| | 1.25 | 0.1 | 0.501 (0.421, 0.582) | 0.502 (0.421, 0.576) | 0.503 (0.427, 0.585) |
| | | 0.3 | 0.502 (0.422, 0.582) | 0.503 (0.423, 0.578) | 0.504 (0.427, 0.587) |
| log(0.75) | 0.25 | 0.1 | 0.953 (0.930, 0.977) | 0.953 (0.928, 0.977) | 0.954 (0.930, 0.977) |
| | | 0.3 | 0.954 (0.929, 0.976) | 0.953 (0.928, 0.977) | 0.954 (0.930, 0.977) |
| | 0.75 | 0.1 | 0.718 (0.641, 0.790) | 0.716 (0.642, 0.783) | 0.723 (0.645, 0.781) |
| | | 0.3 | 0.718 (0.641, 0.791) | 0.718 (0.641, 0.784) | 0.722 (0.644, 0.782) |
| | 1.25 | 0.1 | 0.503 (0.427, 0.580) | 0.501 (0.425, 0.582) | 0.504 (0.426, 0.583) |
| | | 0.3 | 0.503 (0.426, 0.581) | 0.500 (0.426, 0.582) | 0.505 (0.425, 0.584) |

**Web Table 19** Median and interquartile range (IQR) (in parentheses) of relative efficiency (RE) as a function of treatment effect $\delta$, baseline prevalence, and different secular trends, when the true correlation model is exponential decay (ED) and the working correlation correctly is independence (IND). Design factors considered are as follows: number of clusters $I = 24$, number of periods $J = 5$, mean cluster-period size $\overline{n} = 100$. The within-period intraclass correlation coefficient (WP-ICC) $\alpha_0$ is $0.05$, and the decay rate $\rho$ is $0.5$. Between cluster imbalance is measured by coefficient of variation, CV $\in \{0.25, 0.75, 1.25\}$. No within-cluster imbalance is introduced.

| $\delta$ | CV | Baseline prevalence | Constant secular trend | Increasing secular trend | Decreasing secular trend |
|---|---|---|---|---|---|
| log(0.35) | 0.25 | 0.1 | 0.954 (0.939, 0.970) | 0.954 (0.937, 0.969) | 0.954 (0.938, 0.969) |
| | | 0.3 | 0.954 (0.938, 0.970) | 0.954 (0.937, 0.969) | 0.954 (0.938, 0.970) |
| | 0.75 | 0.1 | 0.721 (0.661, 0.771) | 0.712 (0.655, 0.766) | 0.717 (0.660, 0.764) |
| | | 0.3 | 0.720 (0.661, 0.771) | 0.711 (0.655, 0.769) | 0.716 (0.661, 0.767) |
| | 1.25 | 0.1 | 0.499 (0.428, 0.561) | 0.495 (0.430, 0.562) | 0.500 (0.433, 0.570) |
| | | 0.3 | 0.499 (0.429, 0.561) | 0.497 (0.431, 0.561) | 0.500 (0.434, 0.570) |
| log(0.75) | 0.25 | 0.1 | 0.954 (0.938, 0.969) | 0.955 (0.937, 0.968) | 0.954 (0.938, 0.969) |
| | | 0.3 | 0.954 (0.938, 0.969) | 0.955 (0.937, 0.969) | 0.954 (0.938, 0.969) |
| | 0.75 | 0.1 | 0.719 (0.659, 0.773) | 0.711 (0.657, 0.768) | 0.716 (0.660, 0.768) |
| | | 0.3 | 0.718 (0.659, 0.773) | 0.712 (0.656, 0.768) | 0.717 (0.661, 0.768) |
| | 1.25 | 0.1 | 0.499 (0.431, 0.561) | 0.497 (0.430, 0.562) | 0.499 (0.433, 0.567) |
| | | 0.3 | 0.498 (0.432, 0.560) | 0.496 (0.432, 0.562) | 0.499 (0.432, 0.567) |

**Web Table 20** Median and interquartile range (IQR) (in parentheses) of relative efficiency (RE) as a function of treatment effect $\delta$, baseline prevalence, and different secular trends, when the true correlation model is exponential decay (ED) and the working correlation correctly specifies the true correlation model. Design factors considered are as follows: number of clusters $I = 24$, number of periods $J = 5$, mean cluster-period size $\overline{n} = 100$. The within-period intraclass correlation coefficient (WP-ICC) $\alpha_0$ is $0.05$, and the decay rate $\rho$ is $0.9$. Between cluster imbalance is measured by coefficient of variation, CV $\in \{0.25, 0.75, 1.25\}$. Within-cluster imbalance (pattern 4: randomly permuted) is introduced.

| $\delta$ | CV | Baseline prevalence | Constant secular trend | Increasing secular trend | Decreasing secular trend |
|---|---|---|---|---|---|
| log(0.35) | 0.25 | 0.1 | 0.943 (0.925, 0.959) | 0.940 (0.923, 0.955) | 0.942 (0.925, 0.959) |
| | | 0.3 | 0.934 (0.915, 0.953) | 0.930 (0.912, 0.948) | 0.935 (0.916, 0.952) |
| | 0.75 | 0.1 | 0.818 (0.762, 0.872) | 0.819 (0.769, 0.875) | 0.813 (0.763, 0.865) |
| | | 0.3 | 0.802 (0.741, 0.858) | 0.799 (0.743, 0.859) | 0.797 (0.743, 0.852) |
| | 1.25 | 0.1 | 0.637 (0.560, 0.715) | 0.640 (0.562, 0.710) | 0.638 (0.576, 0.725) |
| | | 0.3 | 0.611 (0.531, 0.692) | 0.613 (0.532, 0.687) | 0.618 (0.550, 0.699) |
| log(0.75) | 0.25 | 0.1 | 0.926 (0.905, 0.945) | 0.923 (0.904, 0.942) | 0.924 (0.906, 0.943) |
| | | 0.3 | 0.924 (0.904, 0.943) | 0.922 (0.902, 0.940) | 0.922 (0.904, 0.942) |
| | 0.75 | 0.1 | 0.787 (0.721, 0.843) | 0.784 (0.726, 0.848) | 0.777 (0.719, 0.835) |
| | | 0.3 | 0.784 (0.719, 0.841) | 0.781 (0.725, 0.845) | 0.774 (0.717, 0.833) |
| | 1.25 | 0.1 | 0.593 (0.513, 0.672) | 0.594 (0.518, 0.672) | 0.597 (0.524, 0.679) |
| | | 0.3 | 0.590 (0.511, 0.668) | 0.592 (0.515, 0.670) | 0.594 (0.521, 0.675) |

**Web Table 21** Median and interquartile range (IQR) (in parentheses) of relative efficiency (RE) as a function of treatment effect $\delta$, baseline prevalence, and different secular trends, when the true correlation model is exponential decay (ED) and the working correlation correctly specifies the true correlation model. Design factors considered are as follows: number of clusters $I = 24$, number of periods $J = 5$, mean cluster-period size $\overline{n} = 100$. The within-period intraclass correlation coefficient (WP-ICC) $\alpha_0$ is $0.05$, and the decay rate $\rho$ is $0.5$. Between cluster imbalance is measured by coefficient of variation, CV $\in \{0.25, 0.75, 1.25\}$. Within-cluster imbalance (pattern 4: randomly permuted) is introduced.

| $\delta$ | CV | Baseline prevalence | Constant secular trend | Increasing secular trend | Decreasing secular trend |
|---|---|---|---|---|---|
| log(0.35) | 0.25 | 0.1 | 0.963 (0.953, 0.972) | 0.961 (0.952, 0.969) | 0.962 (0.952, 0.971) |
| | | 0.3 | 0.961 (0.950, 0.970) | 0.959 (0.949, 0.967) | 0.960 (0.950, 0.969) |
| | 0.75 | 0.1 | 0.875 (0.832, 0.914) | 0.878 (0.838, 0.913) | 0.872 (0.826, 0.908) |
| | | 0.3 | 0.871 (0.828, 0.911) | 0.873 (0.831, 0.911) | 0.868 (0.823, 0.905) |
| | 1.25 | 0.1 | 0.724 (0.658, 0.790) | 0.730 (0.662, 0.790) | 0.728 (0.668, 0.797) |
| | | 0.3 | 0.722 (0.649, 0.784) | 0.723 (0.657, 0.785) | 0.724 (0.662, 0.792) |
| log(0.75) | 0.25 | 0.1 | 0.958 (0.948, 0.968) | 0.957 (0.947, 0.966) | 0.958 (0.947, 0.967) |
| | | 0.3 | 0.958 (0.947, 0.967) | 0.956 (0.946, 0.965) | 0.957 (0.947, 0.967) |
| | 0.75 | 0.1 | 0.868 (0.824, 0.907) | 0.870 (0.826, 0.908) | 0.863 (0.818, 0.903) |
| | | 0.3 | 0.867 (0.824, 0.906) | 0.869 (0.825, 0.908) | 0.863 (0.818, 0.902) |
| | 1.25 | 0.1 | 0.720 (0.644, 0.780) | 0.720 (0.657, 0.782) | 0.719 (0.657, 0.788) |
| | | 0.3 | 0.717 (0.644, 0.782) | 0.719 (0.656, 0.781) | 0.719 (0.655, 0.788) |

**Web Table 22** Median and interquartile range (IQR) (in parentheses) of relative efficiency (RE) as a function of treatment effect $\delta$, baseline prevalence, and different secular trends, when the true correlation model is exponential decay (ED) and the working correlation correctly is independence (IND). Design factors considered are as follows: number of clusters $I = 24$, number of periods $J = 5$, mean cluster-period size $\overline{n} = 100$. The within-period intraclass correlation coefficient (WP-ICC) $\alpha_0$ is $0.05$, and the decay rate $\rho$ is $0.9$. Between cluster imbalance is measured by coefficient of variation, CV $\in \{0.25, 0.75, 1.25\}$. Within-cluster imbalance (pattern 4: randomly permuted) is introduced.

| $\delta$ | CV | Baseline prevalence | Constant secular trend | Increasing secular trend | Decreasing secular trend |
|---|---|---|---|---|---|
| log(0.35) | 0.25 | 0.1 | 0.920 (0.893, 0.947) | 0.916 (0.886, 0.944) | 0.920 (0.892, 0.949) |
| | | 0.3 | 0.917 (0.891, 0.944) | 0.914 (0.884, 0.941) | 0.917 (0.889, 0.947) |
| | 0.75 | 0.1 | 0.799 (0.728, 0.874) | 0.805 (0.725, 0.874) | 0.790 (0.725, 0.864) |
| | | 0.3 | 0.797 (0.726, 0.870) | 0.800 (0.722, 0.872) | 0.788 (0.721, 0.862) |
| | 1.25 | 0.1 | 0.649 (0.551, 0.744) | 0.642 (0.550, 0.740) | 0.650 (0.560, 0.742) |
| | | 0.3 | 0.646 (0.554, 0.744) | 0.641 (0.553, 0.739) | 0.652 (0.558, 0.743) |
| log(0.75) | 0.25 | 0.1 | 0.914 (0.888, 0.942) | 0.911 (0.881, 0.938) | 0.914 (0.886, 0.944) |
| | | 0.3 | 0.914 (0.887, 0.941) | 0.911 (0.881, 0.938) | 0.913 (0.885, 0.943) |
| | 0.75 | 0.1 | 0.794 (0.723, 0.865) | 0.798 (0.721, 0.868) | 0.787 (0.719, 0.862) |
| | | 0.3 | 0.794 (0.720, 0.864) | 0.797 (0.721, 0.868) | 0.787 (0.719, 0.861) |
| | 1.25 | 0.1 | 0.642 (0.553, 0.736) | 0.638 (0.557, 0.737) | 0.647 (0.562, 0.741) |
| | | 0.3 | 0.643 (0.554, 0.735) | 0.639 (0.558, 0.734) | 0.648 (0.562, 0.740) |

**Web Table 23** Median and interquartile range (IQR) (in parentheses) of relative efficiency (RE) as a function of treatment effect $\delta$, baseline prevalence, and different secular trends, when the true correlation model is exponential decay (ED) and the working correlation correctly is independence (IND). Design factors considered are as follows: number of clusters $I = 24$, number of periods $J = 5$, mean cluster-period size $\overline{n} = 100$. The within-period intraclass correlation coefficient (WP-ICC) $\alpha_0$ is $0.05$, and the decay rate $\rho$ is $0.5$. Between cluster imbalance is measured by coefficient of variation, CV $\in \{0.25, 0.75, 1.25\}$. Within-cluster imbalance (pattern 4: randomly permuted) is introduced.

| $\delta$ | CV | Baseline Prevalence | Constant Secular trend | Increasing Secular trend | Decreasing Secular trend |
|---|---|---|---|---|---|
| log(0.35) | 0.25 | 0.1 | 0.900 (0.879, 0.918) | 0.895 (0.873, 0.914) | 0.899 (0.877, 0.920) |
| | | 0.3 | 0.897 (0.877, 0.916) | 0.893 (0.872, 0.913) | 0.897 (0.875, 0.918) |
| | 0.75 | 0.1 | 0.755 (0.687, 0.812) | 0.760 (0.687, 0.815) | 0.746 (0.688, 0.806) |
| | | 0.3 | 0.753 (0.688, 0.809) | 0.756 (0.687, 0.815) | 0.744 (0.686, 0.805) |
| | 1.25 | 0.1 | 0.581 (0.491, 0.668) | 0.577 (0.500, 0.664) | 0.585 (0.498, 0.665) |
| | | 0.3 | 0.584 (0.493, 0.668) | 0.577 (0.503, 0.667) | 0.586 (0.498, 0.666) |
| log(0.75) | 0.25 | 0.1 | 0.895 (0.875, 0.913) | 0.891 (0.870, 0.911) | 0.894 (0.873, 0.915) |
| | | 0.3 | 0.895 (0.874, 0.913) | 0.891 (0.870, 0.910) | 0.894 (0.873, 0.915) |
| | 0.75 | 0.1 | 0.752 (0.687, 0.806) | 0.753 (0.686, 0.813) | 0.744 (0.686, 0.805) |
| | | 0.3 | 0.750 (0.686, 0.806) | 0.752 (0.689, 0.812) | 0.742 (0.685, 0.805) |
| | 1.25 | 0.1 | 0.586 (0.495, 0.662) | 0.579 (0.507, 0.667) | 0.587 (0.498, 0.669) |
| | | 0.3 | 0.586 (0.496, 0.662) | 0.578 (0.506, 0.664) | 0.587 (0.498, 0.668) |

#### Web Appendix E. RE of GEE analysis under the true versus independence working correlation model

Figures in this section provide insight about the relative efficiency under the true versus independence working correlation model defined in Section 8 of the main article.

**Web Figure 45** Median of new defined relative efficiency (RE) under the true versus independence (IND) working correlation model as a function of the within-period intraclass correlation coefficient (WP-ICC) $\alpha_0 \in \{0.01, 0.05, 0.1, 0.2\}$ and the ratio of between-period intraclass correlation coefficient (BP-ICC) to WP-ICC, $\alpha_1/\alpha_0 \in [0, 1]$, when the true correlation model is nested exchangeable (NEX). Design factors considered are as follows: number of clusters $I = 12$ and 96, number of periods $J = 5$ and 13, and mean cluster-period sizes $\overline{n} = 100$ and 300.

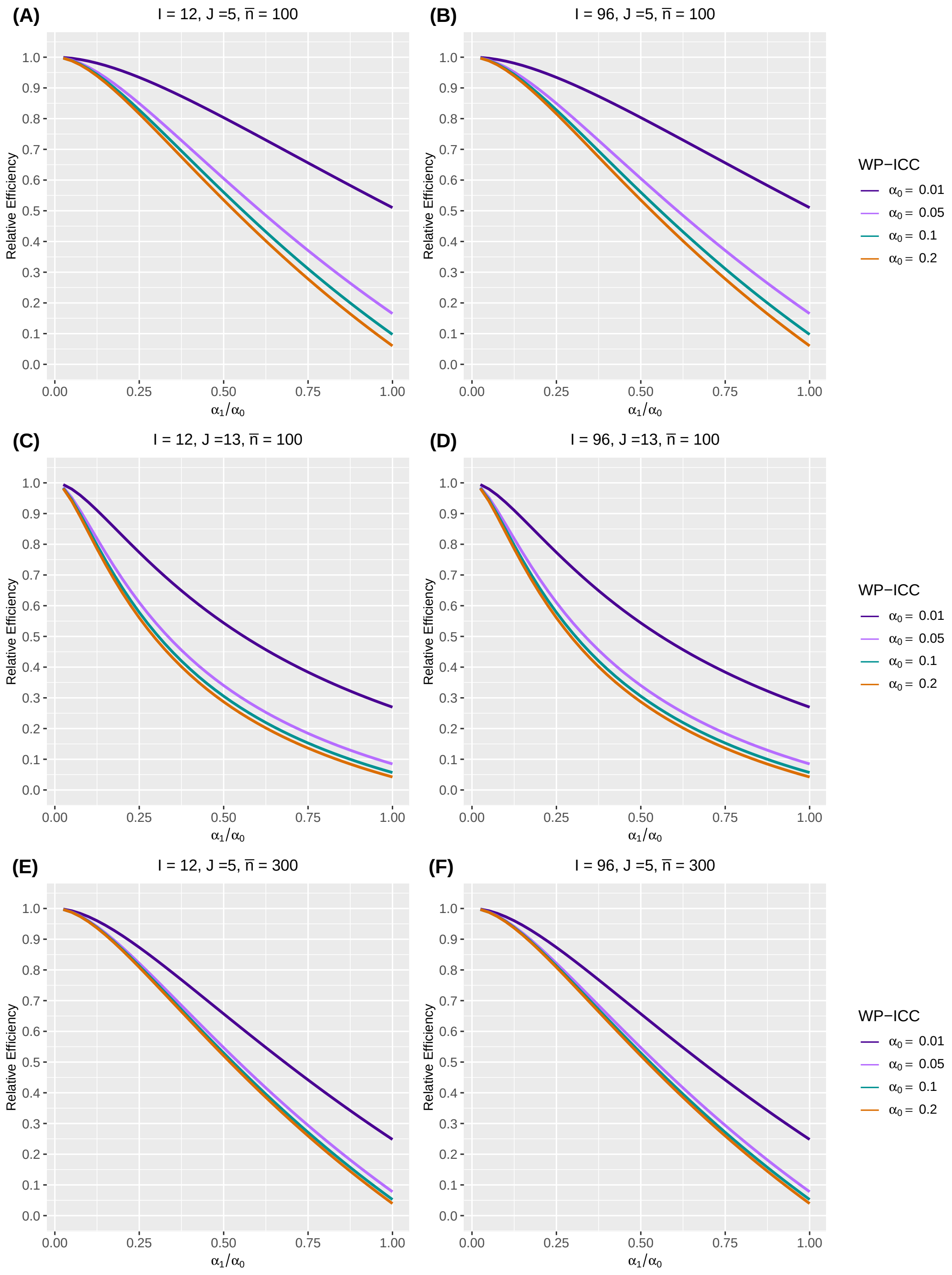

**Web Figure 46** Median of new defined relative efficiency (RE) under the true versus independence (IND) working correlation model as a function of the within-period intraclass correlation coefficient (WP-ICC) $\alpha_0 \in \{0.01, 0.05, 0.1, 0.2\}$ and the decay rate $\rho \in [0, 1]$, when the true correlation model is exponential decay (ED). Design factors considered are as follows: number of clusters $I = 12$ and 96, number of periods $J = 5$ and 13, and mean cluster-period sizes $\overline{n} = 100$ and 300.

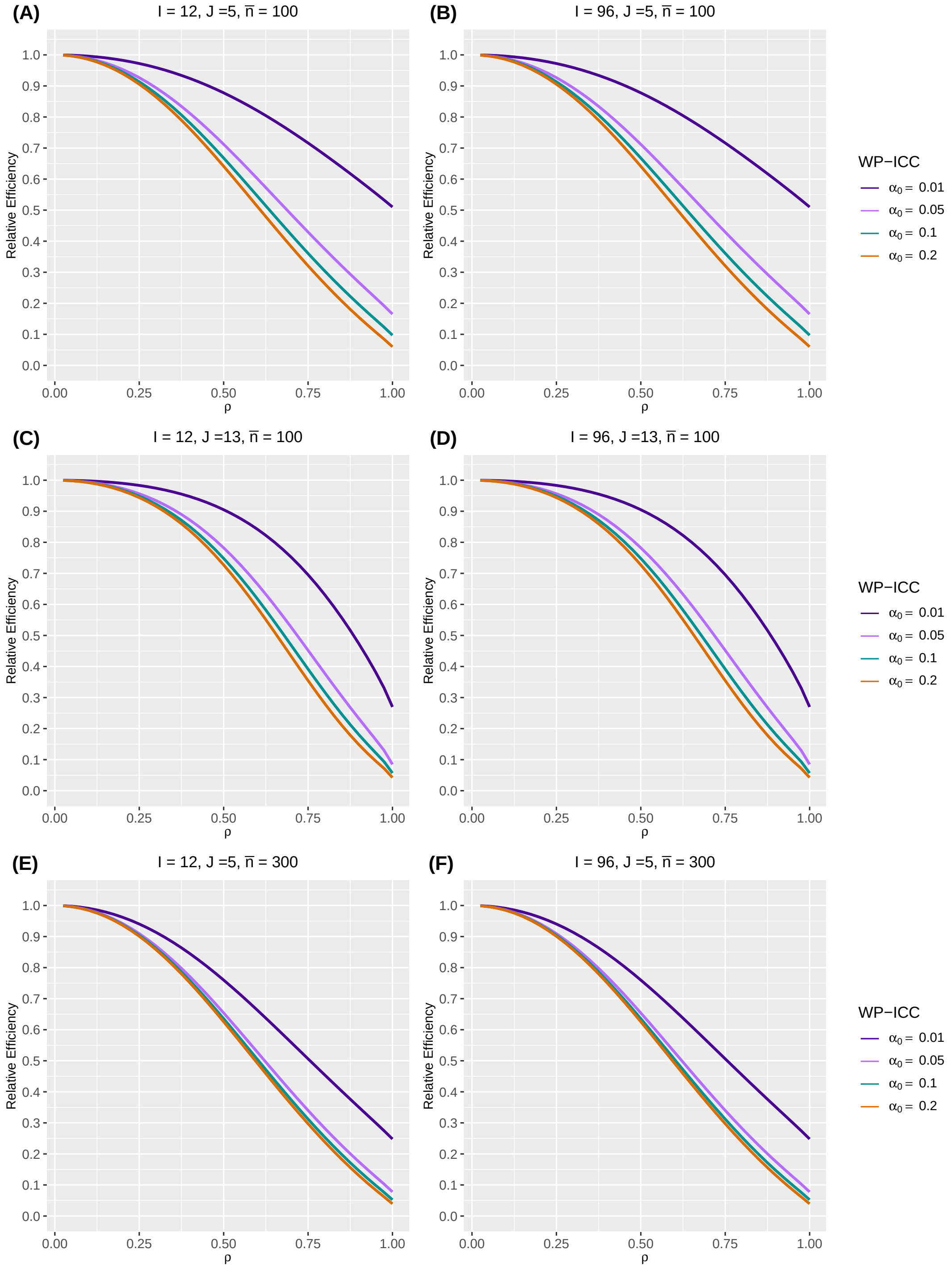

#### Web Appendix F. Compare simulation-based RE and the reciprocal of the conservative inflation factor

Figures in this section compare the relative efficiency in our simulations and the reciprocal of a conservative inflation factor $1 + \mathrm{CV}^2$ mentioned in Section 8 of the main article. CV denotes the coefficient of variation that measures the between-cluster imbalance.

**Web Figure 47** The median and interquartile range (IQR) of relative efficiency (RE) as a function of coefficient of variation (CV) measuring between-cluster imbalance, when the true correlation model is nested exchangeable (NEX). Design factors considered are as follows: number of clusters $I = 12$ and $96$, number of periods $J = 5$. The within-period intraclass correlation coefficient (WP-ICC) $\alpha_0 = 0.05$, and between-period intraclass correlation coefficient (BP-ICC) $\alpha_1 \in \{0.001, 0.025, 0.05\}$. No within-cluster imbalance is introduced. The broken grey curve shows the reciprocal of the conservative inflation factor $1 + CV^2$.

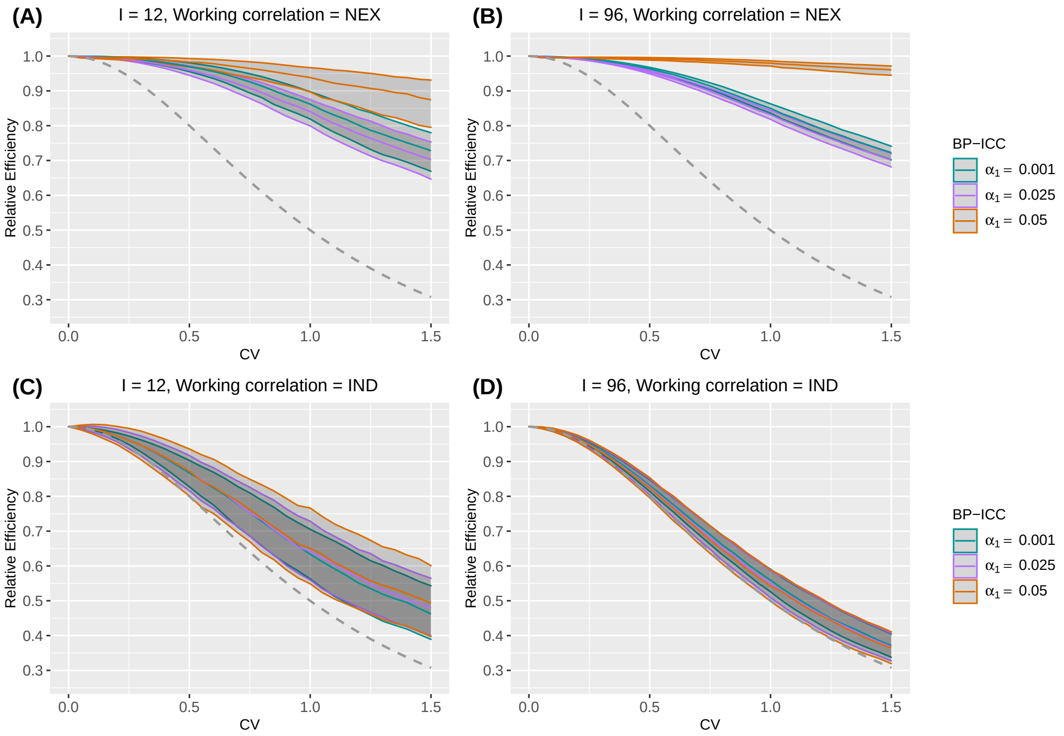

**Web Figure 48** The median and interquartile range (IQR) of relative efficiency (RE) as a function of coefficient of variation (CV) measuring between-cluster imbalance, when the true correlation model is nested exchangeable (NEX). Design factors considered are as follows: number of clusters $I = 24$ and $48$, number of periods $J = 5$. The within-period intraclass correlation coefficient (WP-ICC) $\alpha_0 = 0.05$, and between-period intraclass correlation coefficient (BP-ICC) $\alpha_1 \in \{0.001, 0.025, 0.05\}$. No within-cluster imbalance is introduced. The broken grey curve shows the reciprocal of the conservative inflation factor $1 + CV^2$.

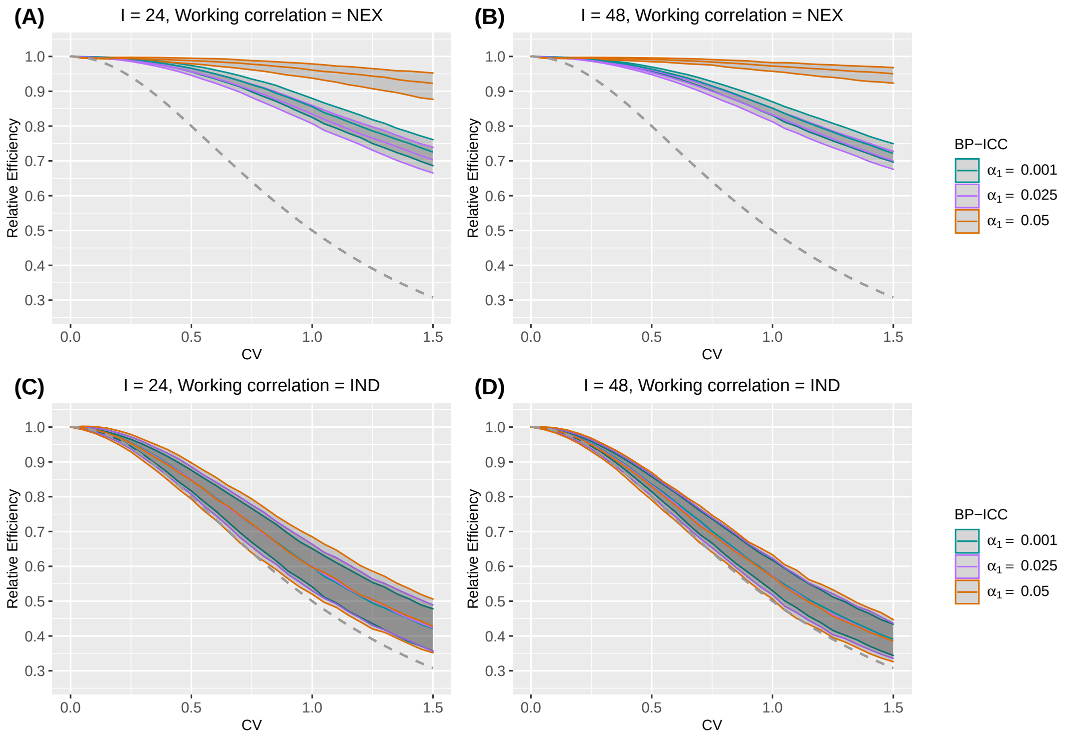

**Web Figure 49** The median and interquartile range (IQR) of relative efficiency (RE) as a function of coefficient of variation (CV) measuring between-cluster imbalance, when the true correlation model is exponential decay (ED). Design factors considered are as follows: number of clusters $I = 12$ and 96, number of periods $J = 5$. The within-period intraclass correlation coefficient (WP-ICC) $\alpha_0 = 0.05$, and decay parameter $\rho \in \{0.1, 0.5, 1\}$. No within-cluster imbalance is introduced. The broken grey curve shows the reciprocal of the conservative inflation factor $1 + CV^2$.

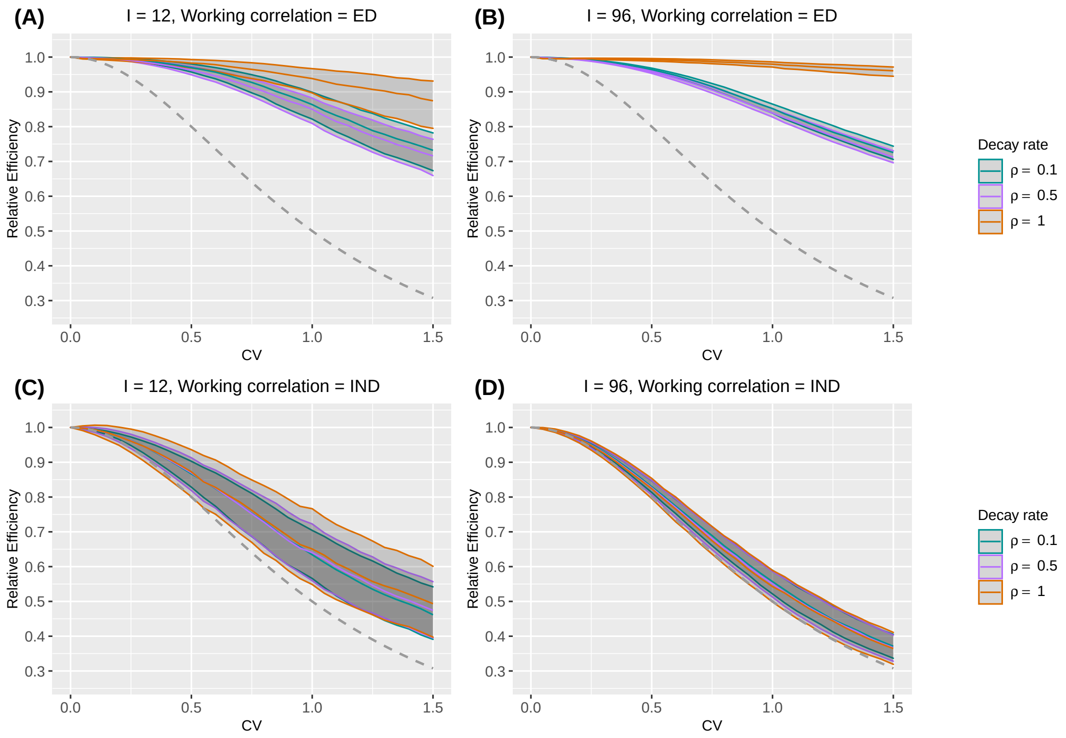

**Web Figure 50** The median and interquartile range (IQR) of relative efficiency (RE) as a function of coefficient of variation (CV) measuring between-cluster imbalance, when the true correlation model is exponential decay (ED). Design factors considered are as follows: number of clusters $I = 24$ and $48$, number of periods $J = 5$. The within-period intraclass correlation coefficient (WP-ICC) $\alpha_0 = 0.05$, and decay parameter $\rho \in \{0.1, 0.5, 1\}$. No within-cluster imbalance is introduced. The broken grey curve shows the reciprocal of the conservative inflation factor $1 + CV^2$.

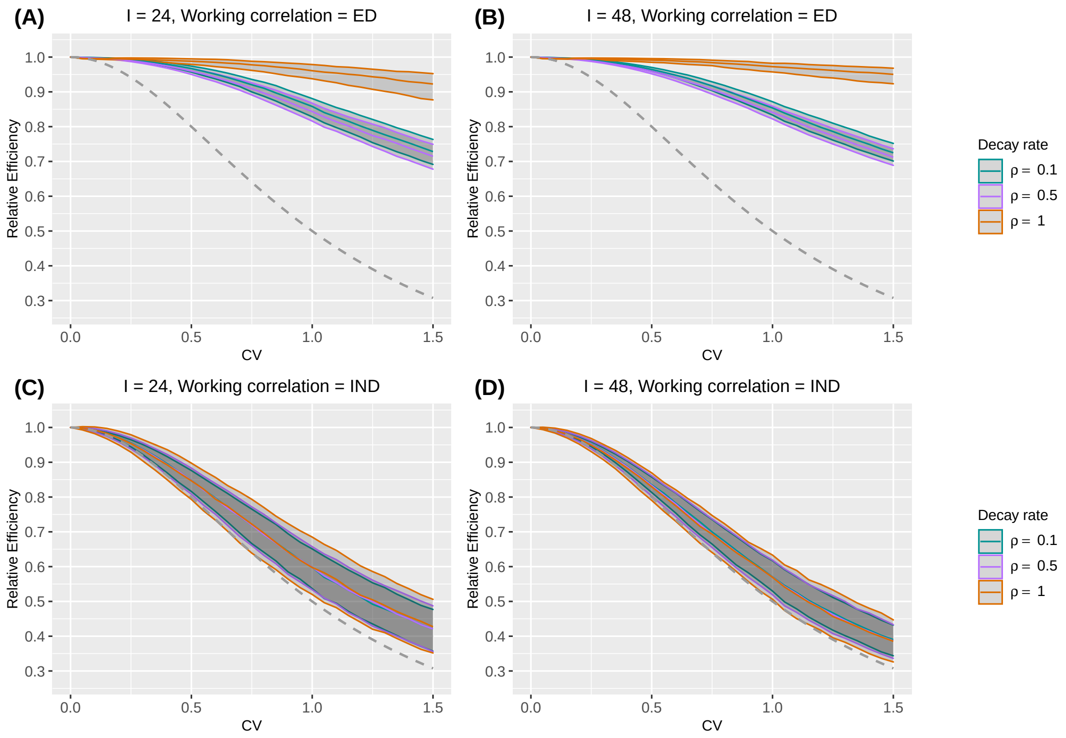
